# Safety of praziquantel in persons with and without schistosomiasis: systematic review and meta-analysis

**DOI:** 10.1101/2022.03.09.22270839

**Authors:** Anthony Danso-Appiah, David Owiredu, Morrison Asiamah, Kwadwo Akuffo, Paolo Eusebi, Guo Jiangang, Pauline Mwinzi, Daniel G. Colley, Paul Hagan, M. Hassan Murad, Amadou Garba Djirmay

**Author notes:** Correspondence: Anthony Danso-Appiah.

## Abstract

Millions of praziquantel doses have been delivered in schistosomiasis endemic populations through preventive chemotherapy. However, no comprehensive assessment of short and long-term safety has been conducted. This systematic review assessed safety of praziquantel in persons with and without schistosome infections who received praziquantel treatment.

**Methods:** We identified relevant studies (published, unpublished, in press or preprint) that assessed safety of praziquantel without language restriction. We searched MEDLINE, EMBASE, CINAHL, and LILACS from 1978 to 31^st^ October 2021, using well-formulated and piloted search strategy. We also searched the Cochrane Infectious Diseases Group Specialized Register, CENTRAL (The Cochrane Library 2021), mRCT, Google Scholar, Hinari and Africa Journals Online. References of relevant studies were checked and experts were contacted for additional studies. One author searched and managed the search output. Two authors selected studies, extracted data and assessed quality of the included studies for risk of bias. Activities at all stages were checked independently by a third reviewer. Discrepancies were resolved through discussion among the authors. Data were analysed with RevMan v5.4 and STATA v17. Binary outcomes were reported as risk ratio using random-effects model and continuous outcomes as mean difference, all presented with their 95% confidence intervals. P-value was set at 0.05. Heterogeneity was assessed using I^2^-static and where possible sensitivity analysis was conducted. When pooling of data was not possible, we presented data in a narrative synthesis and as tables.

**Main results:** The search retrieved 3202 studies of which 134 met the inclusion criteria; 94 (70.1%) were conducted in Africa, 17 (12.7%) in Asia, 15 (11.2%) in the Americas (14 from Brazil), 4 (3.0%) in the Middle East and 3 (2.2%) in Europe. Praziquantel mostly resulted in mild-to-moderate and transient adverse events, however, majority of the included studies had design issues, including very short follow-up times (mostly few hours) for assessing incidence of adverse events. Less than <10% of the studies reported severe or serious adverse events. The subgroup analyses of twenty studies comparing school age children (SAC) and adults, and involved over one million participants found no difference in the nature of adverse events, but SAC experienced higher incidence than adults: headache (RR 3.07, 95% CI 2.32 to 4.06, twenty studies, I^2^ = 98%, p < 0.00001), dizziness (RR 1.80, 95% CI 1.36 to 2.37, p = 0.0001), vomiting (RR 2.43, 95% CI 1.87 to 3.14, I^2^ = 98%; p < 0.00001), four time for abdominal pain (RR 3.97, 95% CI 3.09 to 5.10, I^2^ = 96%, p < 0.00001), nausea (RR 1.67, 95% CI 1.32 to 2.12, I^2^ = 97%, p < 0.0001), general discomfort (RR 1.32, 95% CI 1.03 to 1.68, I^2^ = 97%, p < 0.00001), fever (RR 4.78, 95% CI 3.04 to 7.52, I^2^ = 98%, p < 0.00001), diarrhoea (RR 1.41, 95% CI 1.12 to 1.78, I^2^ = 92%, p < 0.00001), itching (RR 2.42, 95% CI 1.58 to 3.70, I^2^ = 93%, p <0.0001) and breathing difficulty (RR 2.46, 95% CI 1.41 to 4.29, I^2^ = 92%, p = 0.002). There was no statistically significant difference in incidence of swelling. Some of the studies that assessed safety in pregnant women reported serious events including miscarriages, foetal deaths and congenital anomalies, but the evidence is incoclusive given the limited numbers. Some studies reported praziquantel-related visual adverse events, but evidence is limited and remains inconclusive. There was paucity of data on long term adverse events, and events in co-morbidity, polypharmacy, co-infection with taeniasis. Generally, adverse events research in this area lacked methodological rigour.

**Conclusions:** The evidence generated from this review involving millions of people and millions of doses from different geographic locations with mostly mild-to-moderate and transient adverse events shows praziquantel is safe. However, given that the primary studies included in the review had design issues, including over 95% assessing adverse events over very short follow-up times, means serious long-term adverse events would have been missed. Also, the fact that some pregnant women who received praziquantel experienced serious events including miscarriages, foetal deaths and congenital anomalies calls for caution in the inclusion of pregnant women, particularly in their first trimester, in preventive chemotherapy campaigns. Additionally, the studies that reported severe visual adverse events raise safety concerns. Praziquantel is now offered repeatedly in endemic communities and the fact that in some settings up to 90% of those without infection could be offered the drug and the fact that there was no study that compared safety between infected and non-infected recipients, warrants further research. Evidence on safety in pregnant women and their foetuses, co-morbidity, polypharmacy, co-infection with taeniasis, as well as co-administration with drugs used in other preventive chemotherapy programmes, remain inconclusive and further research with long follow-up that should include blood chemistry analysis to provide additional evidence on long term safety, is warranted. This systematic review has exposed the lack of methodological rigour in adverse events studies and recommends future research should use robust and standardized design, methods, conduct and reporting.

## Background

Schistosomiasis is caused by the blood fluke, a group of flat worms that reside in the blood vessels in the human hosts and presents as an acute, but mostly chronic illness (WHO 2002). It has been estimated that around 800 million people are at risk of the infection worldwide and over 250 million may be infected (Steinmann et al. 2006, King et at 2015, WHO 2021, WHO 2022). In 2018, it was estimated that over 230 million infected people required preventive chemotherapy (PC) worldwide (Lo et al. 2018; WHO 2021). Groups at risk for schistosomiasis are preschool-aged children (PSAC), school-aged children (SAC), adults in certain occupational groups such as farmers who rely on water bodies and irrigations for their work, women who are in contact with infected water sources for domestic activities, and entire communities in high-risk areas (WHO 2021). Around 90% of the burden of the disease is concentrated in Sub Saharan Africa (SSA) (WHO 2022).

Preventive chemotherapy (PC), started in the early 2000s as the control strategy recommended by the WHO, is applied in most endemic countries (WHO 2002, WHO 2006, WHO 2012, WHO 2013, WHO 2021). Within the PC concept, endemic countries are urged to embark on mass drug administration (MDA) with praziquantel (PZQ) at a single 40 mg/kg oral dose using infection prevalence thresholds. Several drugs have been used or tried in treating schistosomiasis and later abandoned because of adverse events or poor effect (Cioli 1995, more references). There is no effective antischistosomal vaccine as yet (Gryseels 2000; Fenwick 2006a; McManus 2008; Colley and Secor 2014; Fonseca et al. 2015) and control will continue to rely on praziquantel, a broad spectrum antischistosomal drug introduced on to the market in the early 1980s, with no real alternative. The common adverse effects reported across studies are abdominal pain, nausea, vomiting and diarrhoea, which are usually mild-to-moderate and transient, lasting less than 24 hours.

The neglected tropical disease (NTD) earlier road map and the London Declaration of NTD moved partners to pledge commitment towards supporting the fight against schistosomiasis to reach a target of 75-100% coverage of school-aged children living in endemic regions by 2020, and to eliminate the disease in some regions (reference). The support led to increased donation of PZQ to reach excesses of 250 million tablets per year to take care of the 100 million people that needed treatments. The most recent data estimated the number of people that required PZQ for schistosomiasis globally in 2020 at 241.3 million across 51 countries (133.3 million SAC and 108 million adults), an increase over that in 2019 (235.4 million) in 2019 (WHO 2021). Africa remains the highest burden of schistosomiasis (>90% of the people who require PC for schistosomiasis) according to the latest mapping estimate (WHO 2018, WHO 2020, WHO 2021). In January 2021, WHO launched a new road map to guide action against NTDs and to set the targets achievable for all NTDs by 2030 (WHO 2021). The targets for 2030 are elimination of schistosomiasis and STH as public health problems in all endemic countries and interruption of schistosome transmission in humans in selected countries (WHO 2021).

In the PC strategy, WHO recommends PZQ treatment of all persons in endemic communities with infection levels reaching 10% threshold whether infected or without infection (Danso-Appiah et al. 2022; WHO 2021). Treating individuals without infection (that could reach up to 90% of the village’s population) has moral and ethical implications for public health. Additionally, the recommendation to treat pregnant and lactating women, for example, lacks strong evidence (WHO 2022). Also, in people coinfected with other infections such as taeniasis, or with mixed infections with the different types of schistosomiasis in the same individual, with co-morbidities, or the very young children and infants, the evidence is weak and inconclusive (WHO 2022). Meanwhile, the donation and use of PZQ will continue to increase and there is urgent need to comprehensively assess the overall safety profile of PZQ, particularly its long-term safety profile as this has not been investigated at all or less sufficiently.

The WHO commissioned this systematic review and meta-analysis in 2018 to comprehensively assess both the short- and long-term safety profile of PZQ from comparative and non-comparative studies across schistosomiasis endemic settings that reported adverse events. Specifically, we assessed adverse events of PZQ by dose (40, 50, 60, 70 mg and 80 mg/kg), and when drugs are co-administered with PZQ e.g. Albendazole, Mebendazole, Pyrantel pamoate or Levamisole (for soil-transmitted helminthiasis), or Albendazole plus either Ivermectin or Diethylcarbamazine [DEC] (for lymphatic filariasis) or ACTs (for malaria). Adverse events were quantified, where possible, and classified based on severity (as mild, moderate, severe or very severe) and reporting any occurrence of serious adverse events (SAEs, i.e. fatal events, life-threatening, requiring hospitalization, requiring discontinuation of treatment, or resulting in disability or incapacity).

## REVIEW METHODS

The review was conducted as part of WHO first evidence-based guideline that provided the baseline evidence on which the guideline recommendations were based. Various stakeholders were involved in formulating the review question and the inclusion/exclusion criteria. The review follows the Preferred Reporting Items for Systematic Reviews and Meta-Analyses (PRISMA) Statement (Page et al. 2020) and Meta-analysis of Observational studies in Epidemiology (MOOSE) (Stroup et al. 2000). The review protocol was registered in the International Prospective Register of Systematic Reviews (PROSPERO) with the ID (CRD42018085695).

### Criteria for considering studies for this review

#### Types of studies

Published and unpublished comparative and non-comparative studies that investigated AEs of PZQ treatment.

#### Types of participants

Whole communities or subgroups (preschool age children, school age children, adults including pregnant and lactating women) living in an endemic area and infected with any of the following schistosome species *S. mansoni*, *S. haematobium*, *S. japonicum*, *S. intercalatum* and *S. mekongi* or non-infected, who received PZQ treatment at a single dose of 40 mg/kg or a higher dose (50, 60, 70, or 80 mg/kg).

#### Intervention

1. PZQ at a single oral dose (40 mg/kg)

#### Control

1. Placebo.
2. PZQ divided or higher doses (50, 60, 70, or 80 mg/kg).
3. Medicines co-administered with PZQ such as albendazole, mebendazole, pyrantel pamoate or levamisole (for soil-transmitted helminthiasis), or albendazole plus either ivermectin or diethylcarbamazine [DEC] (for lymphatic filariasis), ACTs (for malaria), or Azithromycin (Zithromax) for trachoma.
4. No treatment.

#### Outcomes

##### Primary adverse events

- Serious clinical adverse events.
- Serious maternal and foetal events.
- Long term adverse events including tissue organ pathology.

##### Secondary adverse events

- All non-serious adverse events.
- Non-serious maternal and foetal events.

### Adverse Events and related terms defined

*Adverse Event (AE)* or Adverse Experience is any untoward medical occurrence in a patient or clinical investigation subject administered pharmaceutical product and which does not necessarily have to have a causal relationship with this treatment. This is different from *Adverse Drug Reaction (ADR)* which represents all noxious and unintended responses to a medicinal product related to any dose should be considered adverse drug reaction. *Unexpected Adverse Drug Reaction (UADR)* is an adverse reaction, the nature or severity of which is not consistent with the applicable product information (e.g., IB for an unapproved investigational medicinal product). *Serious adverse event (SAE)* or experience or reaction encompasses any untoward medical occurrence that at any dose 1) results in death, is life-threatening (an event in which the patient was at risk of death at the time of the event; not an event which hypothetically might have caused death if it were more severe), or requires inpatient hospitalization or prolongation of existing hospitalization or results in persistent or significant disability/incapacity or a congenital anomaly/birth defect.

### Search strategy for identification of studies

We identified all relevant studies without language restrictions or publication status (published, unpublished, in press, and ongoing). We searched MEDLINE, EMBASE and LILACS from 1978 to 31^st^ October 2021, with no language restrictions. Praziquantel was first tried in humans around 1978 and was introduced onto the market around 1979/1980, hence the date restriction. Search strategies were developed using the major concepts: Praziquantel (all brands), preventive chemotherapy, adverse events, adverse drug reactions, side effects, tolerance and schistosomiasis (including the five types infecting humans - *S. mansoni*, *S. haematobium*, *S. japonicum*, *S. intercalatum* and *S. mekongi*). We also searched the Cochrane Infectious Diseases Group Specialized Register, CENTRAL (The Cochrane Library 2021), mRCT, Hinari, Google Scholar, WHO Library Database and Africa Journals Online. The reference lists of articles were reviewed, and experts in the field of schistosomiasis were contacted for additional or unpublished studies. The results of the database searches were stored and de-duplicated in EndNote X9 library and exported to Rayyan for study selection.

### Study selection

Two reviewers screened the results of the search output to identify studies meeting the pre-specified inclusion criteria using a pre-tested study selection flow chart [see Additional file 1]. We included all primary studies (RCTs, quasi-experimental studies, cohort, cross-sectional, case series and case studies). We excluded reviews, opinions or commentaries. Screening was conducted in two stages using the review management software Rayyan (Ouzzani et al. 2016). The first stage screening focused on the titles and abstracts and the second on full texts. Full texts of all potentially relevant studies were retrieved and assessed in detail for inclusion. A screening guide was used to ensure that independent reviewers applied the selection criteria reliably. For authors whose full-text documents were not available via the various internet-based sources, we attempted to contact them directly through the corresponding author’s address. If information required to make the decision for inclusion was not obtained, the article was excluded from the analysis. Discrepancies were resolved through discussion between the reviewers.

### Data extraction and management

Two reviewers extracted data using a pre-tested data extraction form. Study characteristics such as the country and year in which the study was conducted, the study design and the methods; information on adverse events such as clinical, biochemical or pathological events, and severity and duration; and epidemiological and demographic data – e.g., endemicity status, region where the study was conducted, participants’ prior treatment status, target population, sex, age and number of participants, were extracted. When necessary, we contacted the authors of the published articles on included studies to see if they could clarify or supplement the published results or provide raw data that we could use. In case of multinational studies, we separated the results by country. However, where it was not possible to separate the data by country, the aggregated data was presented and the countries in which the study was conducted were shown. When two or more articles were published from the same study, we selected the study with most data as the parent study and linked the rest. Data were managed with Excel and transported to RevMan for analysis. Discrepancies were resolved through discussion between the reviewers.

### Assessment of methodological quality

One reviewer assessed the risk of bias of each trial using The Cochrane Collaboration’s risk of bias tool for RCTs (Higgins et al. 2021) and the assessment results was verified independently by another reviewer. Where information in the trial report was unclear, we contacted the trial authors for clarification. We assessed the risk of bias for six domains: sequence generation, allocation concealment, blinding (investigators, outcome assessors and participants), incomplete outcome data, selective outcome reporting and other sources of bias. For each domain, we made a judgment of ’low risk’ of bias, ’high risk’ of bias or ’unclear’ risk of bias. ROBINS-I (Risk Of Bias In Non-randomised Studies - of Interventions), was used to evaluate risk of bias in estimates of the comparative harm or benefit of interventions from studies that did not use randomisation to allocate units (individuals or clusters of individuals) to comparison groups (Sterne et al. 2021). Seven domains through which bias might be introduced into a NRSI was considered. The first two domains, covering confounding and selection of participants into the study, address issues before the start of the interventions that were compared (“baseline”). The third domain addressed classification of the interventions themselves. The other four domains addressed issues after the start of interventions: biases due to deviations from intended interventions, missing data, measurement of outcomes, and selection of the reported result. The STROBE checklist was used to evaluate the reporting methodology in each observational comparative study (Von Elm et al. 2007). Risk of bias and quality results was presented in a table. The Risk of Bias Tool for Prevalence Studies (Hoy et al. 2012) was used to assess the methodological quality and risk of bias for each prevalence study. We resolved any discrepancies by discussion between the authors.

### Data synthesis

The absolute rates of adverse events were calculated from all studies and relative risks from comparative trials. Data were analysed and presented as risk ratio (for dichotomous outcomes) or mean difference (for continuous outcomes) with their 95% confidence intervals (CIs). The meta-analyses were performed using Review Manager (RevMan) (https://training.cochrane.org/online-learning/core-software-cochrane-reviews). We assessed heterogeneity by inspecting the forest plots for overlapping CIs and outlying data, using the *I*^2^ statistic. When heterogeneity was detected, we used random-effects meta-analysis approach; otherwise, a fixed-effect approach was adopted. We tabulated adverse events and data that could not be meta-analysed. We carried out subgroup analyses to explore potential causes by stratifying the analyses by age – categorized as preschool children (PSAC), SAC or adults – background endemicity by type of schistosome species investigated– categorized as low, moderate and high (for *S. mansoni, S. japonicum, S. mekongi and S. intercalatum*), or low and high (for *S. haematobium*). We conducted a subsidiary, non-randomized comparison of AEs in children with AEs and adults for the same dose (mg/kg) to explore issues around dose applicability in children.

### Heterogeneity assessment

We assessed heterogeneity by inspecting the forest plots for overlapping confidence intervals and outlying data using the Chi^2^ test with a P value < 0.1 to indicate statistically significant heterogeneity; and using the *I*^2^ statistic. Although a *P*-value below 0.05 is generally considered to indicate statistical significance, we used a more sensitive threshold (DerSimonian and Laird 1986; Bossuyt et al. 2013) – i.e. a *P*-value below 0.10 – to indicate statistically significant heterogeneity. Where such significant heterogeneity is detected, we carried out subgroup analyses based on clinical and methodological differences. For data that could not be pooled quantitatively and where heterogeneity existed between studies, we grouped studies based on similarities.

### Summary of findings and grading the evidence

We used GRADE (Alonso-Coello et al. 2016; Santesso et al. 2020) to grade the evidence by looking at 5 main factors that might decrease quality of evidence, 1) study limitations, 2) inconsistency of results, 3) indirectness of evidence, 4) imprecision, and 5) publication bias. We also looked at factors that might increase quality of evidence including, 1) large magnitude of effect, 2) plausible confounding, which would reduce a demonstrated effect, and 3) dose-response gradient (http://www.gradeworkinggroup.org/).

## Results

### Description of studies

The search retrieved 3202 studies; 3047 studies from electronic databases and 155 studies from other sources including grey literature and contacts with experts. Seventy-three (73) studies were excluded because of duplication and 2931 were not relevant to the topic. A total of 198 full text studies were obtained of which 134 met the pre-specified eligibility criteria (Fig 1).

**Fig 1.**
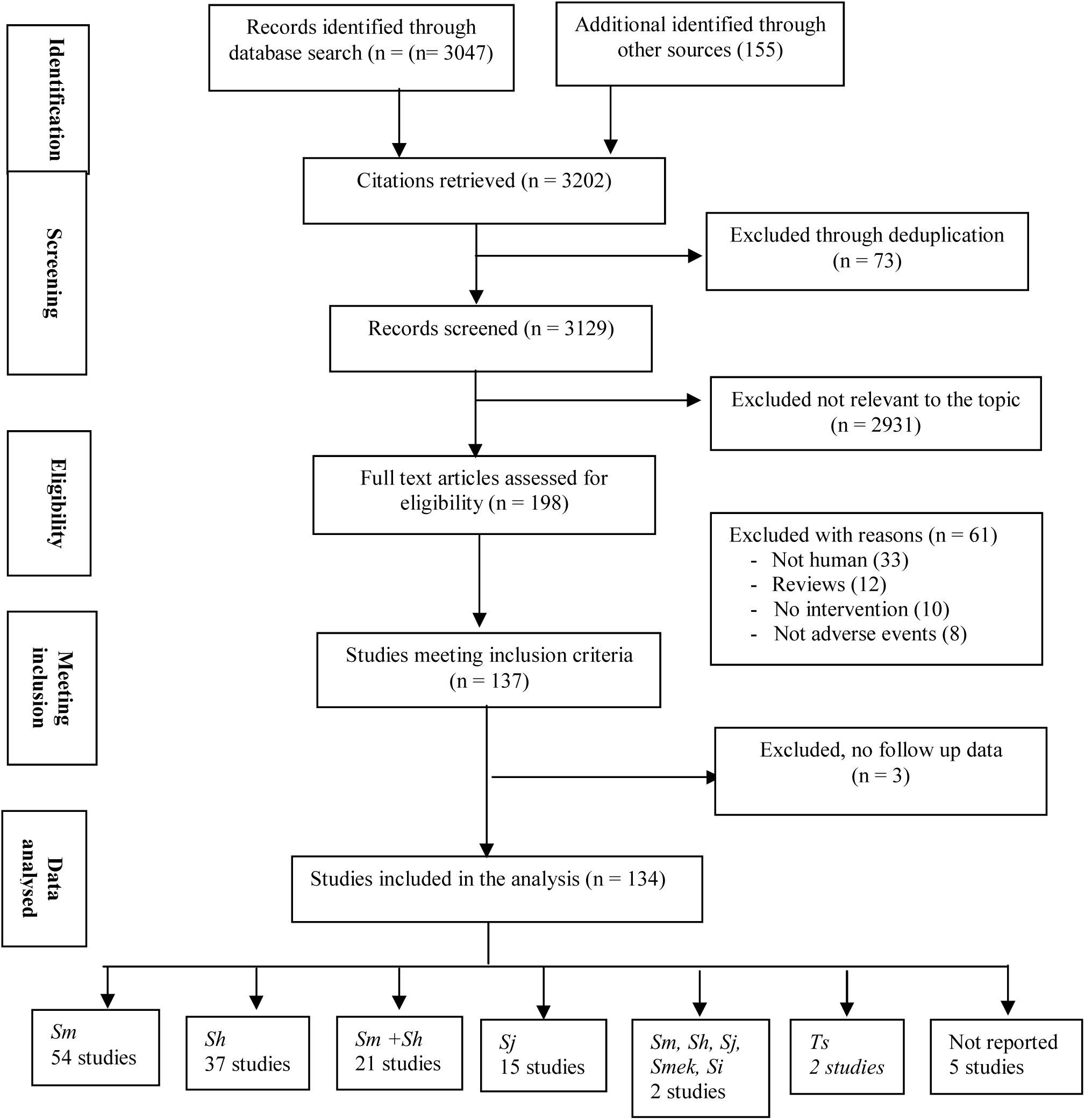
Flow diagram of studies retrieved from relevant electronic databases and other sources. Sm=S*. mansoni*, Sh=*S. haematobium*, Sj=*S. japonicum*, Smek=*S. mekongi*, Si=*S. intercalatum*, Ts=*Taenia solium*. Five studies did not report the species investigated.

Ninety-four of the 134 studies (70.1%) were conducted in Africa; 17 studies (12.7%) in Asia of which eight were conducted in Philippines (Belizario et al. 2007; Olds et al. 1999c; Olliaro et al. 2011c; Olveda et al. 2016; Santos et al. 1979; Santos et al. 1984; Watt 1986; Watt et al. 1988), seven in China (Fu-Yuan et al. 1984; Hou et al. 2008; Minggang et al. 1983; Minggang et al. 1985; Ming-he 1991; Olds et al. 1999a; Qian 2016), one in Japan (Ishizaki 1979) and one in Thailand (Na-bangchang et al. 2006); 15 studies (11.2%) were conducted in the Americas, 14 in Brazil (Branchini et al. 1982; Carvalho et al. 1984; Coutinho et al. 1983; Da Cuhna et al. 1987; da Cunha and Pedrosa 1986; da Silva et al. 1981; da Silva et al. 1986; de Quieroz et al. 2010; Ferrari et al. 2003; Katz et al. 1979; Katz et al. 1981; Katz et al. 1982; Katz et al. 1983; Olliaro et al. 2011a) and one in the USA (Nash et al. 1982); four studies (3.0%) in the Middle East, two in Saudi Arabia (Al-Aska et al. 1990; Azher et al. 1990), one in Israel (Cherit et al. 2014) and one in Yemen (Groning et al. 1985); and three studies (2.2%) in Europe, one each France, Germany and Spain (Bagheri et al 2004; Leopold et al. 1978; Bada et al. 1988) (Table 1).

**Table 1.**
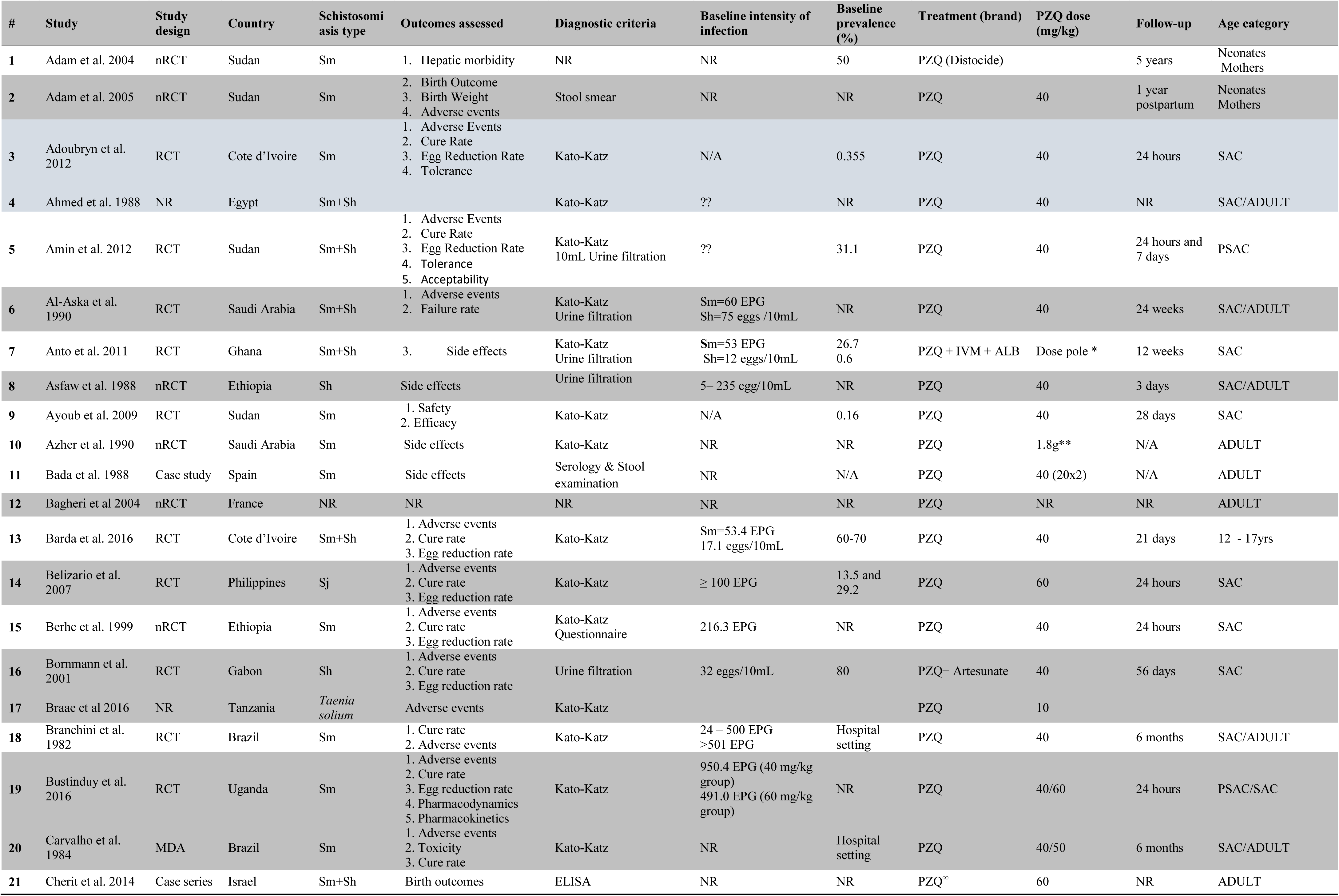
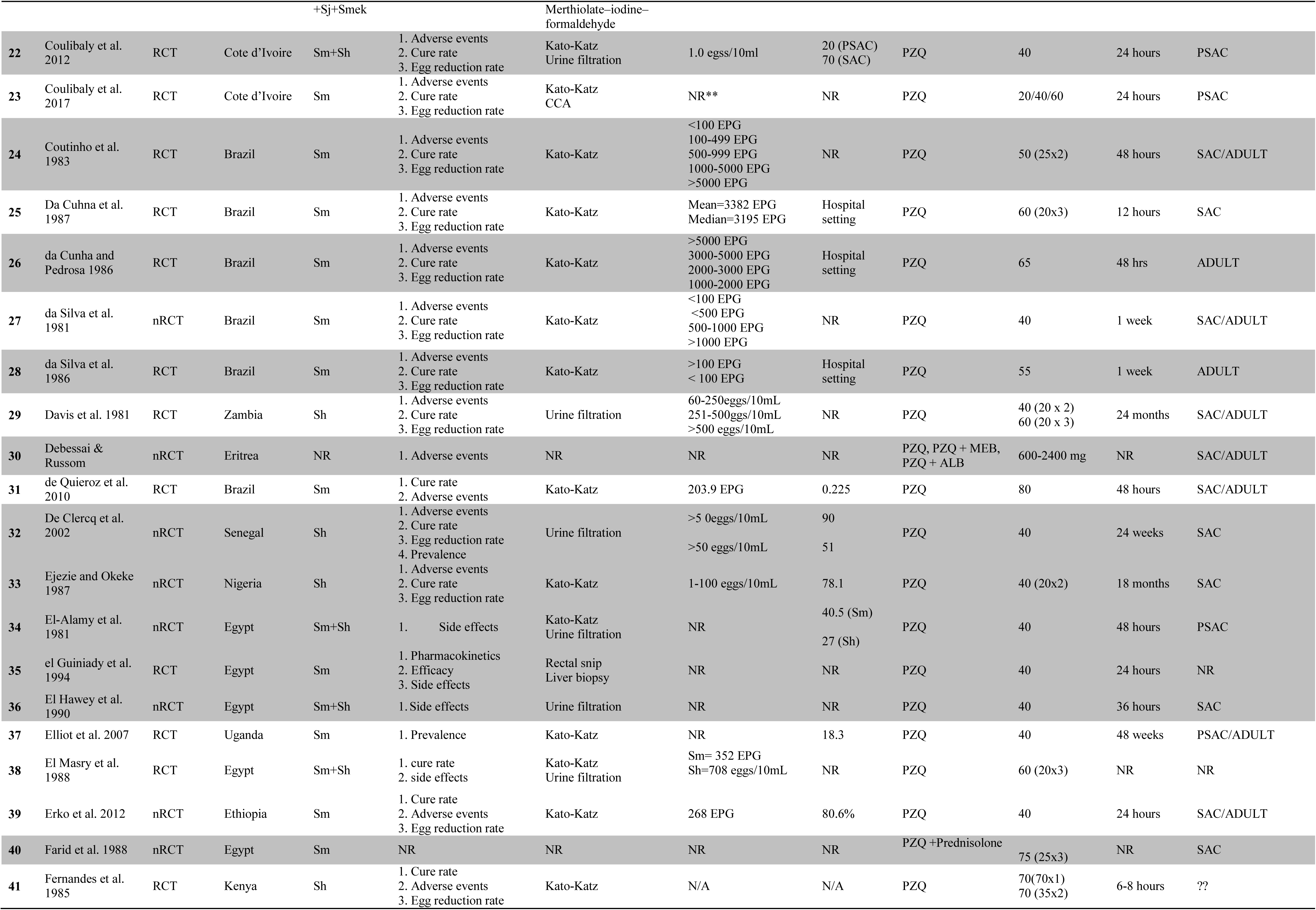
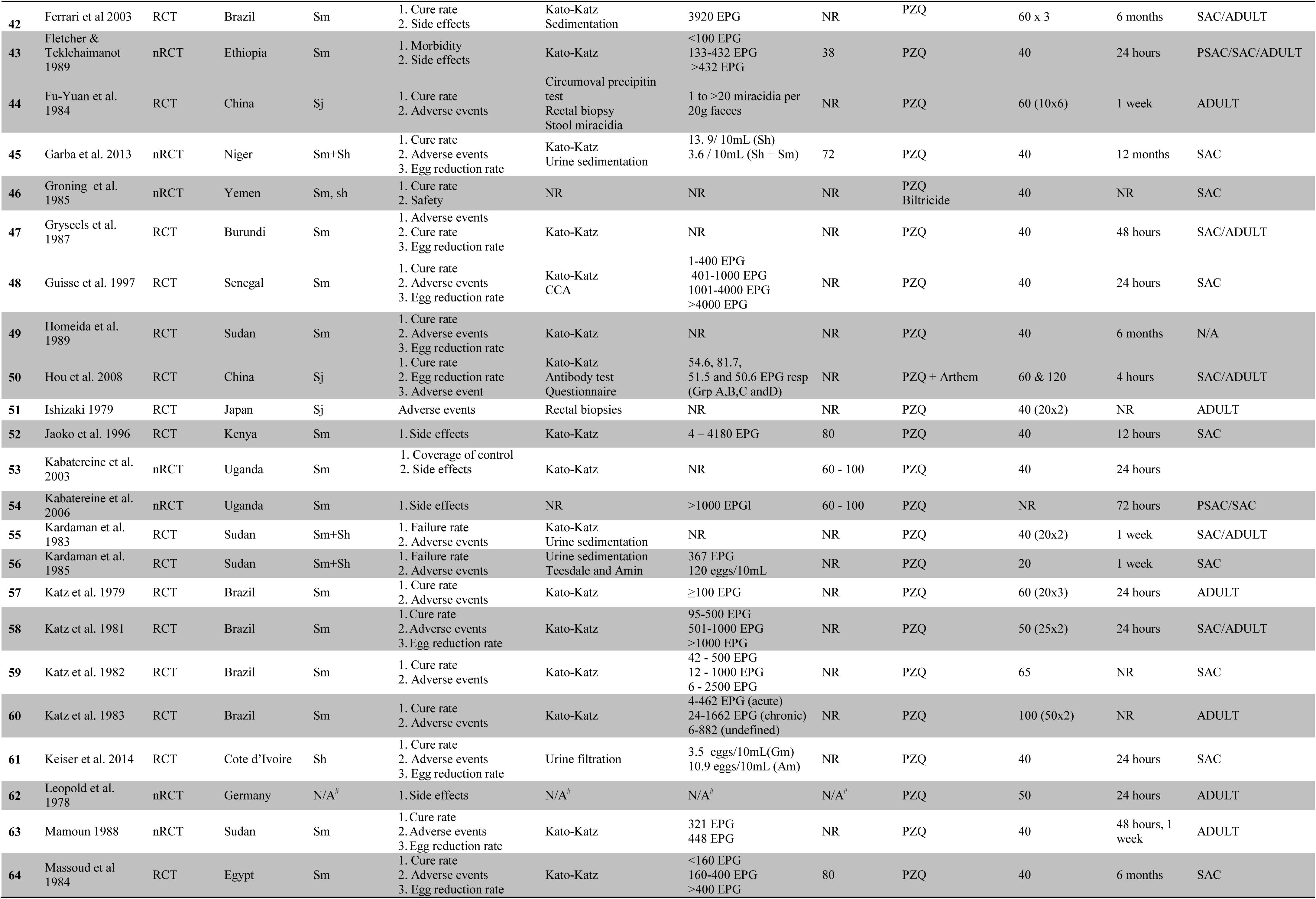
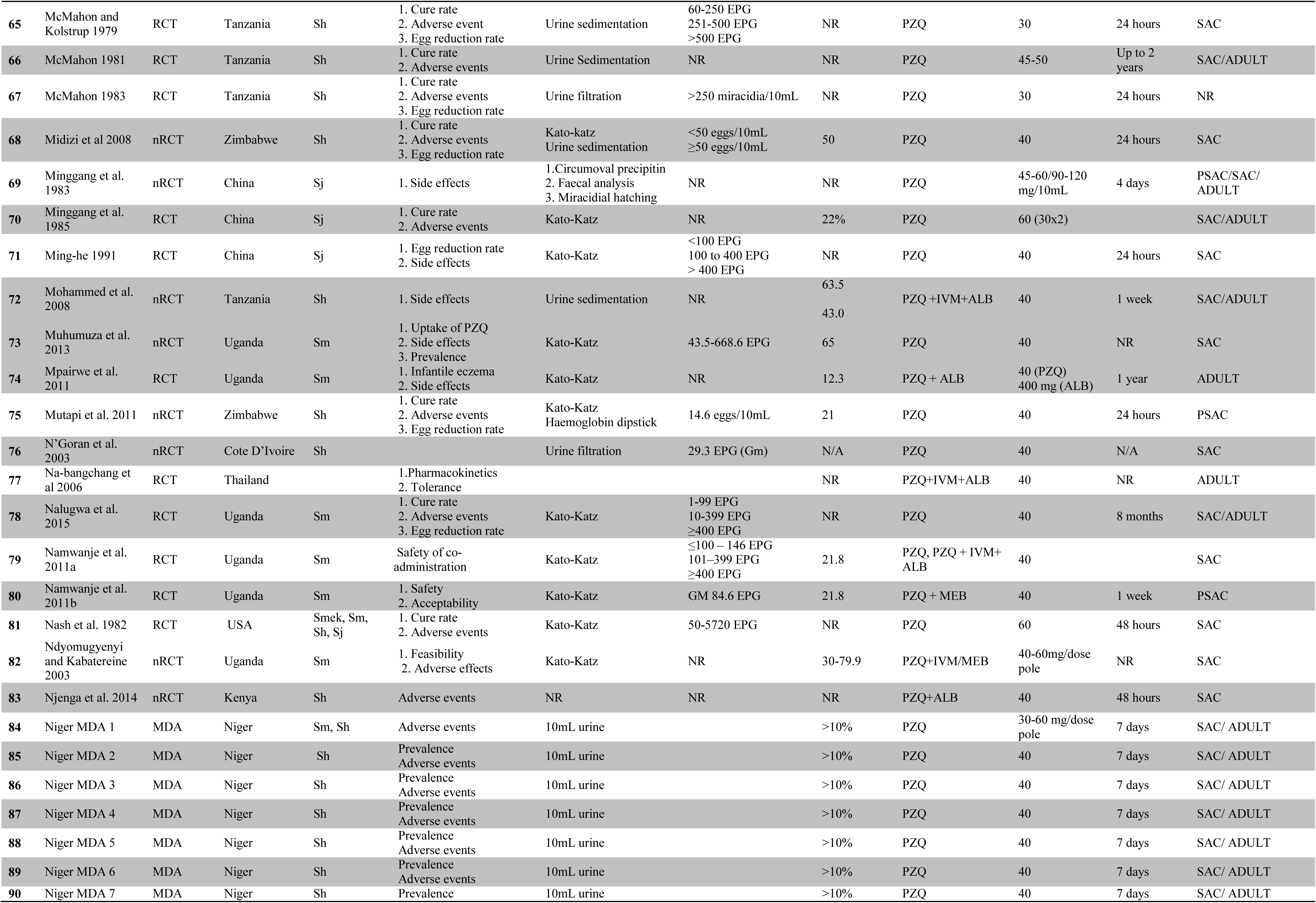
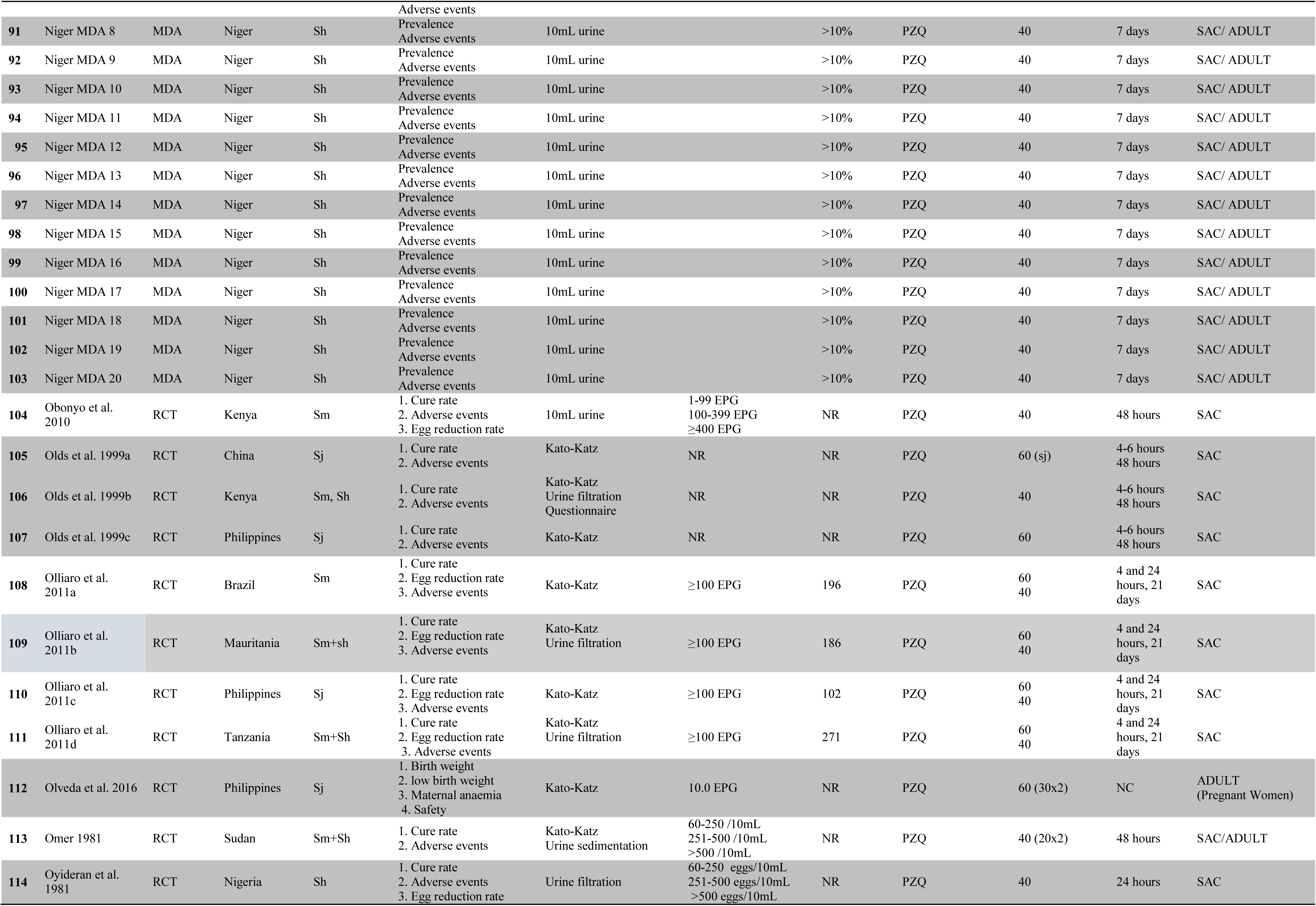
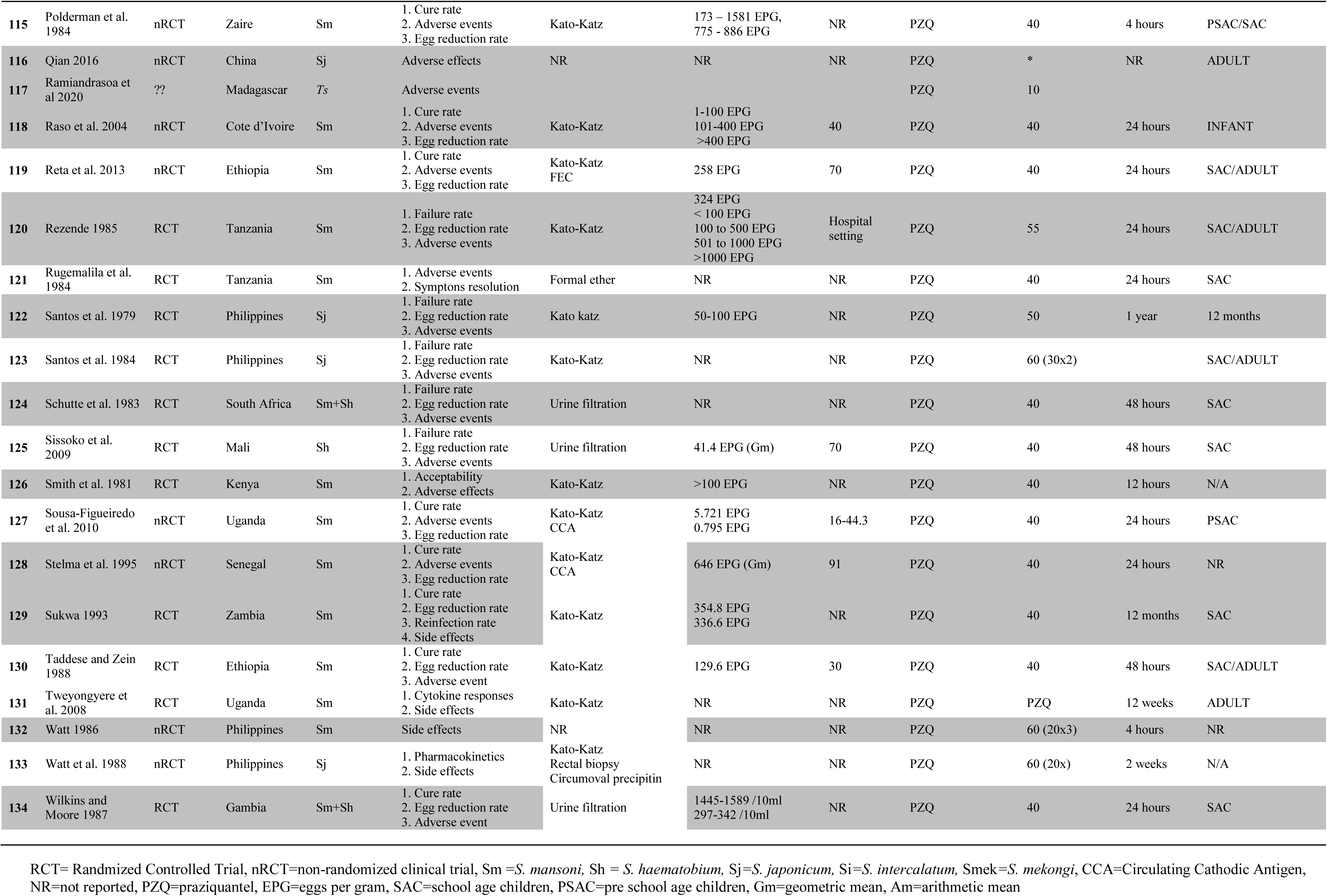
Characteristics of the studies included in the systematic review and meta-analysis

Olds et al. 1999 conducted a multi-country trial involving China (*S. japonicum*), Kenya (*S. manoni* and *S. haematobium*) and the Philippines (*S. japonicum*) which we included as separate studies (Olds et al. 1999a, Olds et al. 1999b, Olds et al. 1999c). The study by Olliaro et al. 2011 was a multi-country trial conducted in Brazil (*S. mansoni*), Mauritania (*S. manoni* and *S. haematobium*), Philippines (*S. japonicum*) and Tanzania (*S. manoni* and *S. haematobium*). We included data from each country as a separate study (Olliaro et al. 2011a, Olliaro et al. 2011b, Olliaro et al. 2011c, Olliaro et al. 2011d). For the country-wide MDA delivered across 20 districts of several villages, we included data from each district in the meta-analysis as a separate study (Niger MDA 1; Niger MDA 2; Niger MDA 3; Niger MDA 4; Niger MDA 5; Niger MDA 6; Niger MDA 7; Niger MDA 8; Niger MDA 9; Niger MDA 10; Niger MDA 11; Niger MDA 12; Niger MDA 13; Niger MDA 14; Niger MDA 15; Niger MDA 16; Niger MDA 17; Niger MDA 18; Niger MDA 19; Niger MDA 20).

Of the 134 studies, 71 (53.0%) were described by the authors as RCTs, 37 studies (27.6%) were non-randomized clinical trials, 21 (15.7%) were MDA and one case series (Cherit et al. 2014) and one case study (Bada et al. 1988). The design of three studies (Ahmed et al. 1988; Braae et al. 2016; Ramiandrasoa et al. 2020) was not stated by the authors but data were extracted and reported as unclear study design. Twenty-four (24) of the RCTs had multiple arms, and we analyzed according to arms in order to preserve the randomization code. In some of the non-randomized trials included in this systematic review and meta-analysis, study populations were grouped according to criteria specified by the primary investigators, for example, by intensity of infection, adverse events etc. De Clercq et al. 2002 assessed the efficacy of artesunate (an antimalarial drug) in treating *S. haematobium* and PZQ; we extracted data from the PZQ arm.

*Schistosoma mansoni* was the common schistosome species investigated followed by *S. haematobium* and *S. japonicum* in that order. Fifty-four of the studies were conducted in *S. mansoni* endemic settings and investigated adverse events associated with PZQ for treating for *S. mansoni* infection alone, 37 studies conducted in *S. haematobium* settings investigated adverse events associated with PZQ treatment for only *S. haematobium*, 21 studies involved both *S. mansoni* and *S. haematobium* infections, 15 studies investigated incidence of AEs events with *S. japonicum* alone, two studies investigated assess events in all five schistosome spp. commonly infecting humans (*S. mansoni, S. haematobium, S. japonicum, S. intercalatum* and *S. mekongi*), no study assessed *S. intercalatum* or *S. mekongi* alone and two studies assessed event rates in Taeniasis. No study assessed incidence of adverse events between those with and without schistosome infection receiving PZQ.

Twenty studies (14.9%) compared events rate between SAC and adults and 11 studies assessed mixed infection with *S. mansoni* and *S. haematobium*. Eight studies, four RCTs (Bustindy et al. .2020; Twenonyere et al. 2009; Elliot et al. 2007; Olveda et al. 2016) and four nonRCTs (Chetrit et al. 2014; Adam et al. 2004; Adam et al. 2005; Qian et al. 2016) investigated PZQ administration in pregnant women and five studies (Anto et al., 2011; Sissoko et al., 2009; Fu-yuan et al., 1984; Leopold et al., 1978; Rezende, 1985) focused on PZQ-related visual AES. Twenty-seven studies (20.1%) used dose schedules higher than 40 mg/kg and eight studies did not report the dose used. The rest of the studies used 40 mg/kg of PZQ either as a single dose or divided doses. With regard to follow up after treatment, only three studies (2.2%) collected AEs events data one year or more, the rest assessed incidence of events few hours, days or weeks after treatment. One hundred and thirteen studies administered PZQ alone (84.3%), and the common drugs co-administered with PZQ were Mebendazole, Albendazole, Ivermectin and the Artemisinin compounds (ACTs). All the included studies reported an element of safety (AEs) following treatment with PZQ.

### Commonly reported PZQ AEs

The commonly reported AEs have been presented (see Table 2). Overall, 75 studies involving 33550 participants that investigated PZQ reported incidence of abdominal pain as the leading AE experienced by persons treated with PZQ. This was followed by headache (68 studies, n = 25986 participants), vomiting (66 studies), dizziness (61 studies, n = 20716 participants), diarrhoea (59 studies, n > 18595), nausea (56 studies, n = 19009 participants), fever (36 studies, n = 23096 participants), itching (34 studies, 16247 participants) and somnolence (23 studies, n = 12805) in that order (Table 2). Two hundred and eighty-nine thousand twenty-four (289024) adverse events were recorded across the 134 studies included in this systematic review. Praziquantel administered alone accounted for 188809 of the adverse events whereas PZQ in combination with other drugs produced 99912 events. All the studies reported mild-to-moderate adverse events that resolved on their own. Some studies reported severe adverse events but all resolved on their own without further intervention. A total of 40 serious SAEs were reported among the general population. Some studies did not define or describe the nature of the AEs.

**Table 2.** Commonly reported adverse experienced by persons who received PZQ treatment alone or in combination with other drugs (ALB, MEB, IVM) arranged according to number of studies

| Adverse event | No. of studies | No of studies by Species |  |  |  |  | Total adverse events | Adverse events by treatment |  | Serious AE's |
| --- | --- | --- | --- | --- | --- | --- | --- | --- | --- | --- |
|  |  | Sm | Sh | Sm+Sh | Sj | S. mek |  | PQZ alone | PQZ + other* |  |
| Abdominal pain | 75 | 43 | 12 | 10 | 8 | 0 | 33550 | 33225 | 325 | 0 |
| Headache | 68 | 36 | 14 | 4 | 8 | 1 | 25986 | 21414 | 4572 | 0 |
| Vomiting | 66 | 36 | 12 | 10 | 5 |  | " | " | ?? | 1 |
| Dizziness | 61 | 30 | 19 | 7 | 4 | 0 | 20716 | 14729 | 5987 | 0 |
| Diarrhoea | 59 | 33 | 13 | 9 | 2 | 1 | 18595 | 11659 | 6936 | 2 |
| Nausea | 56 | 32 | 14 | 6 | 3 | 0 | 19009 | 11653 | 7356 | 9 |
| Fever | 36 | 20 | 8 | 5 | 1 | 1 | 23096 | 10537 | 12559 | 0 |
| Itching | 34 | 19 | 8 | 2 | 3 | 1 | 16247 | 6179 | 10068 | 0 |
| Somnolence | 23 | 11 | 6 | 1 | 2 | 1 | 12805 | 6820 | 5985 | 0 |
| Rash | 23 | 11 | 4 | 4 | 2 |  | 43500 | 31493 | 12007 | 25 |
| Fatigue | 18 | 8 | 2 | 3 | 3 | 0 | 10662 | 5108 | 5545 | 0 |
| Anorexia | 17 | 7 | 5 | 1 | 3 | 1 | 10580 | 2860 | 7720 | 5 |
| Urticaria | 13 | 12 | 0 | 0 | 0 | 1 | 2229 | 1365 | 771 | 0 |
| Blood in stool | 9 | 5 | 0 | 1 | 3 | 0 | 14235 | 7245 | 6990 | 0 |
| Myalgia | 9 | 4 | 2 | 1 | 0 | 1 | 13386 | 1435 | 11951 | 0 |
| vertigo | 8 | 3 | 3 | 0 | 2 | 0 | 1652 | 1263 | 389 | 0 |
| Asthenia | 8 | 7 | 1 | 0 | 0 | 0 | 2201 | 1665 | 536 | 0 |
| Malaise | 8 | 5 | 1 | 1 | 0 | 1 | 765 | 736 | 29 | 0 |
| Bitter taste | 7 | 7 | 0 | 0 | 0 | 0 | 852 | 852 | 0 | 0 |
| Cough | 7 | 4 | 1 | 2 | 0 | 0 | 4234 | 3847 | 186 | 0 |

\*co-administered PZQ with other drugs (ALB, MEL, IVM); AEs=serious adverse events, Sm = *S. mansoni*, Sh = *S. haematobium*, Sj =*S. japonicum*, Smek =*S. mekongi*

### Praziquantel versus placebo

An RCT is the only design that is able to ascertain the true experience of adverse events because it is able to control for confounding effects/events. Eight RCTs, four conducted in Africa (Borrmann et al. 2001; Jaoko et al. 1996; Olds et al. 1999; Tweyongyere et al. 2009) and the other four in the Americas, all conducted in Brazil (Branchini et al. 1982; Ferrari et al. 2003; Katz et al. 1979a; Katz et al. 1979b) assessed PZQ 40 mg/kg or higher doses and reported mild-to-moderate and transient AEs (Table 3). None reported any serious event. Olds et al. 1999 recorded 15% excess of mild to moderate AEs events with PZQ compared with placebo, and Borrmann et al. 2001 assessed PZQ, Artesunate and placebo and reported combined events across comparison groups (127 mild and 6 moderate events). No SAEs were recorded. The rest of the results have been presented in Table 3.

**Table 3.** Adverse events associated with PZQ in comparison with placebo in RCTs

| Study ID | Country | N | PZQ dose (mg/kg) | Adverse events | SAE |
| --- | --- | --- | --- | --- | --- |
| *S. mansoni* |  |  |  |  |  |
| Jaoko et al. 1996 | Kenya | 436 | 40x1 | Minor and transient adverse events.<br>PZQ versus placebo:<br>Dizziness: 36% vs 6%<br>Abdominal pain 35% vs 14 % | None reported |
| Katz et al. 1979a | Brazil | 55 | 40 (20x2)<br>60 (20x3) | Minor adverse events, did not differ between intervention and placebo groups<br>Adverse events were not reported separately for the different dose schedules. | None reported |
| Katz et al. 1979b | Brazil | 61 | 50x1<br>60 (20x3) | Adverse events were minor, did not differ between the two groups, but were not reported separately for the two dosing schedules. | None reported |
| Olds et al. 1999 | Kenya | 174 | 40x1 | PZQ versus placebo:<br>Abdominal pain: 80% vs 50%<br>Diarrhoea: 54% vs 25% | NR |
| Tweyongyere et al. 2009 | Uganda | 387 | 40x1 | Adverse events were minor and transient. The authors pooled adverse events together over the intervention and placebo groups. Event rates were not reported. | None reported |
| Branchini et al. 1982 | Brazil | 70 | 41.2-51.6 | Mild and transient AEs<br>PZQ versus placebo in that order:<br>Dizziness: 46.9% (placebo group not reported).<br>Abdominal pain: 24.5% vs 17.6% | None reported |
| Ferrari et al. 2003 | Brazil | 72 | 180 (60x3) | Mostly headache, dizziness, drowsiness and abdominal pain with no apparent differences between the arms. | None reported |
| *S. haematobium* |  |  |  |  |  |
| Borrmann et al. 2001 | Gabon | 300 | 40 | 127 mild and 6 moderate events.<br>Abdominal pain: (14%)<br>Headache: (12%) | None reported |
| Olds et al. 1999 | Kenya | 380 | 40<br>40+ALB | Adverse events mild to moderate.<br>PZQ: 15% (PZQ) vs 20% (PZQ + ALB) | None reported |

N= number of participants, SAE=serious adverse event; ALB=Albendazole;

### Divided doses of PZQ

Eighteen studies (Bada et al. 1988; Bustinduy et al 2020; Ejezie and Okeke 1987; Da Cunha et al 1983; Da Cunha et al 1987; El-Masry et al. 1988; Farid et al 1988; Fernandes et al. 1985; Fu-Yuan et al 1984; Ishizaki 1979; Kardaman et al 1983; Katz et al. 1979; Katz et al. 1981; Minggang et al 1985; Olveda et al. 2016; Omer et al 1981; Santos et al 1984; Watt et al. 1986; Watt et al. 1988) assessed various divided doses of PZQ across different endemic settings and schistosome types (Table 4). Fourteen of the studies were RCTs, two cohort studies, one case study and one did not report the type of study design. Various dose schedules were assessed including 40 mg/kg administered as a divided dose of 20 mg/kg x2, 50 mg/k (administered as 25mg/kg double doses), 60 m/kg (as a divided dose of 30 mg/kg or 20 mg/kg taken same day), 75 mg/kg (at 25 mg/kg x 3) and very high doses of 180 mg/kg in divided doses (Table 4). Most of the studies reported minor adverse events, predominantly headache, vomiting, abdominal pain, dizziness and nausea. Some studies reported severe (Bustinduy et al 2020; Da Cunha et al 1986; Olveda et al. 2016; Santos et al 1984; Watt et al. 1986) or serious SAEs (Bada et al. 1988; Minggang et al 1985; Olveda et al. 2016), with the highest incidence of AEs recorded in patients who received higher doses. Studies assessing PZQ 40 mg/kg and comparing with higher doses have been reported in Table 5 -supplementary table.

**Table 4.** Adverse events with divided doses of PZQ

| Study and country | Design | Species | PZQ dose | N | Adverse events | Serious adverse events |
| --- | --- | --- | --- | --- | --- | --- |
| Bada et al. 1988 Spain | Case study | Sm | 40 (20x2) | 1 | Patient experienced “tonic-clonic convulsive seizure” after treatment. | Seizure, but patient had a history of convulsion |
| Bustinduy et al 2020 Philippines | RCT | Sj | 60 (30x2) | 47 | Mostly fever, headache, malaise, abdominal pain, vomiting, bloody stools, rash, dizziness etc. which were mild to moderate and self-limiting. Only 2 severe events (headache and malaise). | None reported |
| Da Cunha et al 1986 Brazil | RCT | Sm | 50 (25x2) | 27 | 37% of patients had AEs with 12.5% classified as ‘severe’. General malaise (22.2%), heartburn (18.5%), dizziness (18.5%) and nausea (14.8%). One report of eosinophilic pneumonitis, Loeffler syndrome was diagnosed by X-Ray and laboratory assessment in a patient 5 days post-treatment | None reported |
| Da Cunha et al 1987 Brazil | RCT | Sm | 180 (30x2 daily for 3 days<br>180 (30x1) daily for 6 days<br>120 (30 x2) daily for 2 days<br>60 (30x2 in 1 day) | 79 | Higher proportions with adverse events but all were minor and transient events.<br>Dizziness: 65%, 15%, 45%, 15%<br>Nausea: 55%, 15, 20%, 20%<br>Some AEs were classified as severe<br>The highest incidence of AEs was recorded in patients who received 60 mg/kg daily for 3 days treatment, significantly higher than in those who were treated with 30 mg/kg daily for 6 days | None reported. |
| El-Masry et al. 1988 Egypt | RCT | Sm, Sh | 60 (20x3) | 204 | Epigastric pain was the most common AE (up to 66.6% of participants who received 20 mg twice daily for 3 days), headache, nausea, vomiting and fever up to 38°C were also common | None reported |
| Farid et al 1988 Egypt | Not stated | Sm | 75 (25x3) | 25 | The authors first managed the fever and toxemia in the patients before treating with PZQ to avoid serious AEs | None reported |
| Fernandes et al. 1985 Kenya | RCT |  | 70 (35x2) |  |  |  |
| Fu-Yuan et al 1984 China | RCT | Sj | 60 (10x6) | 716 | Of the 476 with chronic schistosomiasis, 299 (62.8%) had AEs; dizziness (112), headache (73), abdominal pain (48), blurring of vision (11) | None reported |
| Ishizaki 1979 Japan | RCT |  | 40 (20x2);<br>60 (20x3) | 51 | 14% in Grp1, 12.5% in Grp2 and 58% in Grp3 had AEs such as drowsiness, constipation, headache, etc. AEs disappeared within 24hrs<br>AEs higher in patients treated with 20x3mg/kg dose AEs were also high in placebo group as well. | None reported |
| Kardaman 1983 Sudan | RCT | Sm, Sh | 40 (20x2) | 171 | 45% had complaints; abdominal pains (13%), vomiting (7.5%), diarrhoea (7.5%), urticaria (11%), headache (7.5%), fever (7.5%). All events resolved within 24 hours | None reported |
| Katz et al. 1979 Brazil | RCT | Sm | 40 (20x2)<br>60 (20x3) | 50 | In the double-blind trial, 88% of patients treated with 20x2 dose had AEs; 62% of patients treated with 30x2 had AEs. 70% of patients treated with 30x2 PZQ had AEs | None reported |
| Katz et al. 1981 Brazil | RCT | Sm | 50 (25x2) | 46 | 82.6% experienced AEs. AEs were chiefly abdominal distress, giddiness and headache. AEs were slightly higher in SAC than adults (83% and 81%) | None reported |
| Minggang et al 1985 China | RCT | Sj | 60 (30x2)<br>70(<15yrs)<br>90-120 (over six days) | 25693 | .Majority of patients who had SAEs had pre-existing conditions. Events included:<br>Neuropsychiatric events (n=39)<br>Cardiovascular events (n=37)<br>Hepatic changes (n=4)<br>Dermatological events (n=18)<br>Delayed reactions leading to fatigue work (n=29) | 122 (0.47%) relatively serious AEs |
| Olveda et al.<br>2016<br>Philippines | RCT | Sj | 60 (30x2) | 186 | 5 cases of severe AEs were recorded 24hrs post-treatment | 18 events in mothers and 27 events in the foetuses. AEs not likely to be associated with PZQ treatment |
| Omer et al<br>1981 Sudan | RCT | Sm, Sh | 40 (20x2) | 52 | Adverse events were transient and required no additional intervention. | None reported |
| Santos et al<br>1984<br>Philippines | RCT | Sj | 60 (30x2) | 6134 | 67.8% patients had AEs of which 85% were mild, 13.7% moderate and 1.2% severe. AEs were mostly abdominal pain, dizziness, headache, bloody stools, vomiting, fever, etc. The severe AEs were mainly abdominal pain and started within 1 hour after treatment | None reported |
| Watt et al.<br>1986<br>Philippine s | Cohort | Sj | 60 (20x3) | 20 | 4 patients had abdominal pain which progressed to bloody diarrhoea. The affected patients had occult blood in pre-treatment stool and severe liver disease, considered as risk factors for developing post-OZQ bloody diarrhoea | None reported |
| Watt et al.<br>1988<br>Philippines | Cohort | Sj | 60 (20x3) | 30 | All patients experienced mild transient side effects. Drowsiness was the most common followed by headache and then abdominal pain. AEs were found to be associated with elevated plasma concentrations of PZQ | None reported |
| **Sh:** *Schistosoma haematobium* **Sm:** *Schistosoma mansoni* **Sj:** *Schistosoma japonicum* **Si:** *Schistosoma intercalatum*<br>**Sm:** *Schistosoma mekongi* , N=number of participants |  |  |  |  |  |  |

### Diarrhoea

The praziquantel 40 mg/kg group experienced a 2% reduced Praziquantel 40mg/kg did not show an increased odd of an individual developing diarrhea compared to Praziquantel 60mg/kg. Belizario 2007 reported relatively high adverse events following PZQ administration but also suggested that, this could have also been due to the parasitic infection. Coulibaly 2007 also did report no difference in the incidence of adverse events in all treatment groups of either school-aged children or pre-school-aged children.

**Fig 5.**
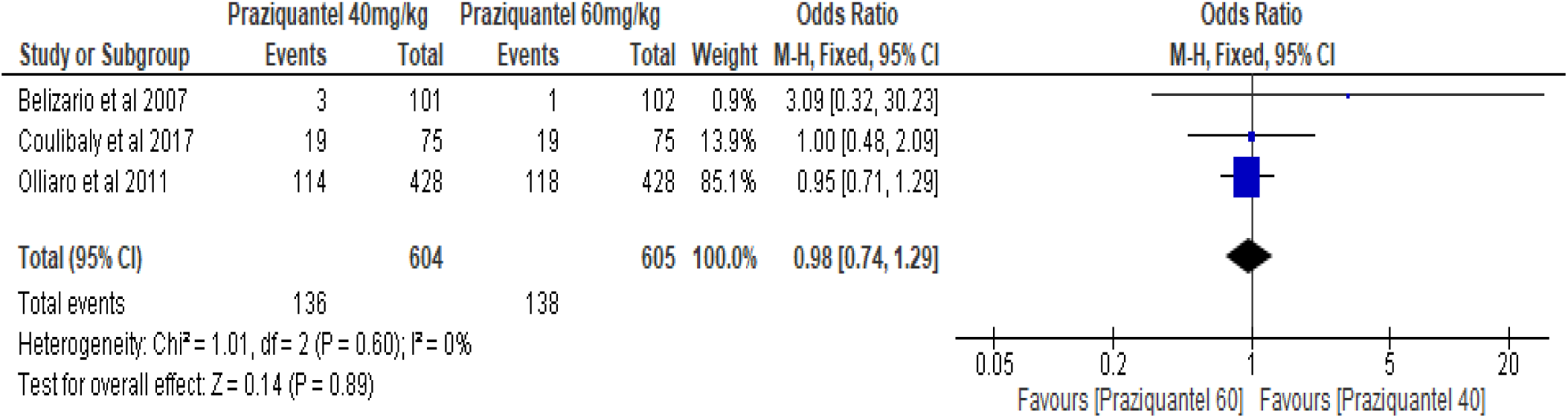
Forest plot showing the odds of diarrhea between praziquantel 40mg/kg vs Praziquantel 60mg/kg

#### Vomiting

The results indicate that individuals receiving Praziquantel 40mg/kg are 57% less chance of vomiting compared to individuals receiving Praziquantel 60 mg/kg (OR= 0.43; CI= 0.43 - 1.29). In two of the three studies (Belizario et al 2007; Olliaro et al 2011), adverse events were assessed as probably or most probably caused by PZQ chemotherapy and the cumulative prevalence of adverse events was found to be significantly different between those that received 40mg/kg and 60mg/kg. Characterizing the incidence of adverse effects by time elapsed post-treatment, Olliaro et al 2011 reported that 2% and 1% in the 40mg/kg and 60mg/kg treatment groups respectively, reported severe vomiting at 4 hours post-treatment. At 24 hours post-treatment, 14% and 33% reported vomiting with a moderate intensity.

**Fig 6.**
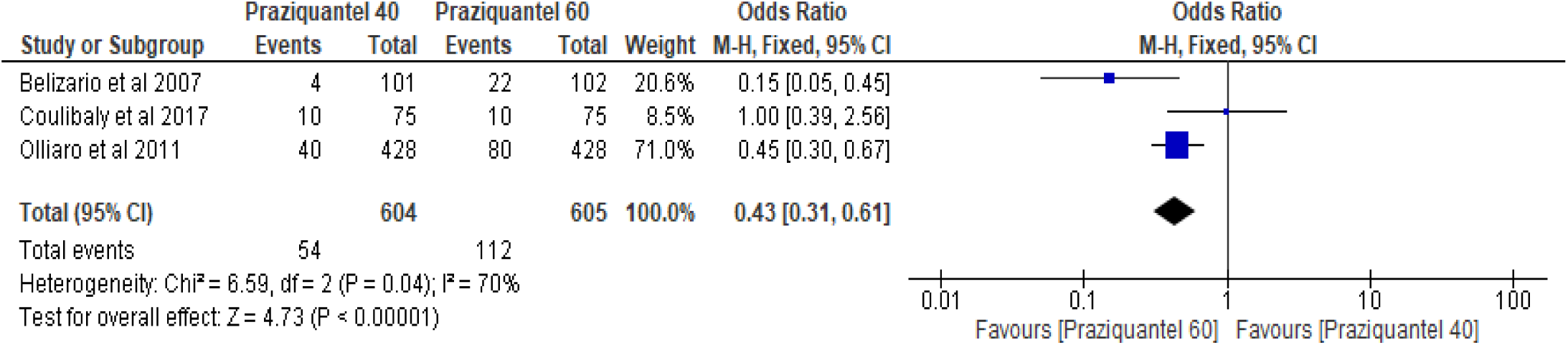
Forrest plot showing the odds of vomiting between Praziquantel 40mg/kg vs Praziquantel 60 mg/kg

### Headache

The meta-analysis suggested that there is no significant difference between the odds of having a headache following treatment with PZQ either at a dose of 40 mg/kg or 60 mg/kg (OR=0.99, CI= 0.74 - 1.33). Belizario et al, 2007 recorded the same number of reports of headache in both groups of the study (31 events in each group). In the two other studies that compared adverse events in 40mg/kg and 60mg/kg, the number of reports of headache following treatment with PZQ did not differ significantly (6 and 11 in Coulibaly et al 2017, 73 and 69 in Olliaro et al 2011).

**Fig 7.**
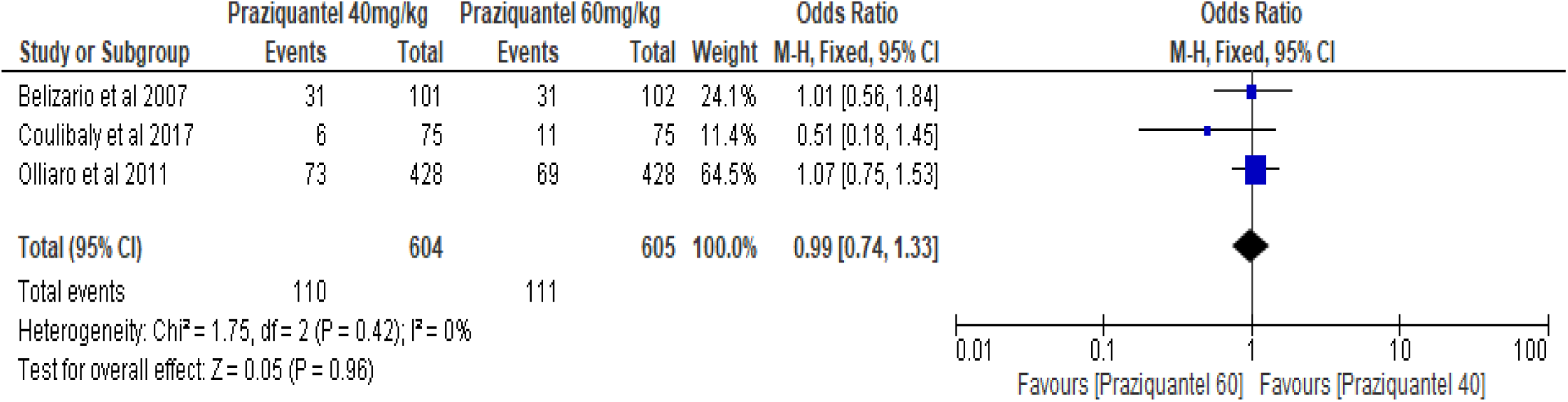
Forest pot showing the odds of experiencing headache between Praziquantel 40mg/kg vs Praziquantel 60mg/kg

#### Praziquantel 40 mg/kg vs Placebo

Two studies made this comparison (Coulibaly 2017; Schutte et al 1983). Coulibaly et al 2017 compared safety and efficacy of PZQ at 40 and 60 mg/kg in PSAC and SAC. They reported that in both PSAC and SAC treatment groups, diarrhea was one of the commonly reported adverse events when assessed at 3 - 4 hours post-treatment. Schutte et al 1983 studied the efficacy of PZQ administered at 40 mg/kg in children aged between 7 and 17 years. In their assessment of adverse events immediately and 48 hours post-treatment, they reported that adverse events were common in the treatment groups compared to the placebo. From the meta-analysis, the is 60% increased odds of diarrhea in the PZQ treatment group compared to the placebo (OR= 1.60, CI= 0.29 - 8.66).

**Fig 8.**
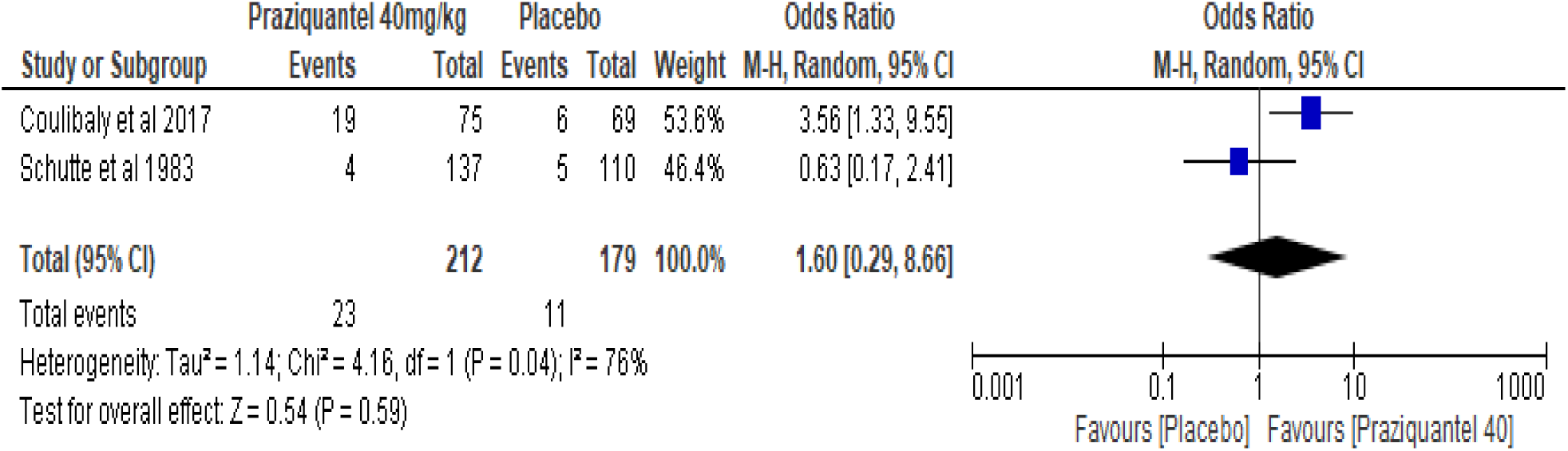
Forest plot showing the odds of diarrhoea between praziquantel 40mg/kg and placebo

### *Schistosom mansoni* and *S. haematobium* mixed infection

Thirteen studies (Ahmed et al. 1988; Al-Aska et al. 1990; Anto et al. 2011; Barda et al. 2016; Coulibaly et al. 2012; El-Hawey et al. 1990; El-Masry et al. 1988; El-Alamy et al. 1981; Garba et al. 2013; Groning 1985; Kardaman et al. 1983; Omer et al. 1981; Schutte et al. 1983) involving eight RCTs (Al-Aska et al. 1990; Anto et al. 2011; Barda et al. 2016; Coulibaly et al. 2012; El-Masry et al. 1988; Kardaman et al. 1983; Omer et al. 1981; Schutte et al. 1983), three clinical trials (El-Hawey et al. 1990; El-Alamy et al. 1981; Garba et al. 2013, one MDA (Groning 1985) investigated adverse events of PZQ in mixed infection with *S. amsoni* and *S. haematobium*. One study (Ahmed et al. 1988) did not state the study design. The studies pooled did not record any SAEs; all events were mild-to-moderate and transient (Table -supplementary file).

### Adverse events between SAC and Adults

Twenty studies involving over one million (1,100,992) participants assessed adverse events associated with PZQ 40 mg/kg and reported abdominal pain, diarhoea, headache, nausea, vomiting, dizziness, allergy, anorexia and fever as the most frequently experienced adverse events, and arranged in a decreasing incidence (Table 7). Adverse events were mostly mild-to-moderate and transient, resolving few hours after treatment. There were reports of some severe events but these were non-life threating and did not require further intervention. No serious event was recorded. Risk ratios, CIs and p-values obtained from the meta-analyses have been reported (Table 7). Children experienced higher frequencies of all the adverse events, and the risk ratios were statistically significance over events in adults, except swelling where there was not apparent difference in incidence between SAC and adults. Overall, 23,939 incidence of headache (20004 out of 694530 SAC and 3937 out of 406462 adults treated with 40 mg/kg PZQ) was recorded. The pooled risk ratio (RR) showed a three times increased risk of headache following PZQ treatment in SAC than adults (RR 3.07, 95% CI 2.32 to 4.06, n=1,100,992, twenty studies, I^2^=98%, p<0.00001) (Fig 10). Homogeneity between studies (measured by the I^2^) was very high (98%). Dizziness was the next frequently reported adverse event and the incidence was much higher in SAC than adults (RR 1.80, 95% 1.36 to 2.37, n=1,100,992, twenty studies, I^2^=98%; p=0.0001) (Fig 11). Vomiting formed about 14% of all adverse events reported with higher risk (2.5 times) of experiencing an event in SAC than adults (RR 2.43, 95% CI 1.87 to 3.14, I^2^=98%; p<0.00001) (Fig 12). The risk of experiencing an adverse event was higher in SAC than adults for abdominal pain (RR 3.97, 95% CI 3.09 to 5.10, I^2^ = 96%, p< 0.00001.), nausea (RR 1.67, 95% CI 1.32 to 2.12, I^2^=97%, p <0.0001), general discomfort (RR 1.32, 95% CI 1.03 to 1.68, I^2^=97%, 0.00001), fever (RR 4.78, 95% CI 3.04 to 7.52, twenty studies, I^2^=98%, p < 0.00001), diarrhoea (RR 1.41, 95% CI 1.12 to 1.78, twenty studies, I^2^=92%, p < 0.00001), Itching (RR 2.42, 95% CI 1.58 to 3.70, twenty studies, I^2^=93%, 0.0001), breathing difficulty (RR 2.46, 95% CI 1.41 to 4.29, twenty studies, I^2^=92%, 0.002) and swelling (RR 1.15, 95% CI 0.98 to 2.31, twenty studies, I^2^=85, 0.06). One study (MDA15) did not have any event in the adult arm for both headache and dizziness hence the wide confidence intervals (Fig 10 and Fig 11). See supplementary documents for figures 12 to 20.

**Table 7.** Incidence of adverse events with the standard 40 mg/kg single dose PZQ pooled from 20 studies involving over one million treated persons

| Adverse event | Adverse events |  | Total | Percent (%) | Risk Ratio | Confidence interval (CI) |  | p-value |
| --- | --- | --- | --- | --- | --- | --- | --- | --- |
|  | SAC | Adults | SAC + Adults |  |  | Lower | Upper |  |
| Headache | 20004 | 3937 | 23939 | 19.1 | 3.07 | 2.32 | 4.06 | <0.00001 |
| Dizziness | 13597 | 5281 | 18878 | 15.1 | 1.80 | 1.36 | 2.37 | <0.0001 |
| Vomiting | 13986 | 4074 | 18060 | 14.4 | 2.43 | 1.87 | 3.14 | <0.00001 |
| Abdominal pain | 12572 | 2152 | 14724 | 11.4 | 3.97 | 3.09 | 5.10 | <0.00001 |
| Nausea | 10248 | 3536 | 13784 | 11.0 | 1.67 | 1.32 | 2.12 | <0.0001 |
| General discomfort | 8785 | 3902 | 12687 | 10.1 | 1.32 | 1.03 | 1.68 | <0.00001 |
| Fever | 9168 | 1771 | 10939 | 8.7 | 4.78 | 3.04 | 7.52 | <0.00001 |
| Diarrhoea | 3846 | 1771 | 5617 | 4.5 | 1.41 | 1.12 | 1.78 | <0.00001 |
| Itching | 1873 | 755 | 2628 | 2.1 | 2.42 | 1.58 | 3.70 | <0.0001 |
| Breathing difficulty | 1181 | 405 | 1586 | 1.3 | 2.46 | 1.41 | 4.29 | 0.002 |
| Swelling | 741 | 432 | 1173 | 0.9 | 1.15 | 0.98 | 2.31 | 0.06 |

**Fig 10.**
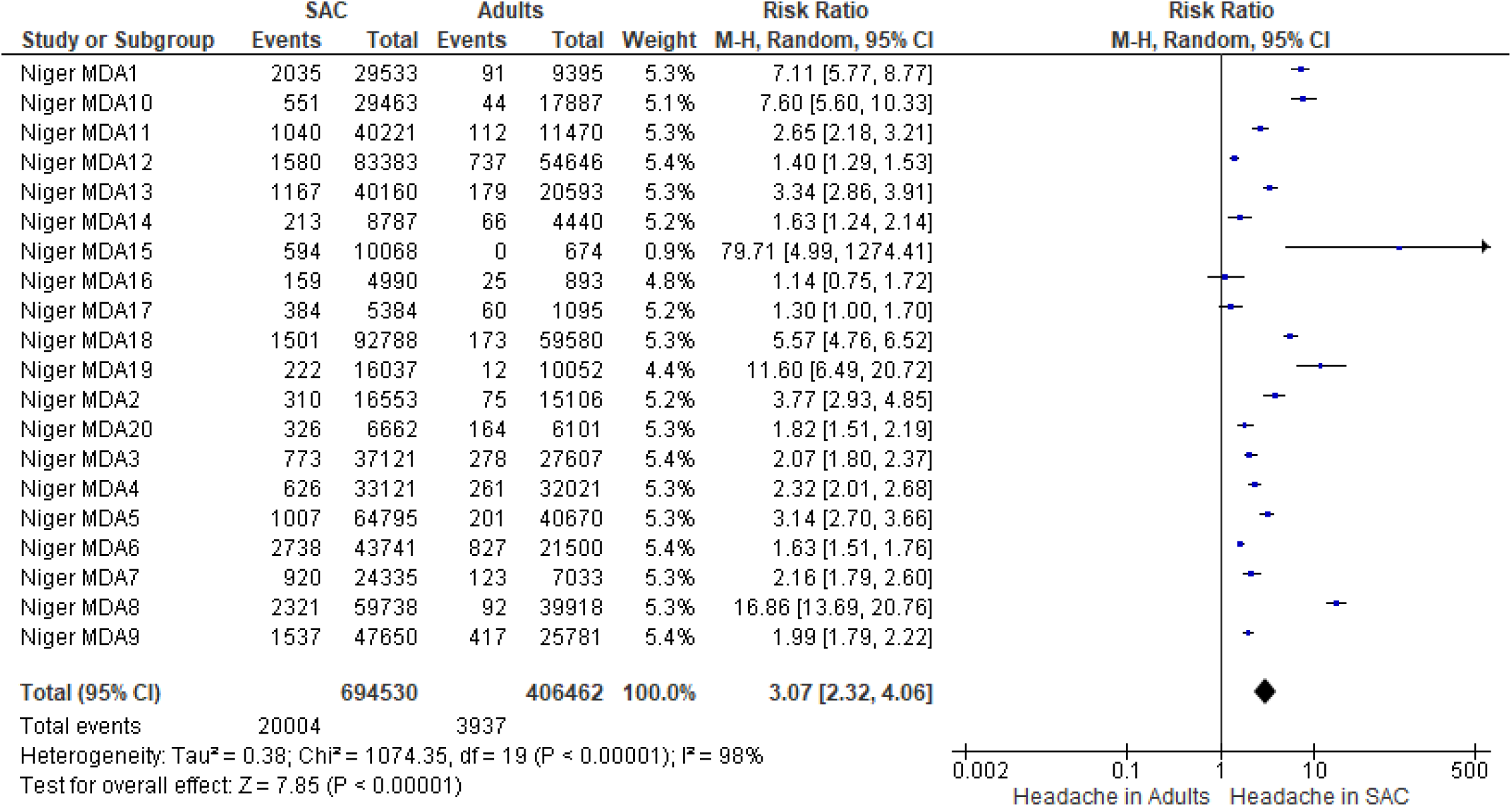
Headache associated with PZQ (40 mg/kg single dose) between school age children and adults in preventive chemotherapy programmes

**Fig 11.**
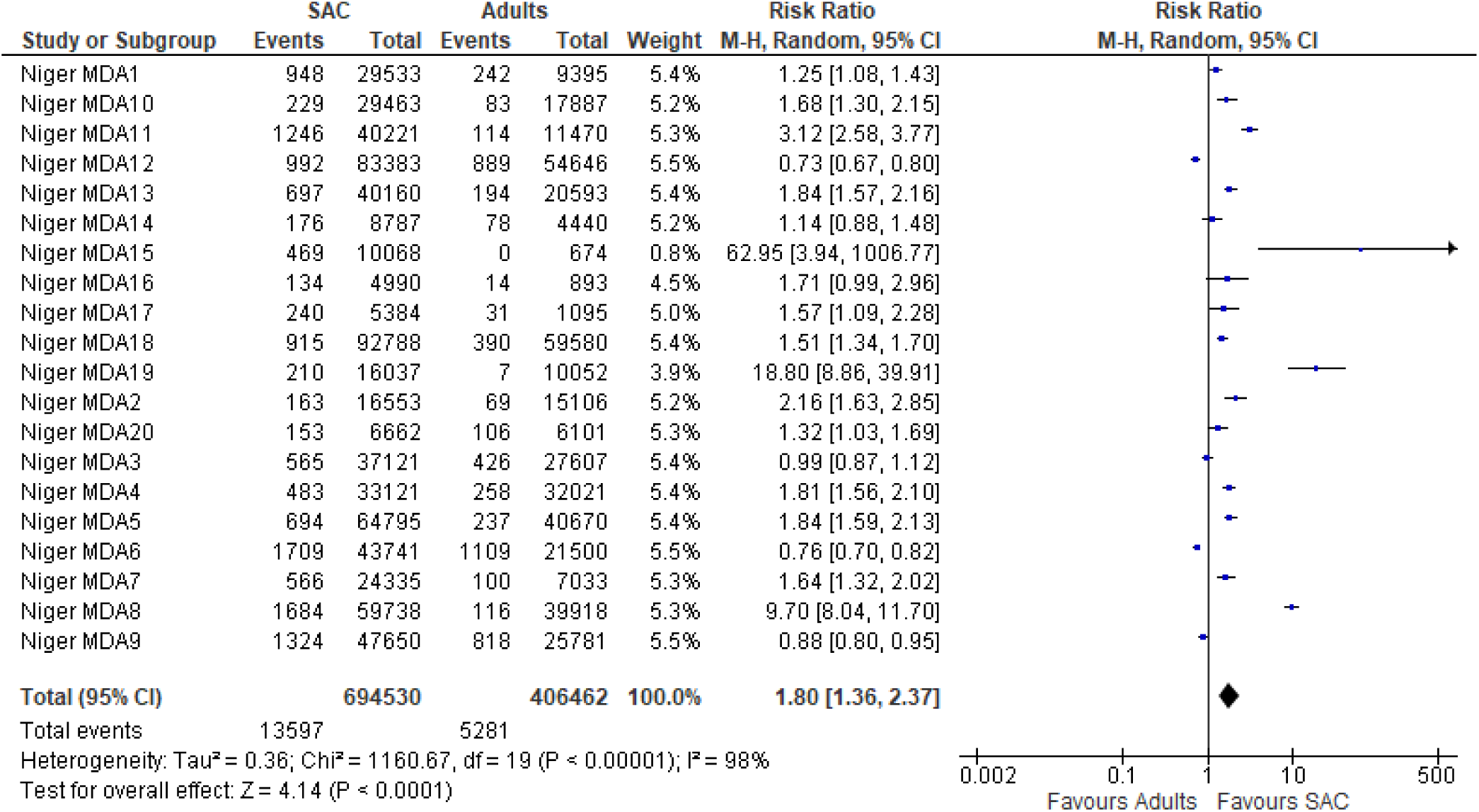
Dizziness associated with PZQ (40 mg/kg single dose) between school age children and adults in preventive chemotherapy programmes

### Adverse events in pregnant and lactating women

Eight studies (4 RCTs and 4 nRCTs investigated PZQ use in pregnancy. Of the eight studies, two RCTs (Bustindy et al 2020; Twenonyere et al. 2009) and 3 nRCTs (Chetrit et al. 2014; Adam et al. 2004; Qian et al. 2016) reported mild to moderate adverse events that were transient. The study by Chetrit et al. 2014 and the RCT by Bustindy et al. 2020 used a higher dose of PZQ at 60 mg/kg. Two RCTs reported a number of serious adverse events with either 40 mg/kg PZQ (Elliot et al. 2007) or 60 mg/kg (Olveda et al., 2016). Elliot et al. 2007 compared PZQ at the standard single dose of 40 mg/kg alone with PZQ plus ALB (400 mg) or PZQ plus placebo or ALB plus placebo. The PZQ plus placebo arm resulted in 4 miscarriages, 13 foetal deaths, 46 low birth weights and 43 congenital anomalies were reported among 626 women who received PZQ during pregnancy. No further details were provided for the type and nature of the congenital anomalies. Similar outcomes occurred in the PZQ plus ALB arm though where 4 miscarriages, 12 foetal deaths, 39 low birth weights and 50 congenital anomalies were reported among 628 women who received PZQ during pregnancy. The ALB + Placebo arm experienced 6 miscarriages, 11 foetal deaths, 45 low birth weights and 42 congenital anomalies among 629 women who were exposed to PZQ during pregnancy. The arm with the inactive ingredient (placebo) also experienced 4 miscarriages, 8 foetal deaths, 46 low birth weights and 42 congenital anomalies. Another RCT (Olveda et al., 2016) that assessed PZQ use during pregnancy compared with Dextrose reported adverse outcomes. This study though used higher PZQ dose schedule at 60 mg/kg administered as a divided dose (30 mg/kg x 2) and compared this with. The PZQ arm experienced 18 serious adverse events including 2 foetal deaths, 1 congenital anomaly and 2 cleft palate among 186 women who received PZQ during pregnancy. The women in the dextrose arm experienced 16 serious adverse events including one congenital anomaly and one cleft palate among 184 women included in the study. The study by Adam et al. 2005 conducted in Sudan and involved 25 pregnant women that assessed, retrospectively, adverse vents associated with PZQ during pregnancy reported one miscarriage (Table 8). The dose used in the study was not reported.

**Table 8.**
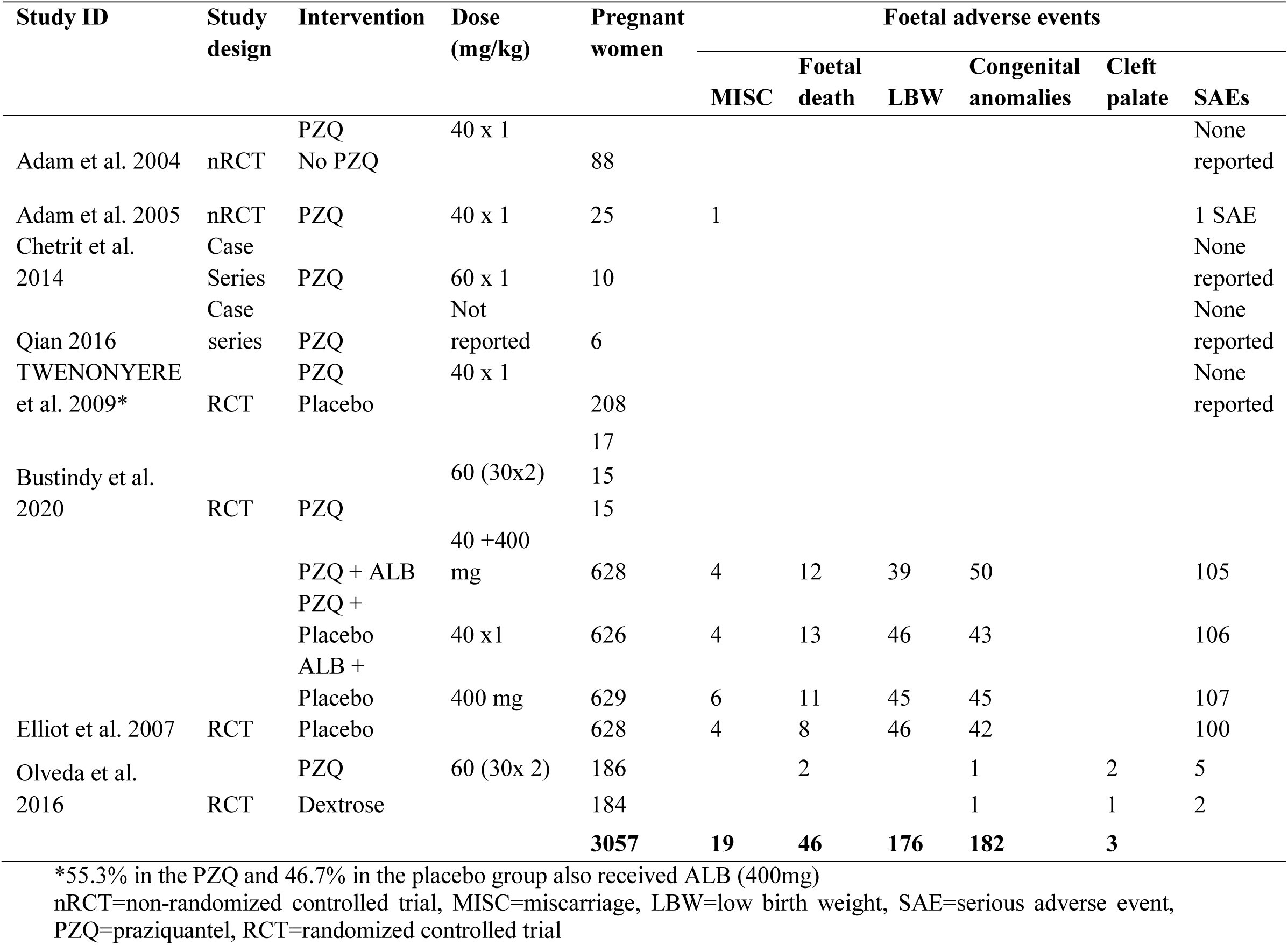
Adverse events associated with PZQ administration among 3057 pregnant women

### Visual adverse events in PZQ treatment

Five of the included studies were RCTs (Anto et al., 2011; Sissoko et al., 2009; Fu-yuan et al., 1984; Leopold et al., 1978; Rezende, 1985), and one was a case-series assessment study (Debesai & Russom, 2020b). Four of the five included studies were conducted in Africa, specifically Ghana (Anto 2011), Mali (Sissoko 2009), Tanzania (Rezende 1985) and Eritrea (Debesai & Russom 2020), one in China (Fu-yuan 1984) and one other in a non-endemic country in Germany involving healthy volunteers (Leopold 1978). One study assessed self-reported PZQ-related adverse effects in Eritrea (Debesai & Russom 2020). As expected, the study conducted in China was on treatment for *S. japonicum*. Table 8) ummarizes incidence and frequency of visual-related safety issues with PZQ from the four RCTs (Anto et al. 2011; Fu-Yuan et al. 1984; Rezende et al. 1985; Sissoko et al. 2009) and the cohort study (Leopold 1978). The study by Debessai & Russom (2020) has been reported in a separate table (Table 8). The study conducted in Ghana involved participants with *S. mansoni* and *S. haematobium* (Anto et al., 2011), in Brazil involved participants was concerned with *S. mansoni* (Rezende et al. 1985) and that from Mali (Sissoko et al. 2009) involved treatment for *S. haematobium* infections. A range of diagnostic procedures was used in the studies included in this review. They comprised sedimentation (Anto et al. 2011), circumoval precipitin test and rectal biopsy (Fu-yuan et al. 1984), Kato Katz (Rezende et al. 1985) and urine filtration (Sissoko et al. 2009). Three of the six studies included (Anto 2011; Sissoko 2009; Rezende 1985) involved school-aged children whereas two included only adults (Fu-yuan 1985, Leopold 1978). Rezende et al. (1985) included both SAC and adult participants.

Anto et al. (2011) reported incidence of some visual-related adverse events which co-administered PZQ with Albendazole and Ivermectin and compared this with Albendazole plus Ivermectin. A total of 180 adverse events were recorded out of which eight (8) were visual-related. Only eight participants out of 15,552 in the control arm (ALB + IVM) experienced visual-related events. The adverse events were similarly characterised as mild and did not require hospitalization. In the Eritrean study conducted by Debesai & Russom (2020), out of a total of 2579 Individual Case Safety Reports (ICSRs) of PZQ use that were retrieved from the WHO VigiBase database. Of the sixty-one reports confirmed to be visual disorders, 47 occurred in Ertitrea following PZQ administration. The authors applied the Austin Bradford-Hill criteria to assess a possible causal relationship between PZQ use and the reported visual disorders and the results were confirmed , the authors concluded that there was a strong association between PZQ use and there was a strong temporaly plausibile association that was consistent and specific. Leopold et al., 1978 reported a bizarre case of visual disorder in one participant out of the six that received 75 mkg/kg of PZQ in divided doses of 25 mg/kg. The authors reported that, one participant complained of a blurry vision and lack of concentration. This event occurred about 30 minutes after the patient received the second dose of PZQ.

**Table 9.** Summary of reported visual adverse events in the included RCTs

| Study | Mean age (SD)/yrs | Mean weight (SD)/kg | Treatment (Control) | Dose /mg/kg | No. of Visual disorders (N) | Nature of disorder |
| --- | --- | --- | --- | --- | --- | --- |
| Anto et al. 2011 | 12 (2.0) | 28.1 (8.51) | PZQ+ALB+IVM versus ALB+IVM | ** | 8 (6896) * | Reddening of eyes |
| Fu-Yuan et al. 1984 | -- | -- | PZQ<sup>+</sup> | 60 | 11 (476) | Blurred vision |
| Rezende et al. 1985 | 20.6 | 52.4 | PZQ (Oxamniquine) | 55 | 3 (272)* | Blurred vision |
| Sissoko et al. 2009 | 10.45 | -- | PZQ versus AS+SMP | 40 | 1 (392)* | Conjunctivitis |
| Leopold 1978 | -- | -- | PZQ versus Placebo | 25 | 1 (6) | Blurred vision |

\* Visual disorder occurred in the control group AS+SMP (Artesunate + Sulfamethoxypyrazine/Pyrimethamine)
\*\* Height-dosage relationship used where; 94–109 cm = 1 tablet; 110–124 cm = 1½ tablets; 125–137 cm = 2 tablets; 138–149 cm = 2½ tablets; 150–159 cm = 3 tablets; 160–177 cm = 4 tablets and ≥178 cm = 5 tablets
<sup>+</sup> Participants were group based on clinical presentation of schistosomiasis (i.e, advanced or chronic, presence of coinfections)

**Table 10.** Summary of reported visual adverse disorders from Debesai & Russom (2020)

| Sex | Number | Mean age (SD) | Reported visual disorders |  |  |
| --- | --- | --- | --- | --- | --- |
|  |  |  | Blurred vision (%) | Visual impairment | Lacrimation increased |
| Male | 18 | 23.06 (12.88) | 14 (29.8%) | 4 (8.5%) | 0 |
| Female | 29 | 28.72 (13.37) | 21 (44.7%) | 7 (14.9%) | 1 (2.1%) |
| Total | 47 | 26.63 (13.33) | 35 (74.5%) | 11 (23.4%) | 1 (2.1%) |

### PZQ adverse events in taeniasis

Two studies (Braae et al., 2016; Ramiandrasoa et al 2020) reported on the use of PZQ in treating Taeniasis. In two of the studies (Braae et al 2016 and; Ramiandrasoa et al 2020), the treated species was *Taenia solium* but the species in Hong 2018 was not known as the study relied on data from a drug safety surveillance database where safety issues were self-reported. The two former studies both administered PZQ at the WHO recommended dose of 10 mg/kg for treating Taeniasis, much lower than the 40 mg/kg for treating Schistosomiasis. Hong 2018 reported 108 cases (some events overlapped in people who co-administered Albendazole) of adverse events but it is unclear how many people were treated although the author estimates in excess of 152,000 Korean people between 2012 and 2015. Apparently, only a single case out of the 108 was found to be associated with PZQ treatment. Braae et al 2016 used a “track and treat” approach in administering PZQ to people infected with Taeniasis after a PZQ MDA to treat targeted at treating Schistosomiasis. None of the 20 people treated with 10 mg/kg PZQ experienced any adverse event. Ramiandrasoa et al 2020 recorded adverse events in less than 0.5% in each of the three rounds of MDA that treated 221,308 people. Majorly, nausea and temporary diarrhoea were the reported adverse events.

**Table 10.** Adverse events of PZQ after treatment or taeniasis

| Study | Country | Worm | PZQ dose | Number treated | Adverse events | Serious adverse events | Remarks |
| --- | --- | --- | --- | --- | --- | --- | --- |
| Ramiandrasoa et al 2020 | Madagascar | *Taenia solium* | 10 mg/kg | 221,308 | Minor side effects mainly nausea and diarrhoea. 0.4%, 0.3% and 0.2% of AEs were recorded during the 1<sup>st</sup>, 2<sup>nd</sup> and 3<sup>rd</sup> MDA rounds | None reported | Adverse events were monitored both actively and passively |
| Braae et al 2016 | Tanzania | *Taenia solium* | 10 mg/kg | 20 | No AEs were recorded during the study but it is unclear how AEs were assessed. | None reported | The study tracked and treated participants infected with Taeniasis after rounds of Schistosomiasis MDA. |

N/A=Not applicable, NR=Not reported, MDA= Mass Drug Administration, AEs= Adverse events

### Risk of bias assessment in the included RCTs

Majority of the included RCTs were of good quality methodologically with several studies presenting sufficient details of the methods of the RCTs. Some studies, though, did not report how the random sequence was generated making it difficult to ascertain methodological rigour. Allocation of participants was concealed in a good number of studies using varying methods but mainly through the use of sealed envelopes. Apart from three open-label studies, all other included RCTs reported blinding of either participants, investigators or both in allocation, administering treatments and/or assessing outcomes. Few studies reported substantial loss to follow-ups whereas some studies did not explore the effect of incomplete outcome data on the resultant outcomes. In all the included RCT studies, the reported outcomes or main conclusions were at par with individual study objectives thus reducing reporting bias. To a large extent, no other biases were identified but, in some studies, sample sizes were relatively small. See Appendix for the results of risk of bias assessment and the authors judgement.

### Risk of bias assessment in the included cohort studies

The validity of the design of a cohort study depends on sufficient follow-ups, proper matching and a study without selection bias. If follow-ups are carried out correctly, the exposed and unexposed groups are checked for the outcome. Adequate matching often caters to all potential confounders that can impact the research result. However, the internal validity and strength of the study can be compromised by the impact of losing patients during follow-up in cohort studies. In addition, in designing intervention groups, selection bias is likely to cause a systemic error, allowing them to vary in terms of prognosis.

The sampling source for all the cohort studies was properly carried out with reference to the current review, except for a couple of them who could not identify their sample source. Exposure and outcome are defined as appropriate if the outcome is expected by the exposure. In this respect, the exposure measurement of the sample and the result were satisfactory. For most of the cohort studies, though, matching was not performed. Therefore, the result of the research will be influenced by potential behaviour characteristics. On most of the research, follow-ups were performed adequately, except for a few of them that were inconclusive.

**Table 4.**
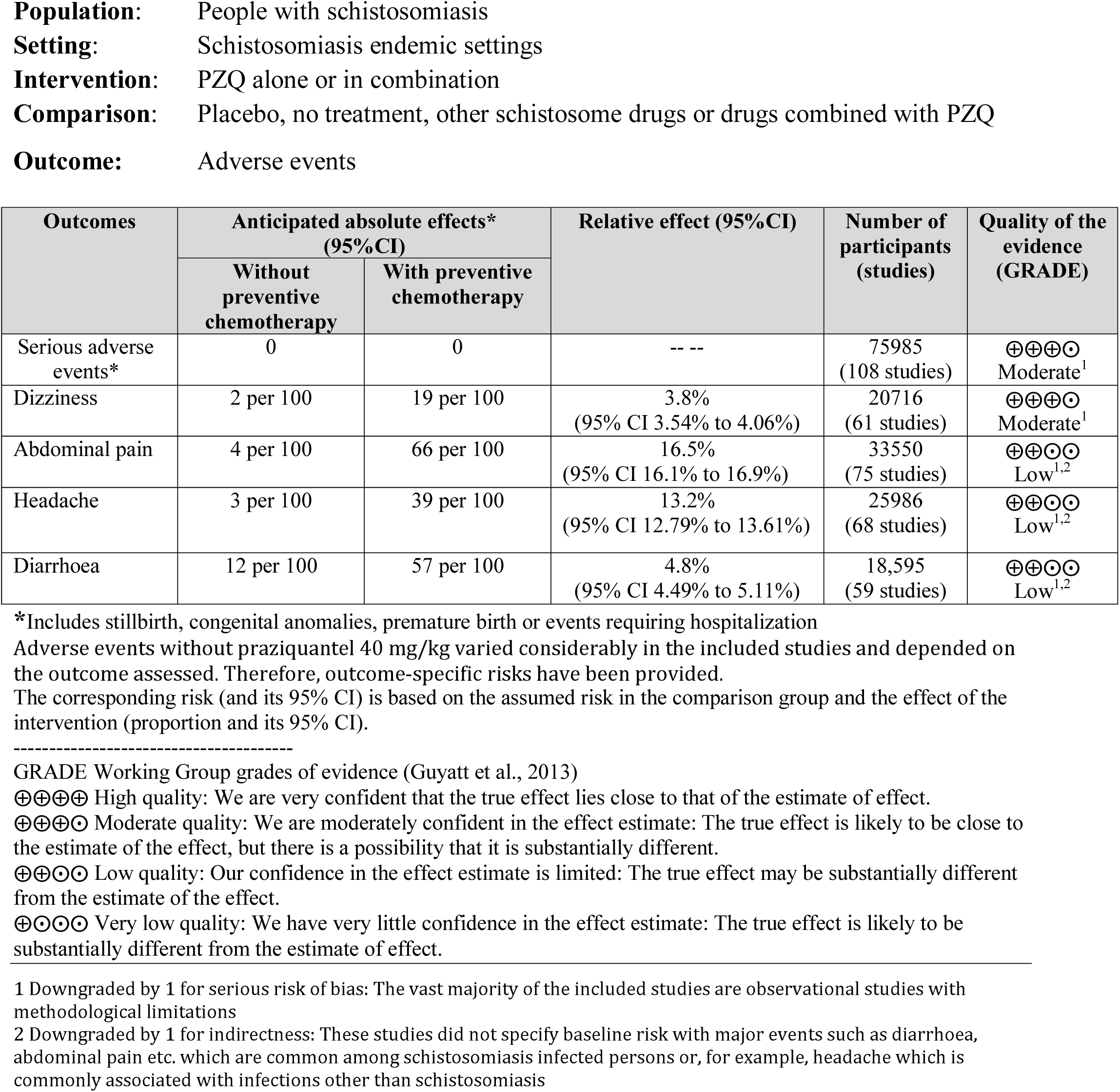
Grading the level of evidence using GRADE

## DISCUSSION

Most of studies reported mild-to-moderate and transient adverse effects indicating that PZQ is safe. Adverse events were mild-to-moderate and transient. From the meta-analyses that compared school age children and adults, there were excesses of adverse events in the SAC groups for all the events analysed but these were mild-to-moderate and no serious event was reported. However, the few studies that assessed safety in pregnant women reported some serious adverse events (miscarriage, foetal deaths and congenital anomalies) which raises safety concerns. Given that those in the non-PZQ arms also experienced serious adverse events in almost similar numbers means that the evidence is inconclusive and further research is warranted. Similarly, the few studies that focused on PZQ-related visual adverse events reported incidence of serious visual adverse events after PZQ treatment, raising further concerns about PZQ. Again, there was limited or no data on long term adverse events (including blood biochemistry and organ pathology), safety in co-morbidity states and co-infection with taeniasis, calling focused and robust analysis targeted to these specific questions. Although this systematic review and meta-analysis included 129 studies, most of them were limited by sample sizes, very short follow-up times that captured only immediate adverse events (missing long term events associated with PZQ use) and methodological rigour. We did not find any study that had investigated safety of PZQ between those with infection and those without infection to compare event rates. This has serious public health implications and raises an important ethical debate as persons without infection who receive PZQ could be very high in some settings where endemicities are going down and the number of persons without infection that get treatment could reach as high as 90% in some communities earmarked for PC. An important observation, is that studies on schistosomiasis adverse events research generally lack methodological rigour in design, methods, conduct and reporting. Given that PZQ is now administered repeatedly in endemic populations, further effort is needed to obtain and assess in detail the aforementioned areas with limited evidence to inform sound guidance and policies in the future.

Considering the severe adverse events, one study (DaSilva et al, 1986) reported 5cases of severe dizziness associated with PZQ intake but the event subsided within two hours of administration. This study did not explore any causal assessment of the event. In one study (Minggang et al, 1983), 98 serious adverse events including hepatic coma, epileptic seizures, atrioventricular block were reported for treatment for *S. Japonicum*, but the investigators used dose schedules that were slightly higher (60 mg/kg, 70 mg/kg and 90-120 mg/kg). In this retrospective study, with varying dose schedule, the minimum reported schedule was 45mg/kg and others were as high as 120 mg/kg over extended period of over six days. The investigators reported that those who received PZQ treatment had acute or advanced schistosomiasis. The evidence obtained from this study was inconclusive because some of the participants had other medical conditions such as chronic hepatitis, nephropyelitis, neurosis etc. which could confound the association between the praziquantel and the serious adverse events. For example, it was realized that 8patients who experienced seizures were epileptic. Nonetheless the study was retrospective thus reducing the robustness of the evidence. Also, causal assessments could not confirm from this study. Though inconclusive, we are therefore, concerned with the likelihood of serious events when praziquantel is administered to persons with chronic conditions. We thus recommend that administration to persons with underlying other medical conditions such as epilepsy, Hepatitis, etc. should be done with caution.

Another study of interest was Berhe et al, 1999, which recorded some severe adverse events. Berhe reported generally transient and mild symptoms with most of them resolving on their own. However, 90 of the 611 patients experience serious adverse events associated with gastrointestinal disorders such that they were administered antispasmodics and analgesics. This study is a cohort study and causal relationship was not be established, and more so as the variables needed to assess association between PZQ were not properly reported..

### Meta-Analysis of Events

A subgroup meta-analysis conducted on 3 studies reveal that individuals taking 40mg/kg of praziquantel are 57% reduced risk of vomiting compared to individuals taking in praziquantel 60mg/kg. Therefore, it is imperative that where there is no evidence of possible resistance to praziquantel or an evidence of impaired renal function, then prescribers should maintain a praziquantel dose of 40mg/kg during management. Since increasing the dose to 60mg/kg may associated with an increased risk of vomiting amongst the patients. The meta-analysis for the comparison of the events such as headache, vomiting and abdominal pain associated with praziquantel administration as against other doses of praziquantel or placebo reveals that there is no increased risk of event with praziquantel compared to the other doses or placebo. Since the included studies were all randomized control trial the meta-analysis is robust. But the number of trials included in this meta-analysis were not many reducing the power and the reliability of statistically detecting any differences.

The researchers of this review conclude that there is no evidence of serious adverse events associated with praziquantel intake at various doses. Considering the 108studies that were included in this review, the evidence is that praziquantel is still safe for administration though it may be associated with mild to moderate side effects such as headache, dizziness, diarrhea and abdominal pains. However, these events are highly tolerable and do not require hospitalization. Though most studies included in this review were old and have reduced robustness in the method of conduct, we found little or insufficient evidence to believe that praziquantel is not safe for administration to persons who are not pregnant. We recommend the continuous use of praziquantel 40mg/kg for mass drug administration. The reviewers recommend further investigation on the safety of praziquantel for administration to persons with underlying medical conditions such as neurological disorders, cardiovascular events and other organ damages.

### Comparison between school age children and adults

From the meta-analyses that compared school age children and adults, there were excesses of adverse events in the SAC groups for all the events analysed but these were mild-to-moderate and no serious event was reported. However, the few studies that assessed safety in pregnancy reported serious adverse events (miscarriage, foetal deaths and congenital anomalies) to raise safety concerns. Nonetheless, given that those in the non-PZQ arms also experienced serious adverse events in almost equal numbers means that the evidence is inconclusive and further research is warranted. Similarly, the few studies that focused on visual adverse events reported severe visual adverse events raise concerns for further research as this review could not provide answers to the underlying factors. Again, there was limited or no data on long term adverse events (including blood biochemistry), safety in co-morbidity states, mixed schistosome infections and co-infection with taeniasis, calling future focused analysis or new studies generate tailored evidence. Although this systematic review and meta-analysis included 123 studies, most of them were limited by sample size, very short follow-up times that captured only immediate adverse events and missing long term events associated with PZQ use and methodolical rigour. We did not find any study that had investigated safety of PZQ between those with infection and those without infection. This raises serious public health concerns and ethical implications as persons without infection who receive PZQ could reach as high as 90% in communities earmarked for preventive chemotherapy. Generally, studies on adverse events in schistosomiasis as identified in this systematic review lacks methodological rigour in design, methods, conduct and reporting. Given that PZQ is now administered repeatedly in endemic populations, further effort is needed to obtain and assess in detail the aforementioned areas with limited evidence to inform sound guidance and policies in the future.

### Quality of the evidence

GRADE methodology was implored in the assessment of the quality of evidence and presented in summary of findings (SOF) tables for the main comparisons. a 4-point scale was used to assess the level of quality of the evidence. High quality evidence implies that researchers have high levels of confidence in the effect estimate and that further research is unnecessary. Moderate quality evidence implies lower confidence in the result and further research may have an important impact on the result. Low and very low-quality evidence shows the increasing ambiguity in the result and an imperative for further research to be conducted. The evidence presented is generally considered to be of moderate or low quality due to concerns related to three key factors: i) most studies included in this review were very old thus reducing the robust ness of the findings, ii) there poor evidence in the reporting of the methods of the included studies resulting in some limitations, iii) poor reporting of adverse events and lack of causal assessment for the reporting of the events and iv) the number and size of the trials being small and often underpowered to reliably detect statistically significant differences.

### Safety of praziquantel in pregnant women

Schistosomiasis has become one of the most prevalent parasitic infections and has significant economic and public health consequences. Studies across the globe have reported on the safety of praziquantel in pregnancy. No records of severe human pregnancy adverse effects have been reported over the years of experience with praziquantel. In a retrospective study by Adam et al., (2004) in Sudan, 88 pregnancies with inadvertent exposure to praziquantel, including 37 with exposure during the first trimester, were compared to 549 other pregnancies with no exposure to praziquantel. Clinical examination of babies born to exposed or exposed women found that none of the 88 exposed pregnancies resulted in abortion or stillbirth and no congenital defects were reported (Adam et al 2004). The challenge with this study is that the study was retrospective and as such, the dose of administration could not be assessed. Since the incidence of adverse event is dependent on drug dosage, it is inconclusive to establish from this study that praziquantel is safe. Again, as stated in the study, the proportion of respondents assessed, 88, was too small to generalise especially for adverse events that are rare, the incidence of the adverse event may not be observed in this study.

Another prospective study which involved 25 pregnant Sudanese women with *S. mansoni* infections were treated with a single dose of praziquantel, at 40 mg per kg, during different trimesters (Adam et al 2005). While at 10 weeks of gestation, one of these women had a spontaneous abortion, this adverse event cannot be attributed to the intervention because it happened about 3 weeks after administration of praziquantel. We therefore rate this causality assessment as highly unlikely. This is because the rate of abortion among the treated women was close to the baseline rate in the study population. Even in the first trimester of pregnancy, it appeared that PZQ was a safe medication to use against *Schistosomiasis mansoni*. Both studies reported praziquantel is safe in pregnancy, however possible limitations might have affected the outcome or results of the study. In as much as cohort studies are rated very highly in terms of level of evidence for an intervention study, in retrospective cohort studies data collected may not originally collected for the study purpose and hence may be incomplete which could affect the outcome of the study. In addition, they could be a recall bias since participants may give information, which may not be a true reflection of themselves.

For a small case series of schistosomiasis travel-related acquisitions, four pregnant women were treated with 60 mg per kg of praziquantel, including two treated during the first trimester. For mothers or neonates, treatment did not culminate in any significant adverse reactions (Ben-Chetrit et al, 2015). Nonetheless, the case series cannot be generalised because there are several confounding variables that limits the robustness of this study. In the treatment of neurocysticercosis, daily doses of praziquantel for up to 21 days were administered to pregnant women without any apparent adverse events (Paparone 1996).

Considering more robust method of assessing the safety of praziquantel in pregnant women, randomized controlled trials were considered in this review. A trial in Leyte, the first elaborate study of the pharmacokinetics of praziquantel in pregnant women was performed (Bustinduy et al, 2020). The findings of this research, which included assays quantifying the concentrations of praziquantel in the milk of treated women, showed that administration of praziquantel in pregnant women did not present any severe adverse event. The limitation of this study is that the primary outcome was to look at the pharmacokinetics of the drug thus the sample size of the study was small to generalise the findings of adverse events of this study. For rare adverse events, huge sample size would be required. The recorded adverse events in the Leyte trials included Headache, diarrhoea, fever, apnoea (Table 2). Most of these adverse events were mild and did not pose any threat to the pregnancy of the mothers. Another concern from this study is that the dose of the drug administered is 30mg/kg at 3 hours apart. This dosage is different from the dosage often used for mass drug administration hence the expected incidence of adverse event may differ.

Another trial that assessed the safety of praziquantel in pregnant women is the Entebbe Mother and Baby study. This study assessed pregnant women who had infection and those who did not at a dose 40mg/kg. This design approach and dosage imitates the mass drug administration recommended by the WHO. The Entebbe mother and Baby trial assessed several outcomes and reported as several publications (Elliot et al 2007, Twenyogyere et al 2011, Ndibazza et al 20, Nampija et al 2012, Webb et al 2011, Namara et al 2017). Thus, for this review, the publications were viewed as separate studies. For Elliiot et al, report on the safety of praziquantel amongst pregnant women was not stated. Therefore, it is inconclusive to draw the safety of praziquantel amongst these high-risk groups. Due to the robustness of the study and the appropriateness of the method of conduct, the evidence that praziquantel administered to pregnant mothers did not influence anaemia and birthweight of the child. The study observed that there were congenital anomalies and perinatal mortalities, and these were evenly distributed across the arms. However, the study failed to assess the causality of the adverse event.

Olveda et al in a clinical trial conducted in North Eastern of Leyte in Philipines assessing the immunological response of mothers at 12-16 weeks of gestation to praziquantel. This study is powered compared to the Bustindy et al. Olveda reported 18 severe adverse events. The study reported congenital anomalies such as talipes equinovarons and foetal cleft palate. For the club feet, the study identified even distribution of this event in both arms of the study. Yet the incidence of cleft palate was 2:1 in the praziquantel arm against the comparator. A similar anomaly of cleft palate was reported in a case series by Qian et al., which seem to suggest that there could be a potential case of cleft palate induced by praziquantel. Through it all, the study did not establish the causality of these serious adverse events and this is a limitation to this study. Considering the study participants, it can be argued that the participants were within the second trimester and thus foetal development may be far advanced. Thus, the incidence of congenital anomaly may be reduced compared to mothers in the first trimester.

### Visual adverse event of praziquantel

Several studies have assessed the adverse effect of praziquantel and have reported several events with headache, diarrhea and vomiting been predominant events of praziquantel administration. Recently, there has been concerns of possible adverse effect of praziquantel on vision.

Visual disorders following praziquantel chemotherapy have recently been reported by an Eritrean study, presenting an alarming debate on the safety of the continued use of the drug in treating schistosomiasis. Any such adverse events are not described as the known ‘side effects’ of the use of the drug which chiefly presents with vomiting, dizziness and abdominal discomfort after ingestion. The current review drew on all studies that have reported on the safety of praziquantel, specifically to ascertain the evidence regarding the occurrence of visual disorders after PZQ chemotherapy. None of the five studies included reported serious adverse events requiring hospitalization. In fact, in two of these studies, visual disorders rather occurred in the groups that were not treated with PZQ. Anto et al 2011 recorded eight cases of ‘reddening of the eyes’ in the control area of their study which sought to assess the adverse effects associated with the co-administration of albendazole, praziquantel and ivermectin. In their two-arm comparative study, participants in the control area received only ivermectin and albendazole, and adverse effects were recorded through passive surveillance. Generally, participants that received praziquantel together with the other two drugs had significantly higher reports of headache and general body weakness but all such cases were mild and did not require hospitalization. Existing studies does not support the evidence of praziquantel having an adverse visual effect.

### Comparison of side effects between SAC and adults

Over the years, there have been several concerns about the dilapidating effect of Schistosoma on the growth and development of infected children. As such the WHO in 2010 recommended the administration of praziquantel in children particularly pre-school attending children (Coulibaly 2007). This is to help reduce the burden of schistosomiasis on the children. Since schistosomiasis in children has been associated with anaemia (Friedman et al 2005) impaired growth and impaired cognitive function (Nampijja et al 2012; Ezeamam et al 2005).

From the meta-analysis, it is observed that for most of the side effects witnessed, the risk of experiencing an adverse event was high in the school attending children compared to the adults. Several studies such as Stelma et al 1995; Berhe et al 1999 have attributed the increased risk of side effect and its intensity to the pre-treatment Schistosoma egg load. From these studies, it was concluded that increased Schistosoma egg load was associated with an increased intensity of side effects. Additionally, several studies have established that in the Schistosoma egg load is increased in school attending children compared to adults. Thus, This increased risk of side effect in School attending children is due to the viral load. since the incidence of side effect largely depends on the drug concentration of praziquantel in blood, an age-related model by Bonate et al 2018 reveals that since the metabolism of praziquantel depends on the CYP activity, the drug remains in circulation longer, resulting in a higher AUC in children due to decreased CYP activity. Thus, the increased likelihood of experiencing side effects in children compared to an adult.

### Authors Conclusions

Praziquantel showed to be safe as frequently reported adverse events were mild-to-moderate and transient, and although there were some severe events, these resolved without further intervention. Only few studies reported serious adverse events but these raise concerns. Given that adverse events studies have assessed incidence of events within a very short window, intermediate to long term events would have been missed with serious public health implications. Although the evidence is inconclusive, the fact that some of the few studies that assessed safety in pregnant women reported some serious events, calls for caution in the inclusion of pregnant women in preventive chemotherapy campaigns. The results from studies suggesting that Praziquantel is associated with serious visual adverse events raise concerns but the evidence is inconclusive. This systematic review has exposed the lack of methodological rigour in adverse events studies and recommends that future research should incorporate robust designs, methods, conduct and reporting, using quality-assured tools and following best practices. Further research is needed to obtain and collate topic-specific empirical evidence in the following areas: tissue organ damage and blood chemistry analysis to provide evidence on long term safety of Praziquantel, as well safety in pregnant women and their foetuses. Other areas that need further evaluation are Praziquantel use in those with co-morbidity and on polypharmacy, mixed schistosome infections and schistosome co-infection with other diseases such as taeniasis, filariasis and trachoma, as well as co-administration with drugs used in other preventive chemotherapy programmes.

## Data Availability

All data produced in the present work are contained in the manuscript.

## APPENDIX 1 SEARCH TERMS

((((((((((((((((((((((("praziquantel"[MeSH Terms] OR "praziquantel"[All Fields]) OR distocide[All Fields]) OR ("praziquantel"[MeSH Terms] OR "praziquantel"[All Fields] OR "biltricide"[All Fields])) OR pharmamed[All Fields]) AND ("safety"[MeSH Terms] OR "safety"[All Fields])) OR "side effects"[All Fields]) OR "adverse drug reactions"[All Fields]) OR "adverse events"[All Fields]) OR ("toxicity"[Subheading] OR "toxicity"[All Fields])) OR ("immune tolerance"[MeSH Terms] OR ("immune"[All Fields] AND "tolerance"[All Fields]) OR "immune tolerance"[All Fields] OR "tolerance"[All Fields] OR "drug tolerance"[MeSH Terms] OR ("drug"[All Fields] AND "tolerance"[All Fields]) OR "drug tolerance"[All Fields])) OR "safety profile"[All Fields]) OR "side effect"[All Fields]) OR "adverse drug reaction"[All Fields]) OR "adverse event"[All Fields]) AND ("schistosomiasis"[MeSH Terms] OR "schistosomiasis"[All Fields])) OR ("schistosoma"[MeSH Terms] OR "schistosoma"[All Fields])) OR ("schistosoma"[MeSH Terms] OR "schistosoma"[All Fields] OR "schistosome"[All Fields])) OR ("schistosoma"[MeSH Terms] OR "schistosoma"[All Fields] OR "schistosomes"[All Fields])) OR ("schistosoma"[MeSH Terms] OR "schistosoma"[All Fields])) OR mansoni[All Fields]) OR japonicum[All Fields]) OR intercalatum[All Fields]) OR haematobium[All Fields]) OR mekongi[All Fields] AND ("2009/01/01"[PDAT] : "2017/08/31"[PDAT])

### Study selection flow chart

**Fig 2.**
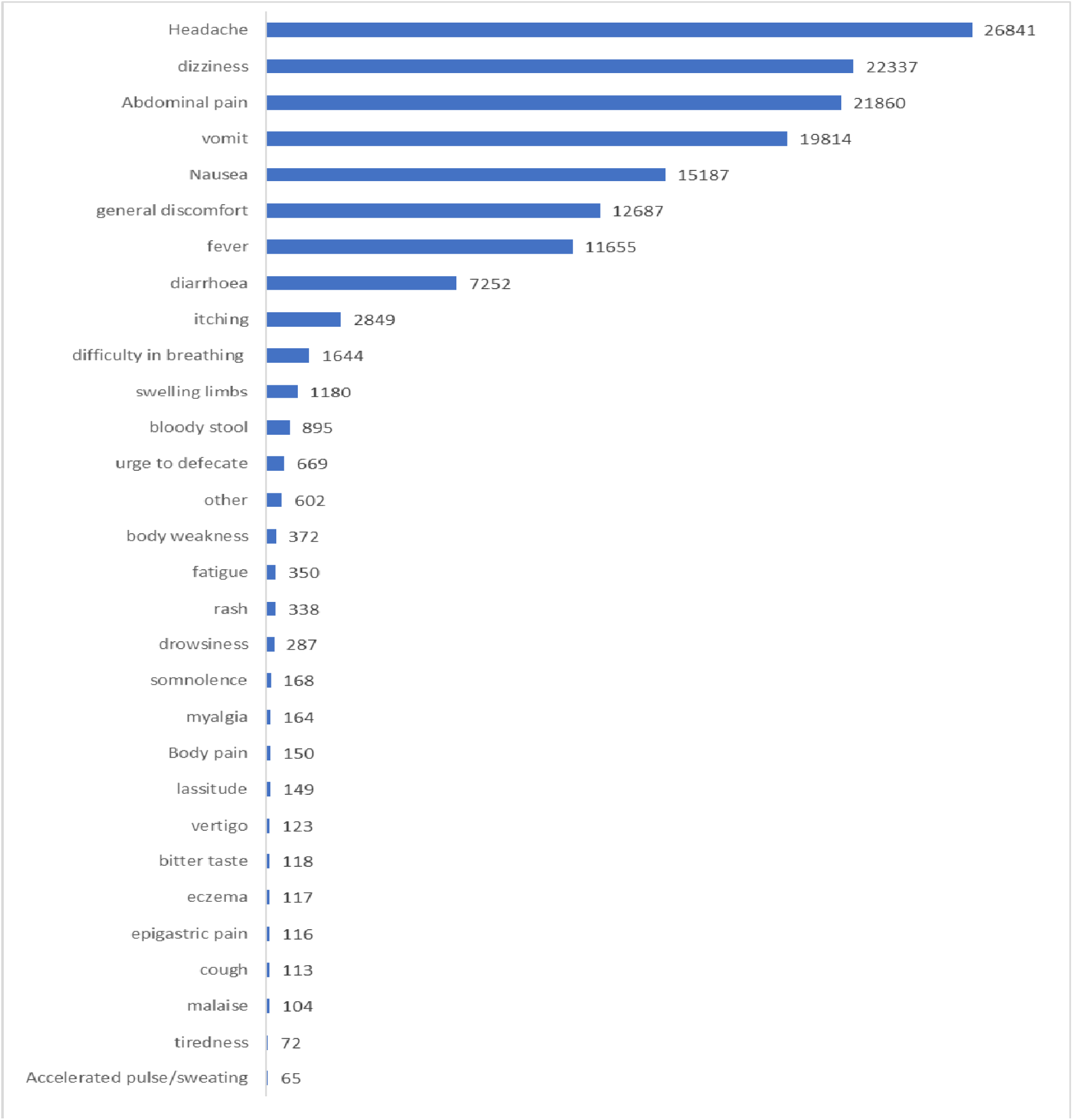
The first 30 common adverse events experienced and reported by persons with or without schistosomiasis who received PZQ (arranged in decreasing frequency)

**Fig 3.**
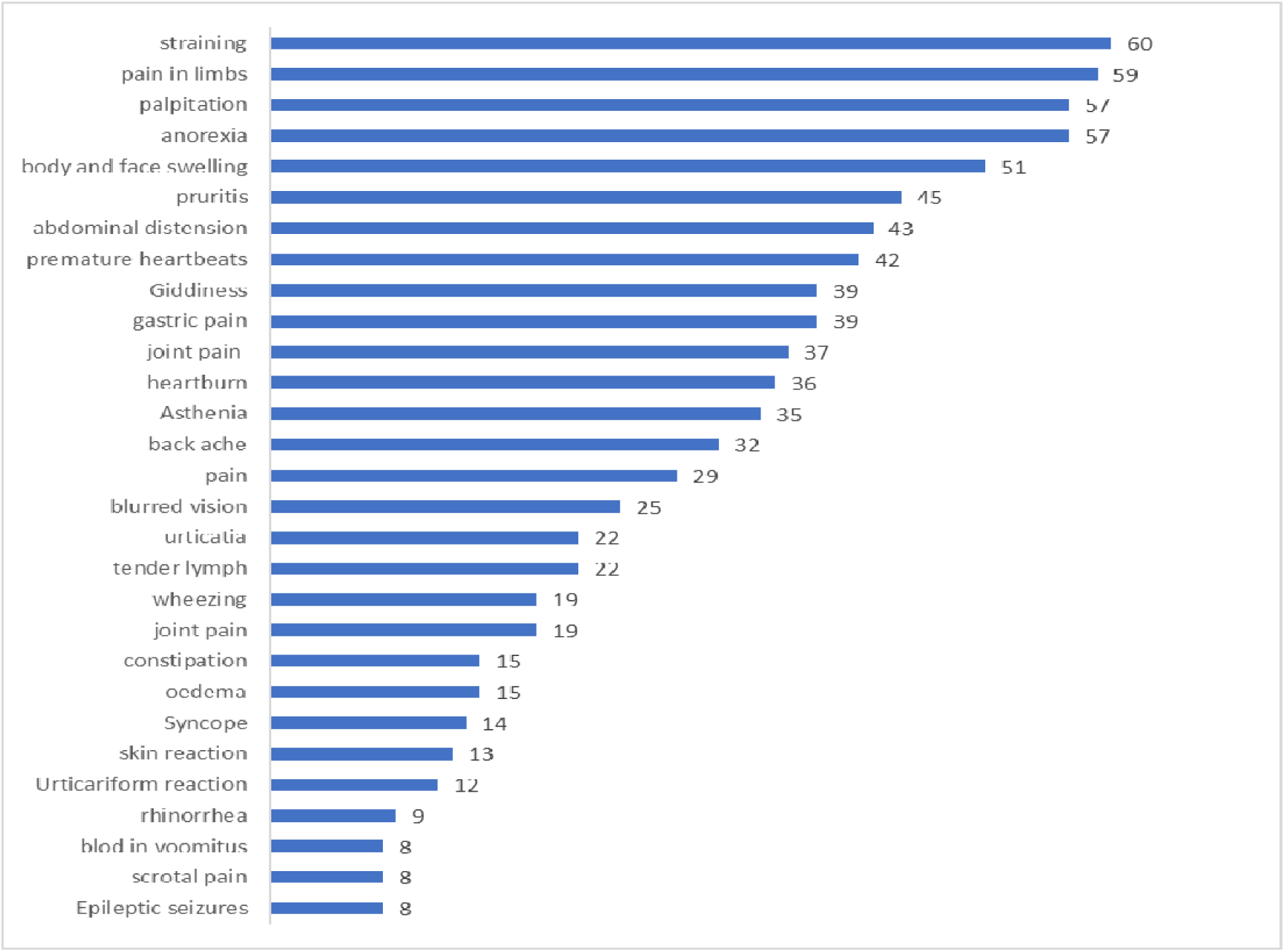
The second set of adverse events (number 31 to 60 events) arranged in decreasing frequency of events

**Fig 4.**
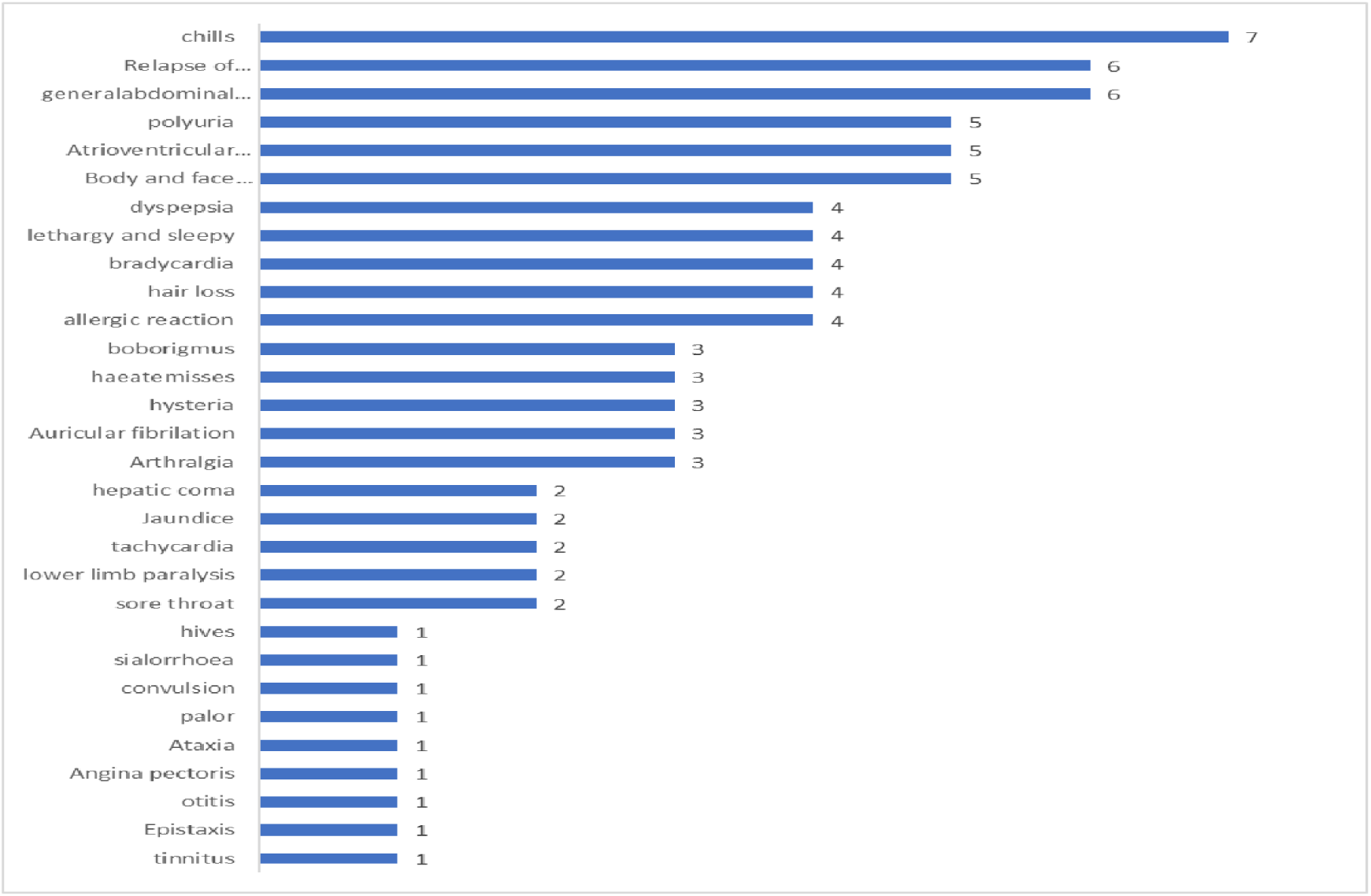
The third common adverse events (number 61 to 90 events) in persons with and without schistosomiasis who received PZQ (arranged in decreasing frequency of events)

### Comparing adverse events of PZQ40mg/kg single dose between SAC and Adults

**Fig 12.**
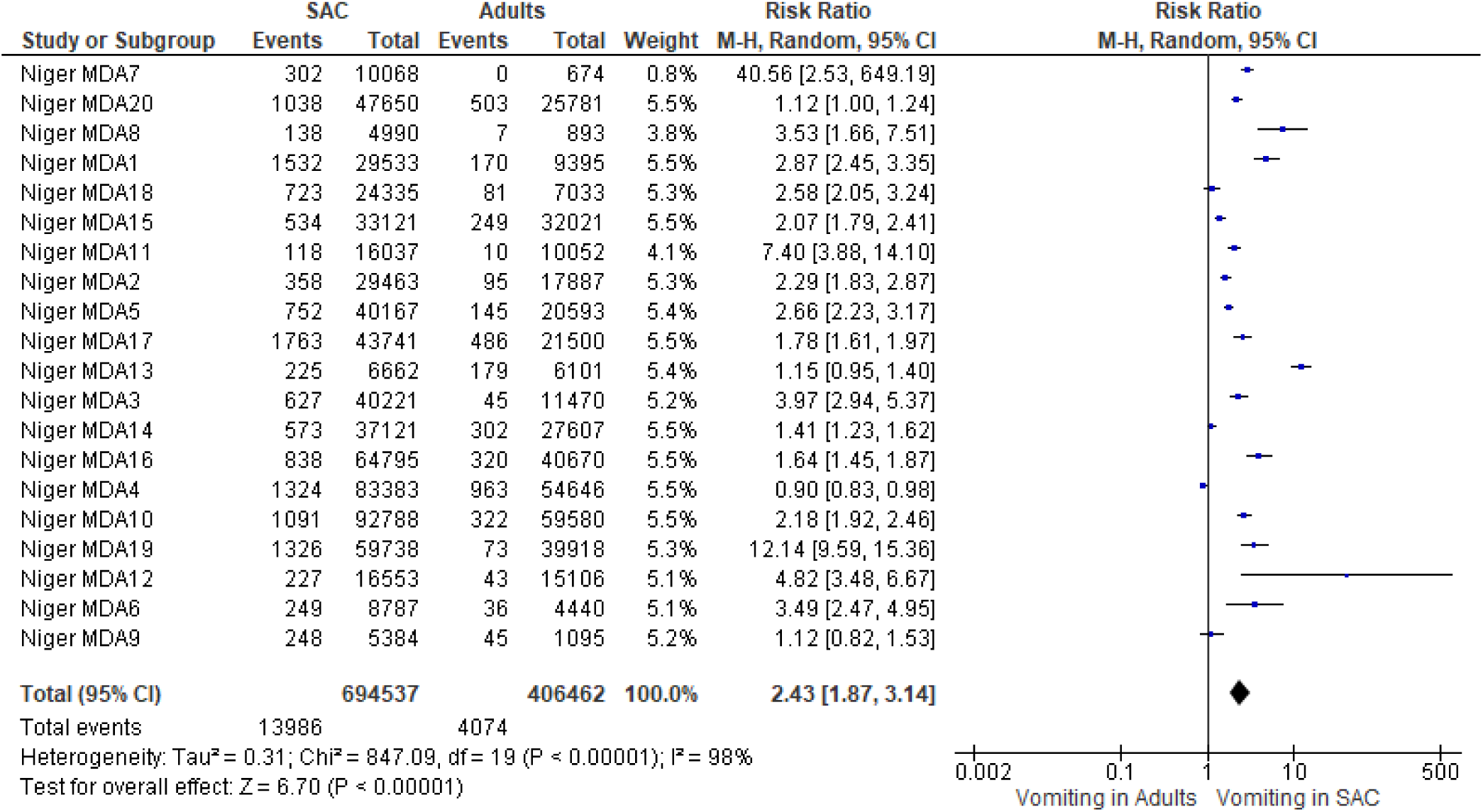
Vomiting

**Fig 13.**
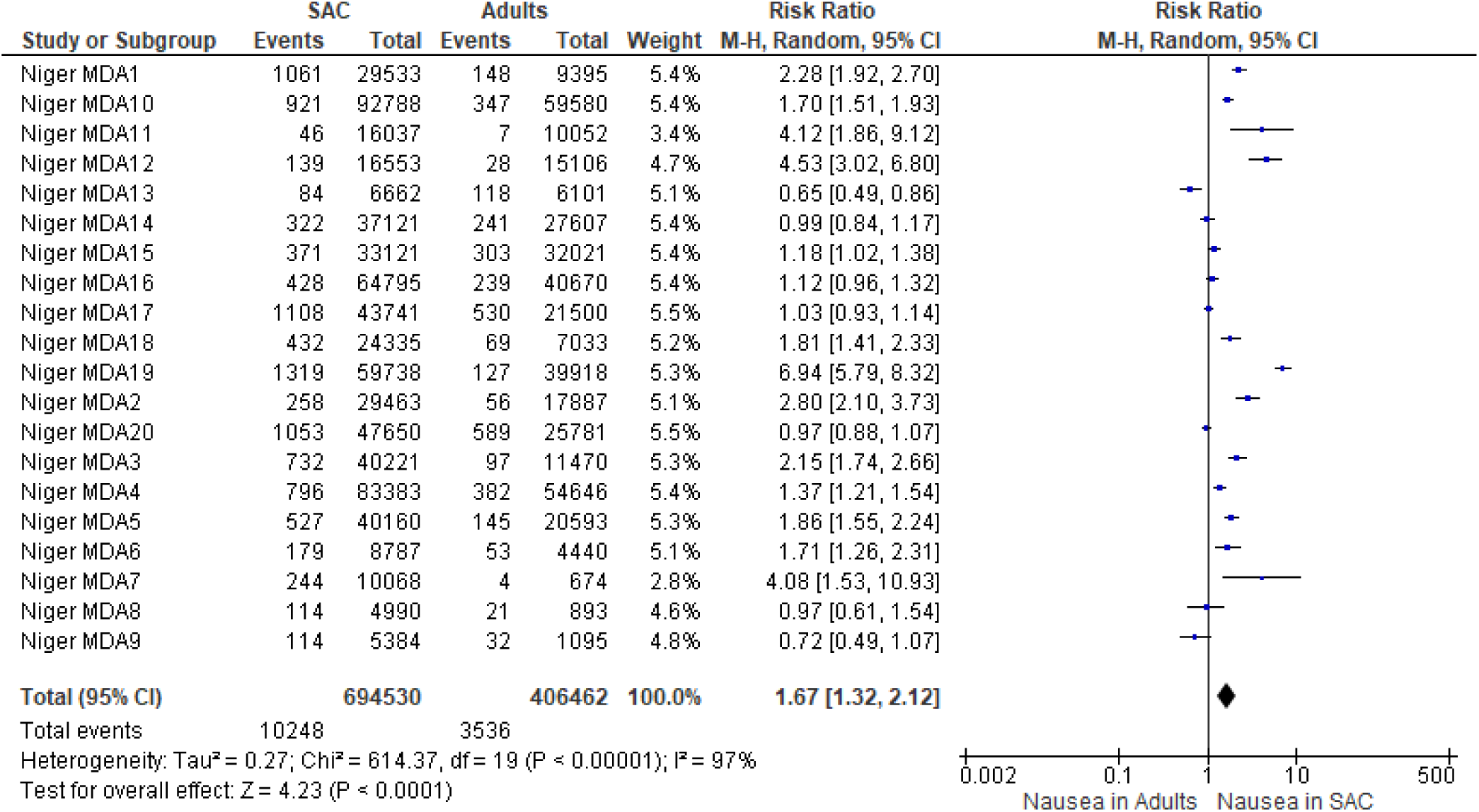
Abdominal pain

**Fig 13.**
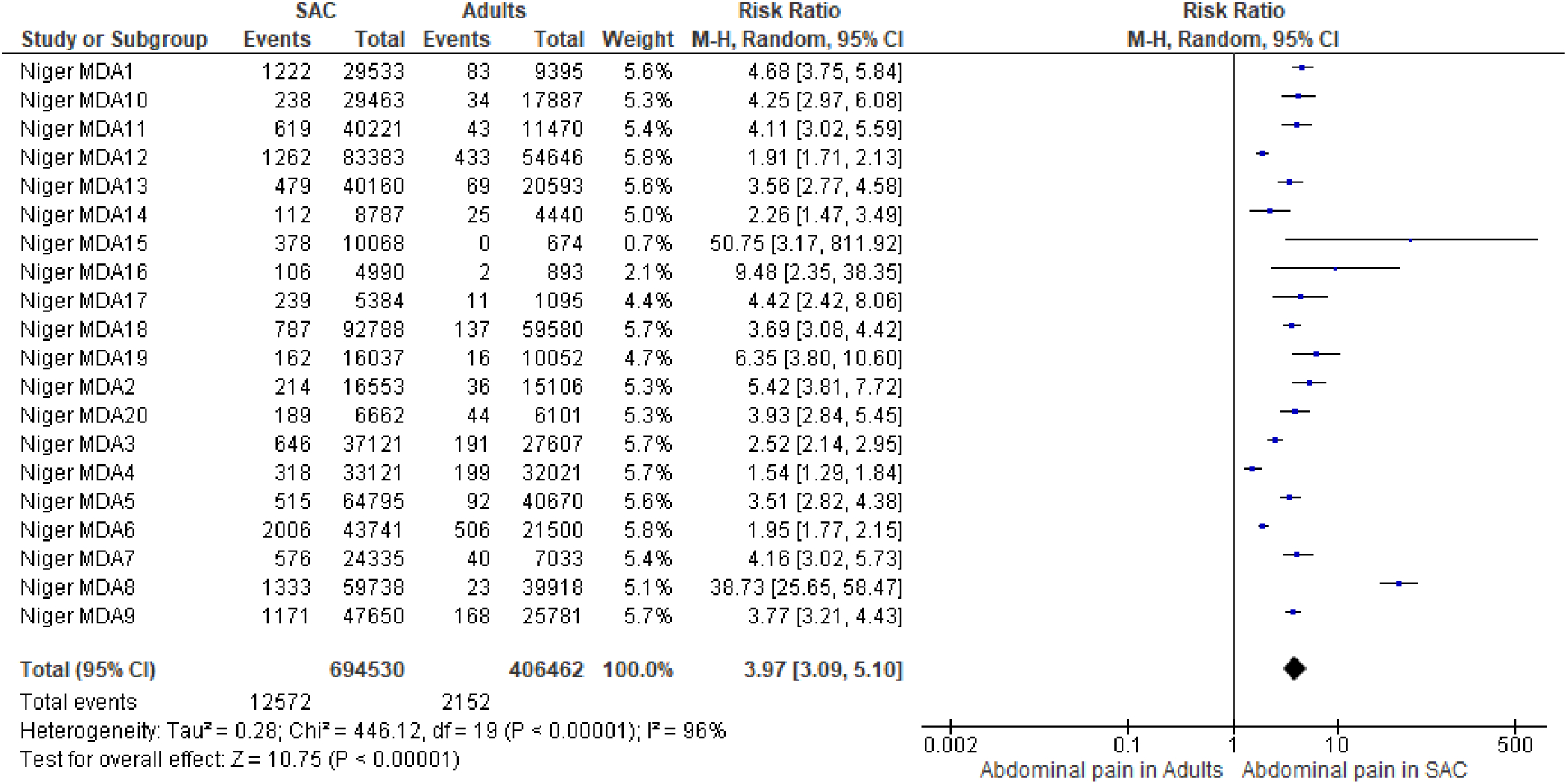
Nausea

**Fig 14.**
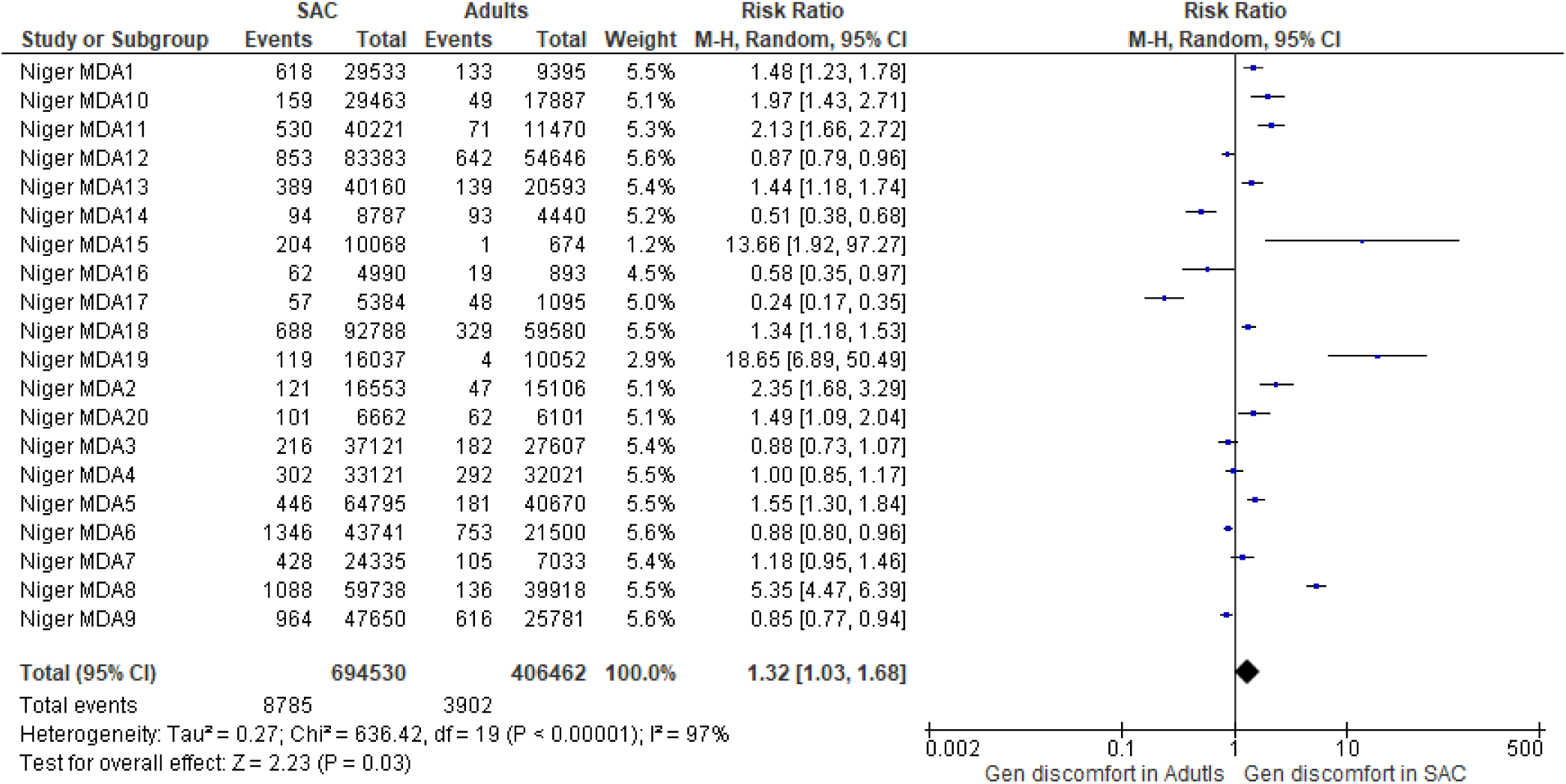
General discomfort

**Fig 15.**
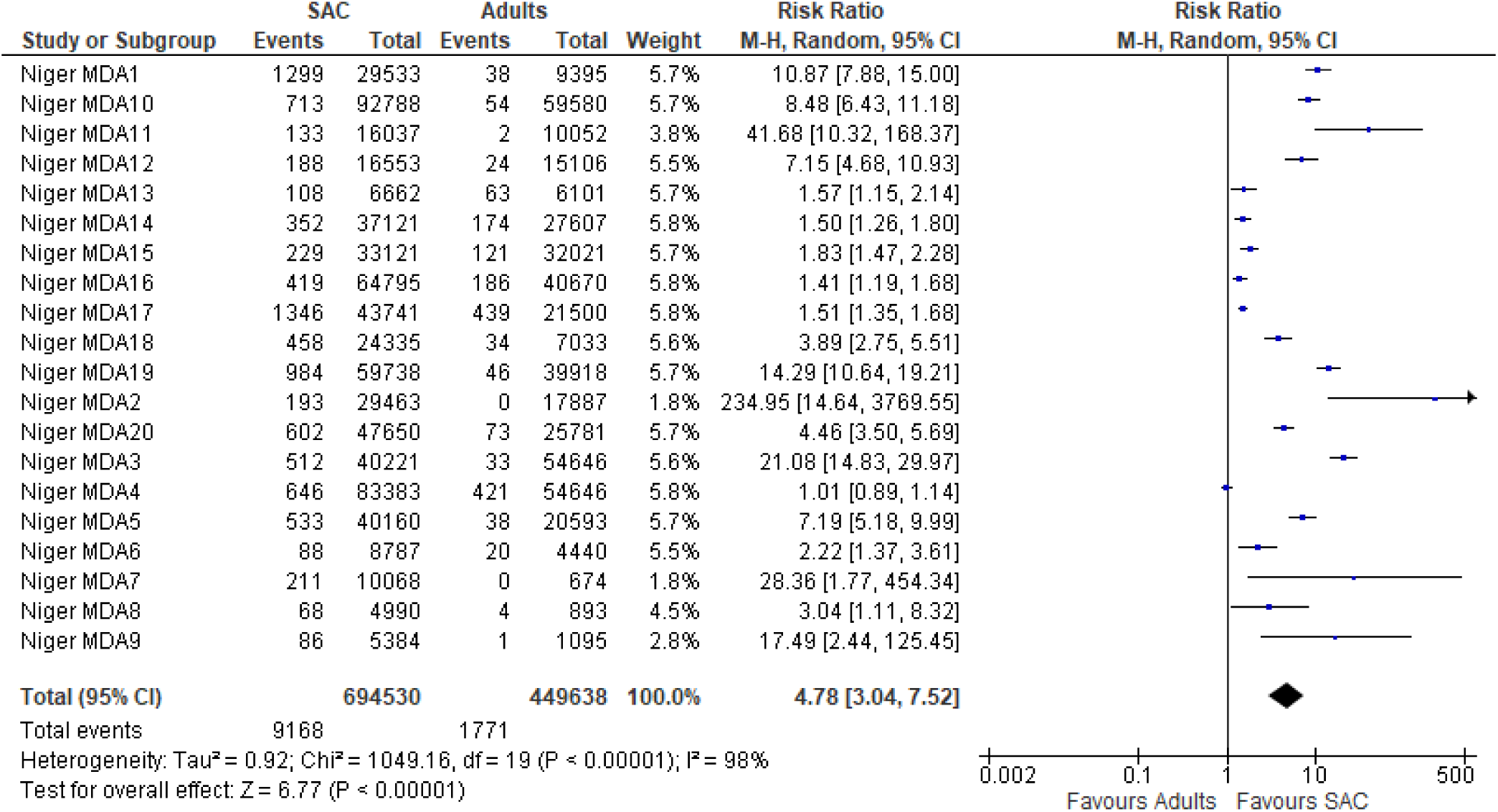
Fever

**Fig 16.**
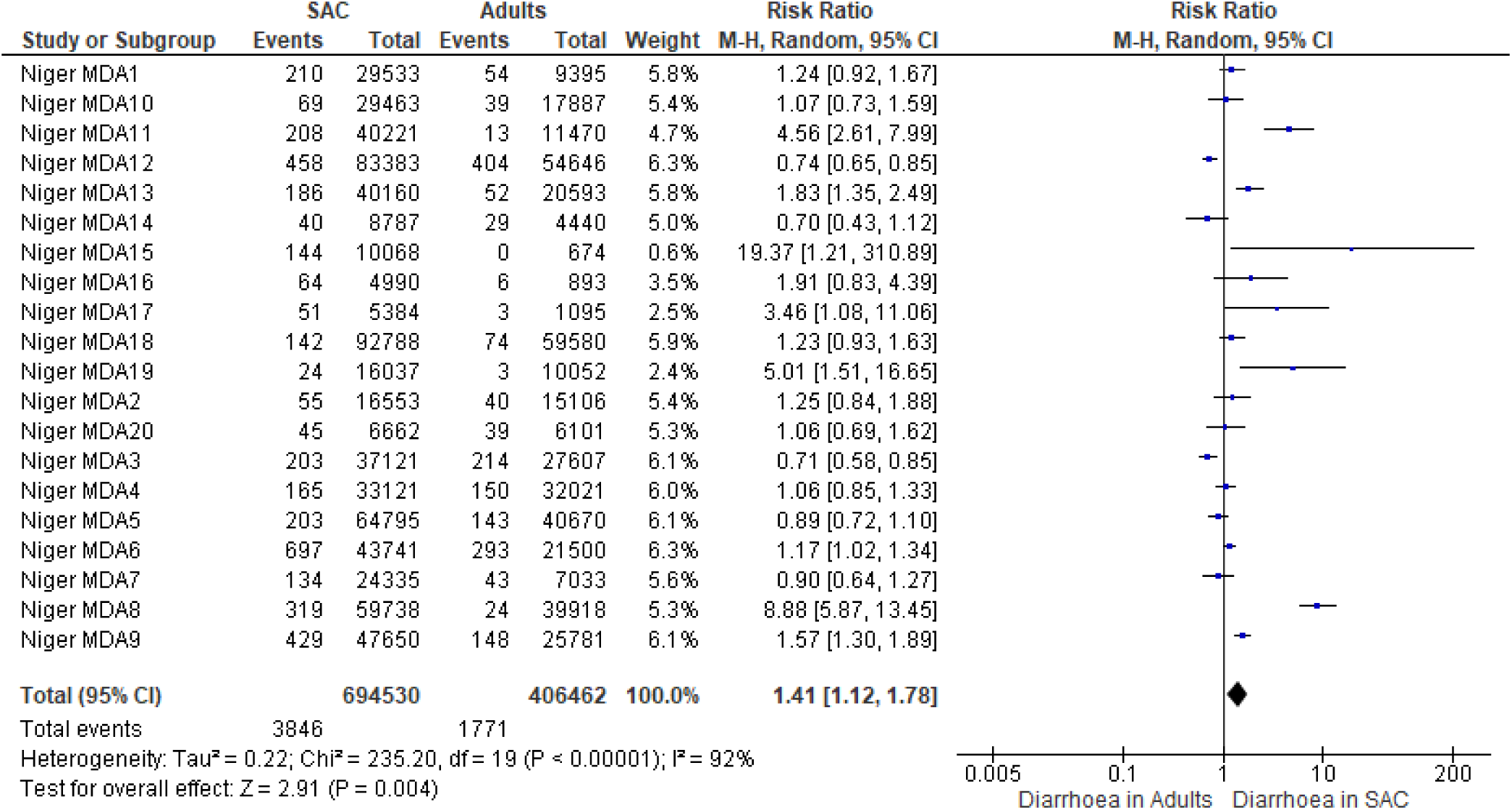
diarrhoea

**Fig 17.**
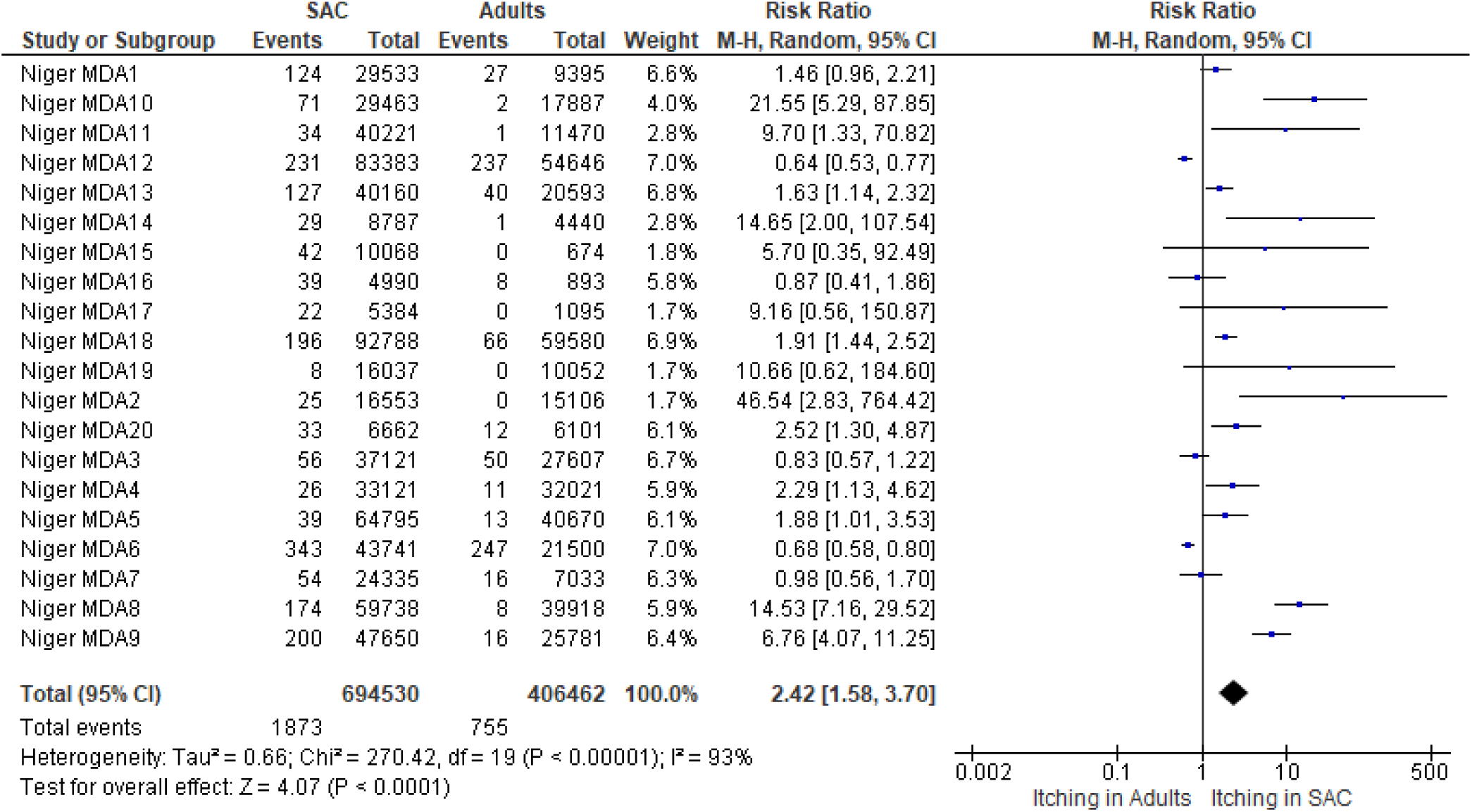
Itching

**Fig 18.**
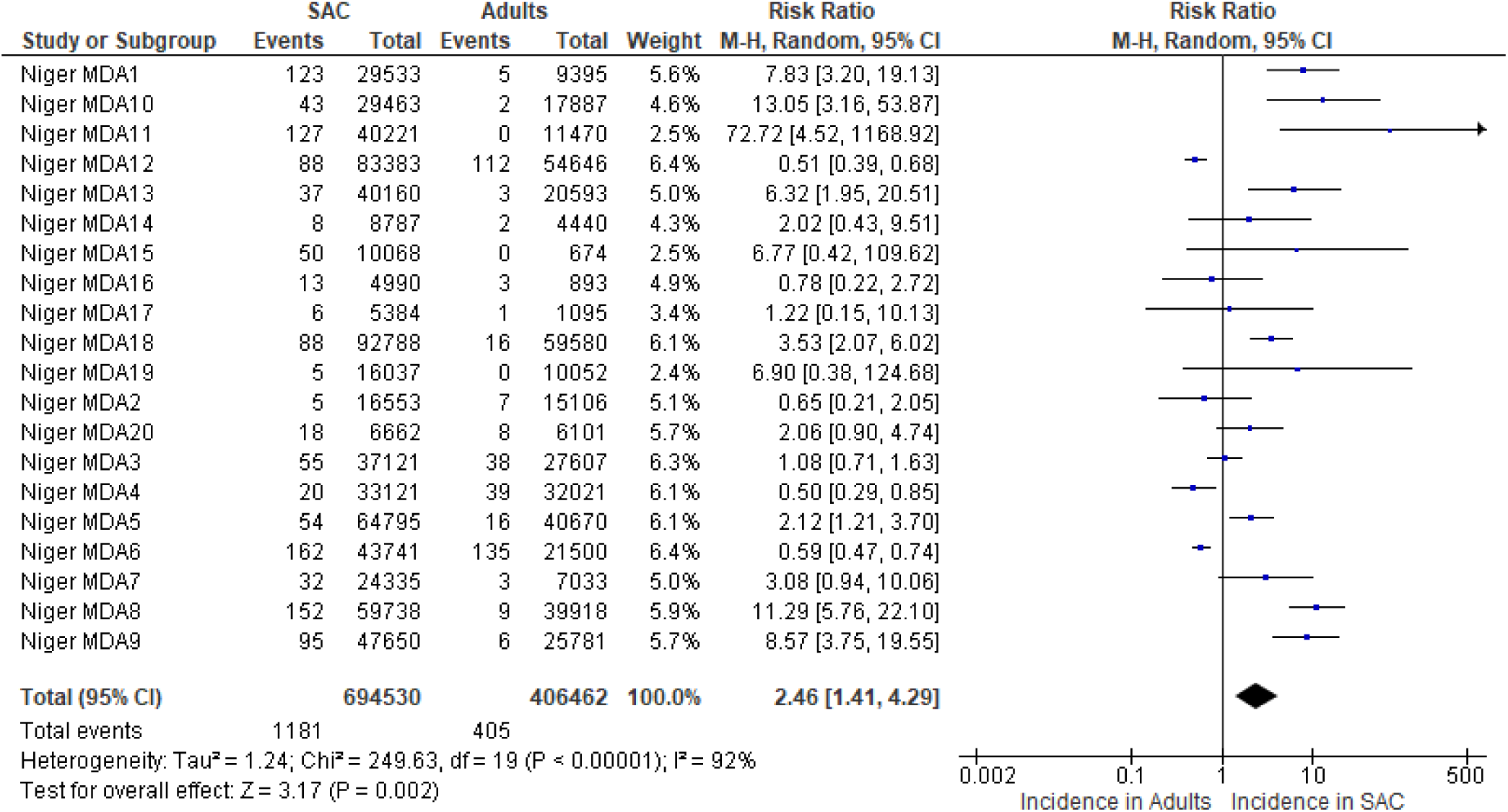
breathing difficulty

**Fig 19.**
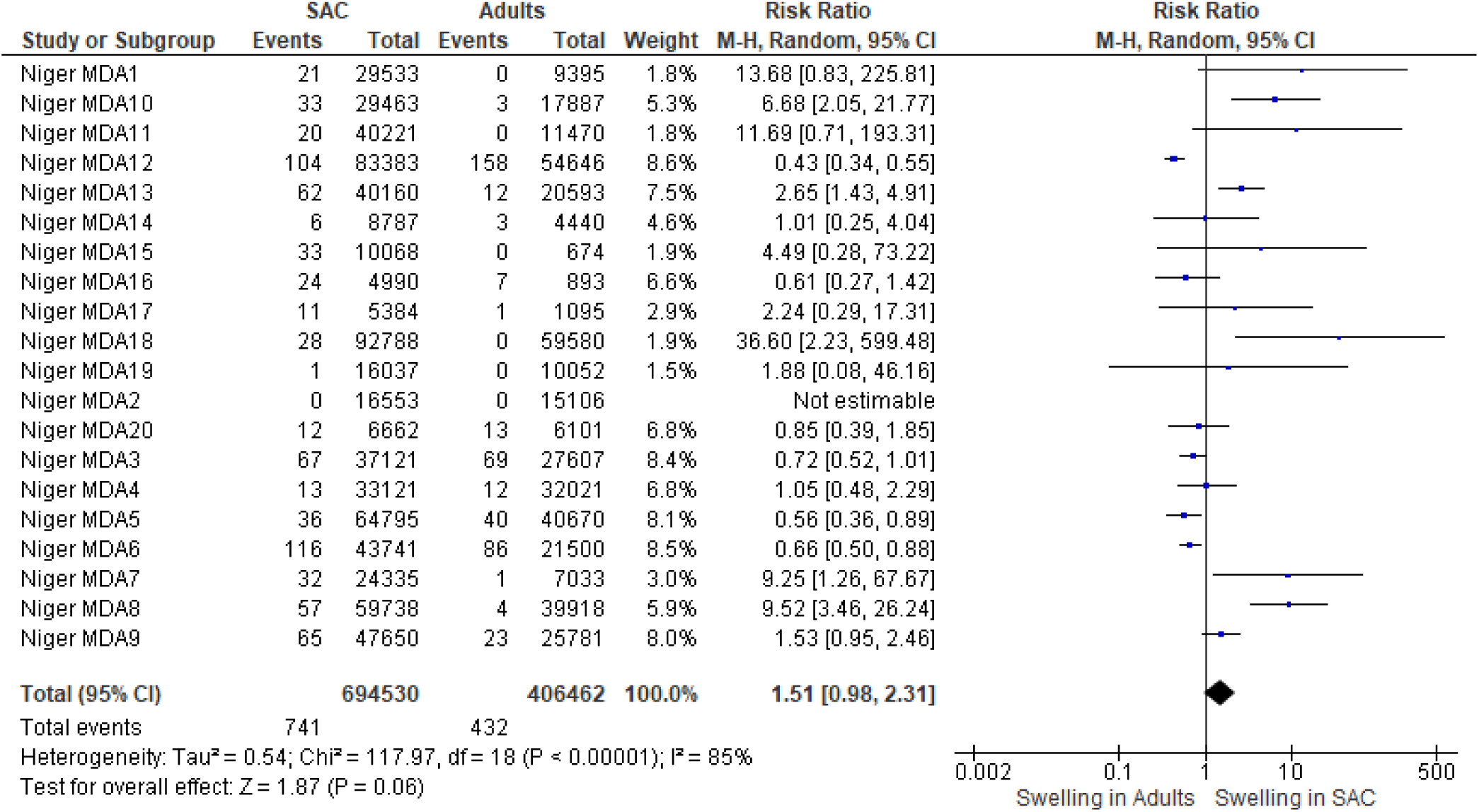
swelling

**Table 5.**
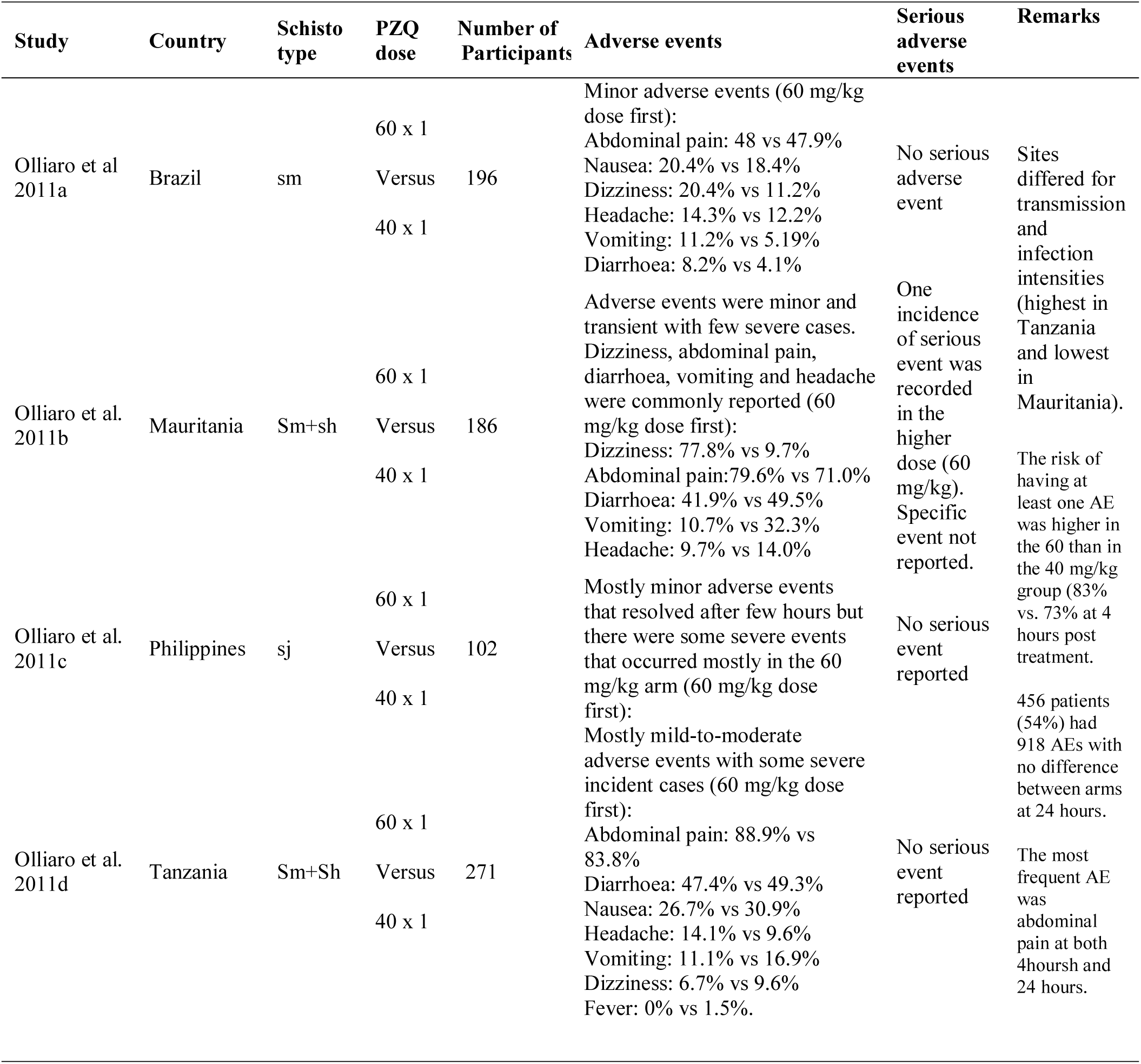
Comparing PZQ 40 mg/kg with higher doses in RCTs

**Table 6.** Adverse events in mixed infection with *S. mansoni* and *S. haematobium*

| Study | Country | Study design | Number of participants | Dose of PZQ (mg/kg) | Adverse events | Serious adverse events | Remarks |
| --- | --- | --- | --- | --- | --- | --- | --- |
| Ahmed et al. 1988 | Egypt | Not stated |  | 40 |  |  |  |
| Anto et al. 2011 | Ghana | RCT | 22448 | * | Prevalence of AEs reported as 8.02 per 1000. All AEs were mild, transient and required no hospitalization | None reported | The study did not explicitly report AEs in the 3 participants with coinfection |
| Coulibaly et al. 2012 | Cote D'Ivoire | RCT | 234 | 40 | 43 children reported adverse events. **12/43** children had Sm + Sh coinfection. No further information on severity were reported | None reported | But for 4 cases of body and face inflammation classified as moderately severe, all adverse events were mild and resolved on its own |
| El-Alamy et al. 1981 | Egypt | Clinical Trial | 353 | 40 | All adverse reactions categorized as mild and self-limiting. Reported signs included fever, rapid pulse and rash. Symptoms included nausea, abdominal pain, dizziness, etc. | None reported | The selected signs and symptoms were present before treatment but increased in frequency post-treatment |
| Garba et al. 2013 | Niger | Clinical Trial | 877 | 40 | After 1<sup>st</sup> dose, 33% of SAC reported AEs within 4 hrs post-treatment; 6.2% reported AEs within 4-24 hrs post-treatment. After 2<sup>nd</sup> dose, 1.5% of SAC reported AEs within 4hrs post-treatment and none within 4-24hrs AEs were chiefly abdominal pain, sleepiness and headache | None reported | No further information provided on the distribution of AEs. It is unclear how participants with Sh+Sm coinfections were affected |
| Groning 1985 |  | MDA |  | 40 |  |  |  |
| Schutte et al. 1983 | South Africa | RCT | 294 | 40 | AEs were more frequent immediately after treatment compared with 48 hrs post-treatment. There were all mild and required no treatment. | None reported | AEs were more common in participants with Sh+Sm coinfection (Embokodweni) compared with those with only Sh infection (Emophela) |
| Kardaman et al. 1983 | Sudan | RCT | 43 | 40 (20x2) | In the camps (n=81) where there were 43 | None reported | There was no significant |
|  |  |  |  |  | confected participants, abdominal pain (22.2%), vomiting (19.8%), diarrhoea (14.8%), etc.<br>At other sites (Group A and Group B), similar trend of AEs was recorded. |  | difference in the frequency and severity of AEs observed in the three sites. AEs were described as mild and resolved within 24 hrs |
| Wilkins 1987a |  | ?? | 184 | 40 | No serious adverse event. Commonly reported side effects included headache, weakness, dizziness, nausea/vomiting, diarrhoea, abdominal pain, general malaise, and fever. Among these events, abdominal pain, general malaise, and fever were reported more frequently with PZQ treatment | None reported |  |
| Omer et al. 1981 | Sudan | RCT | 153 | 40 (20x2) | None reported |  | All AEs were mild and transient. ECG changes were found to be minimal and unalarming. |
| El-Masry et al. 1988 | Egypt | RCT | 204 | 60 (30x2) | Epigastric pain was the most common AE (up to 66.6% of participants who received 20mg twice daily for 3 days), headache, nausea, vomiting and fever up to 38°C were also common | Not reported | AEs occurred less often in patients treated with 30mg dose and more often in those treated with 120mg |

\*height-dosage relationship used where; 94–109 cm = 1 tablet; 110–124 cm = 1½ tablets; 125–137 cm = 2 tablets; 138–149 cm = 2½ tablets; 150–159 cm = 3 tablets; 160–177 cm = 4 tablets and ≥178 cm = 5 tablets

### RISK OF BIAS ASSESSMENT FOR INCLUDED RCT STUDIES

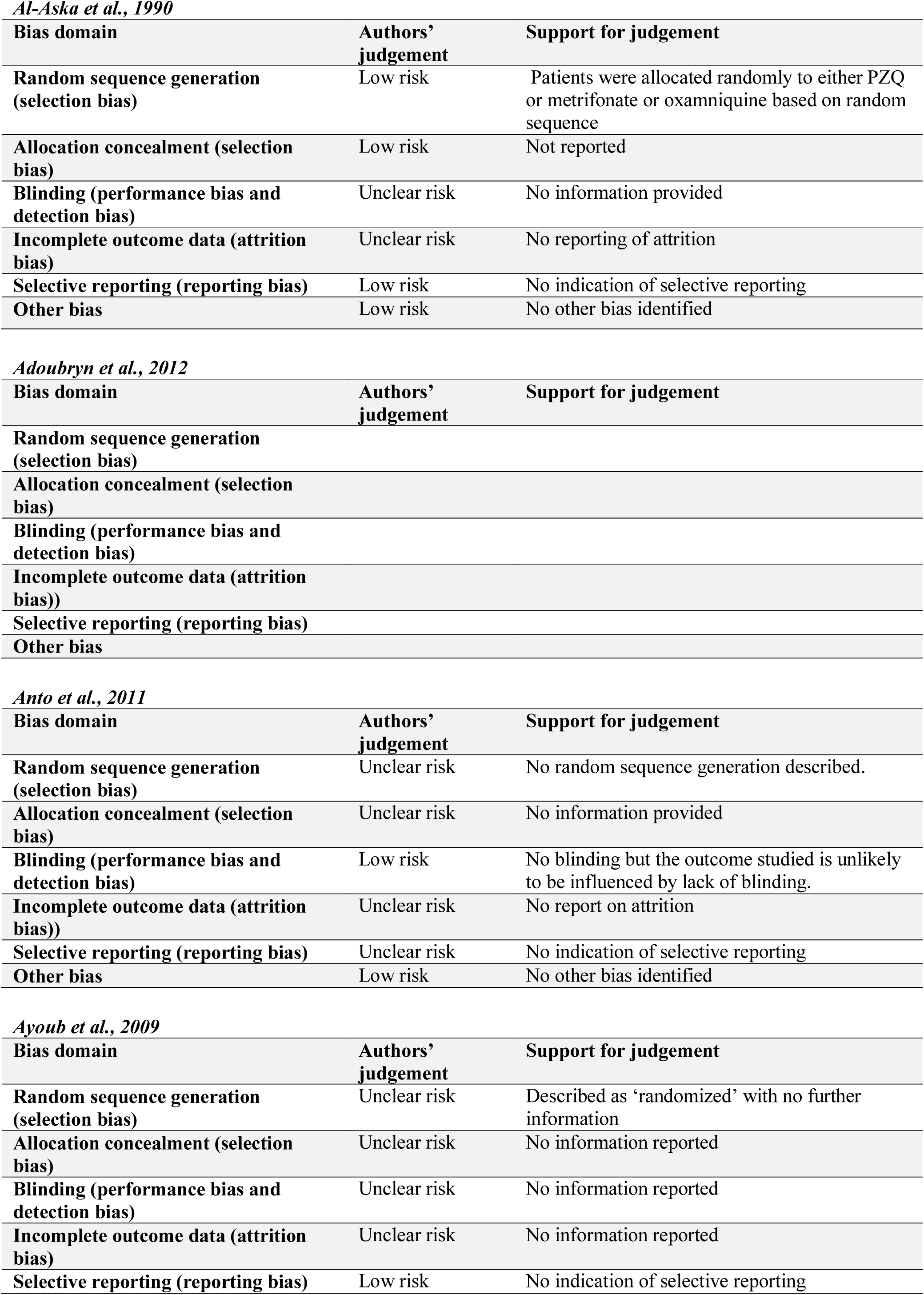
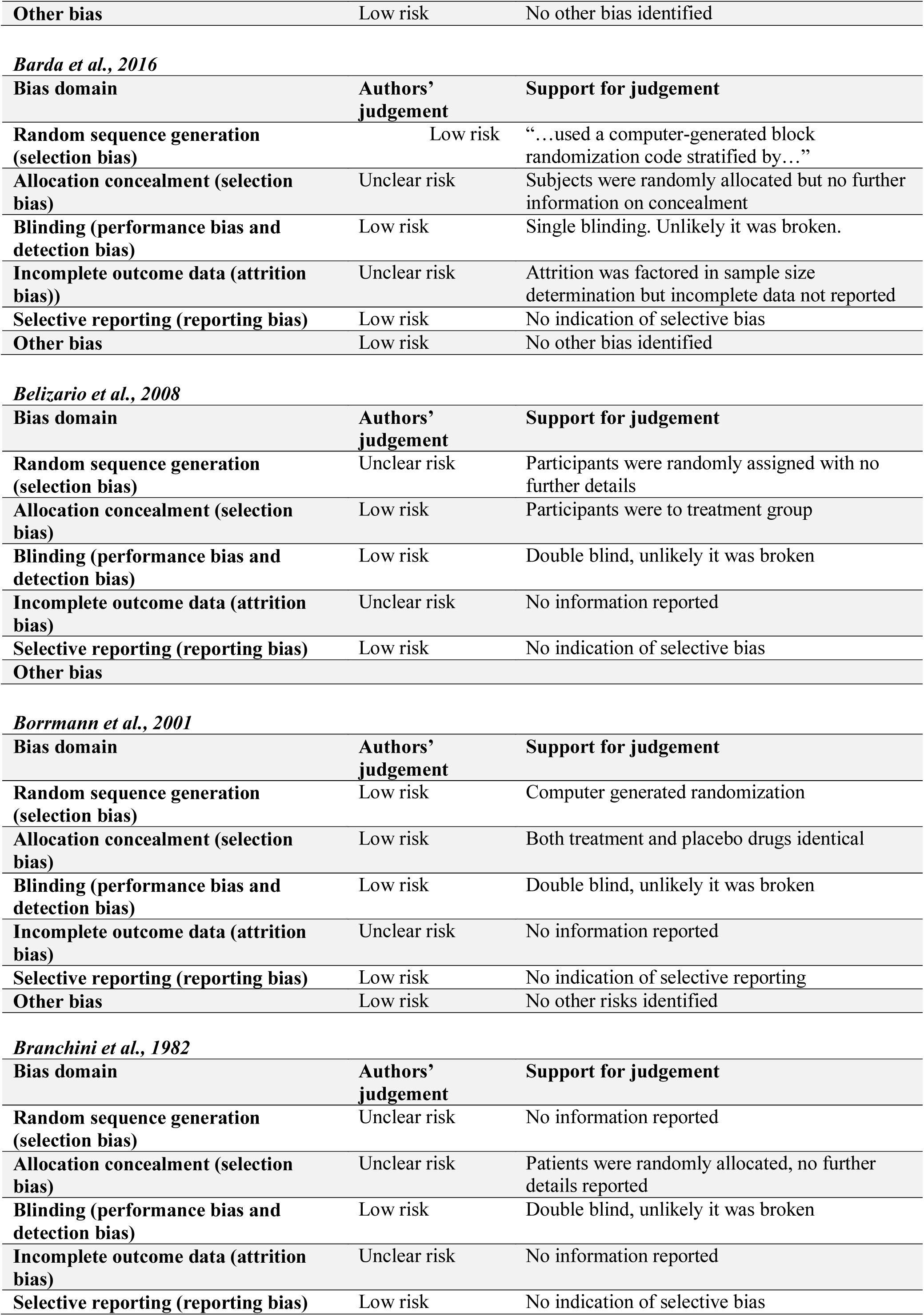
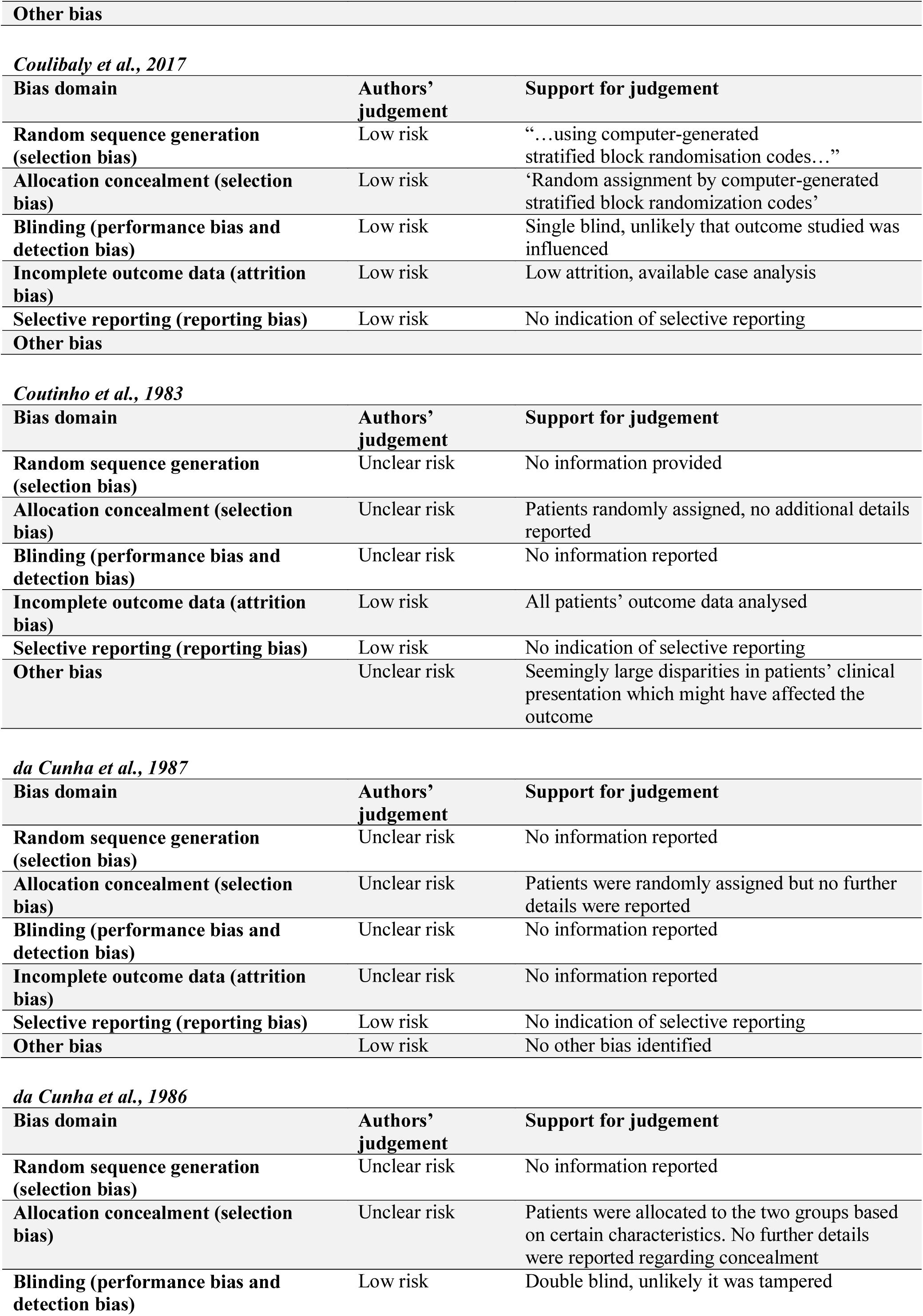
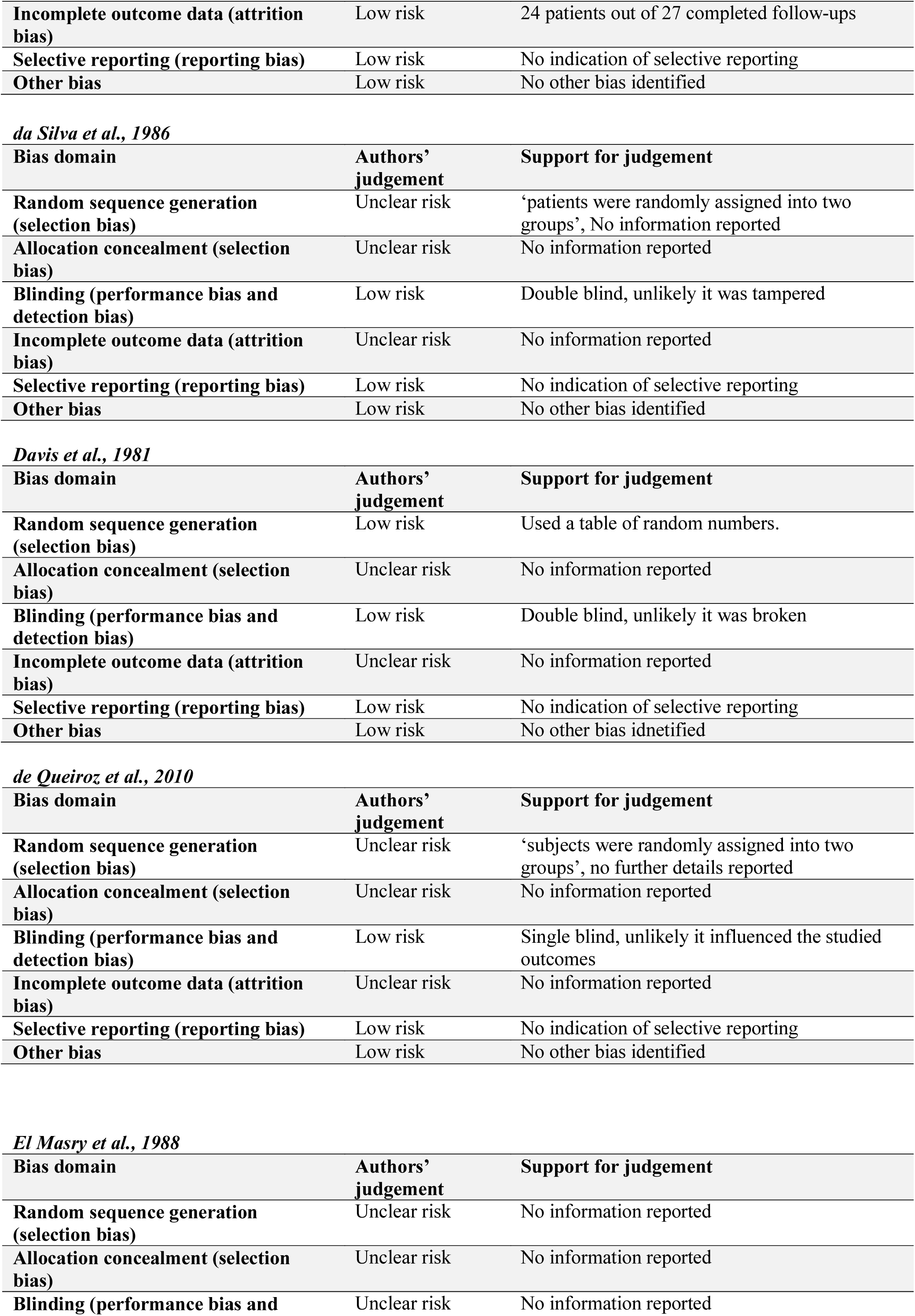
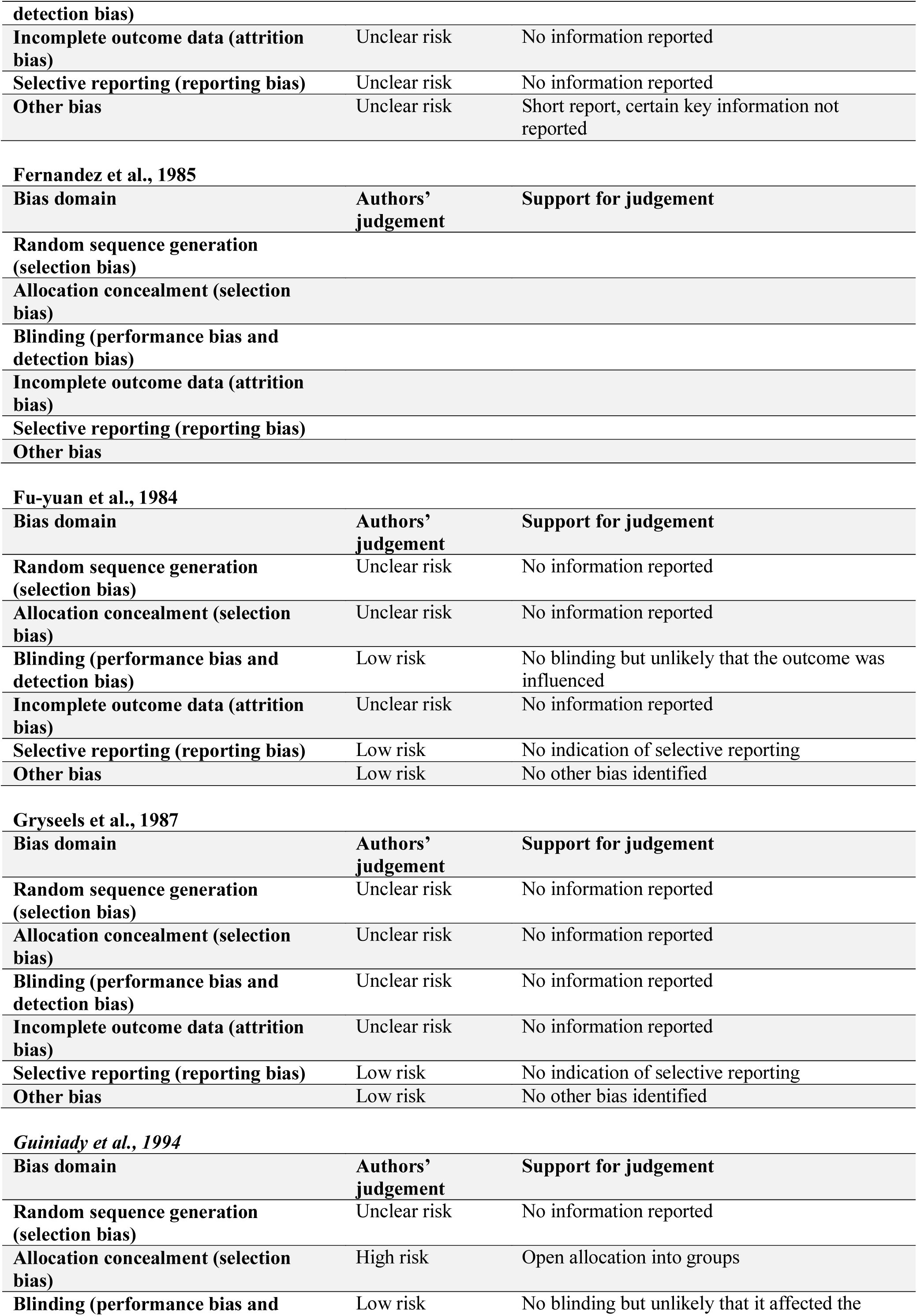
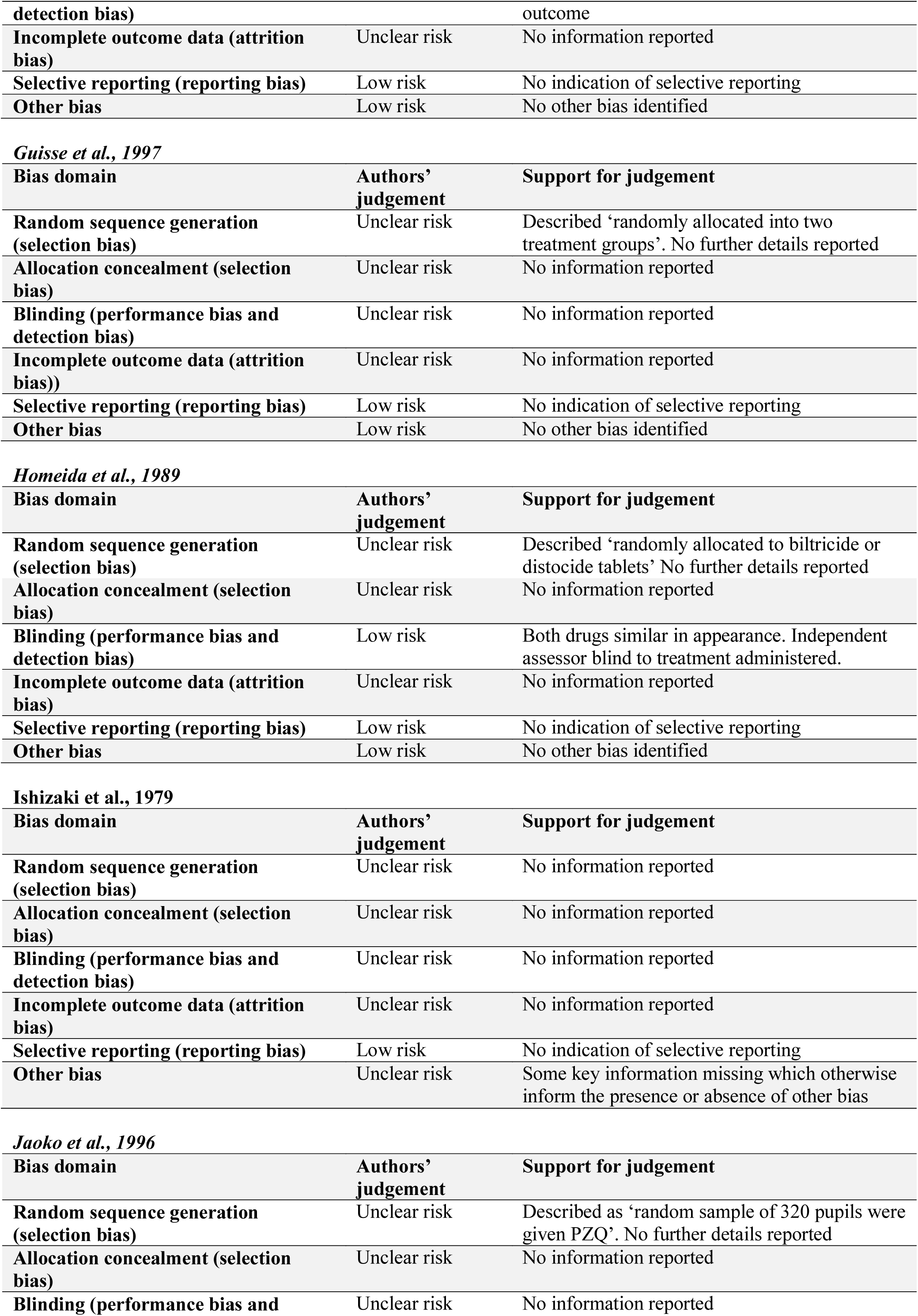
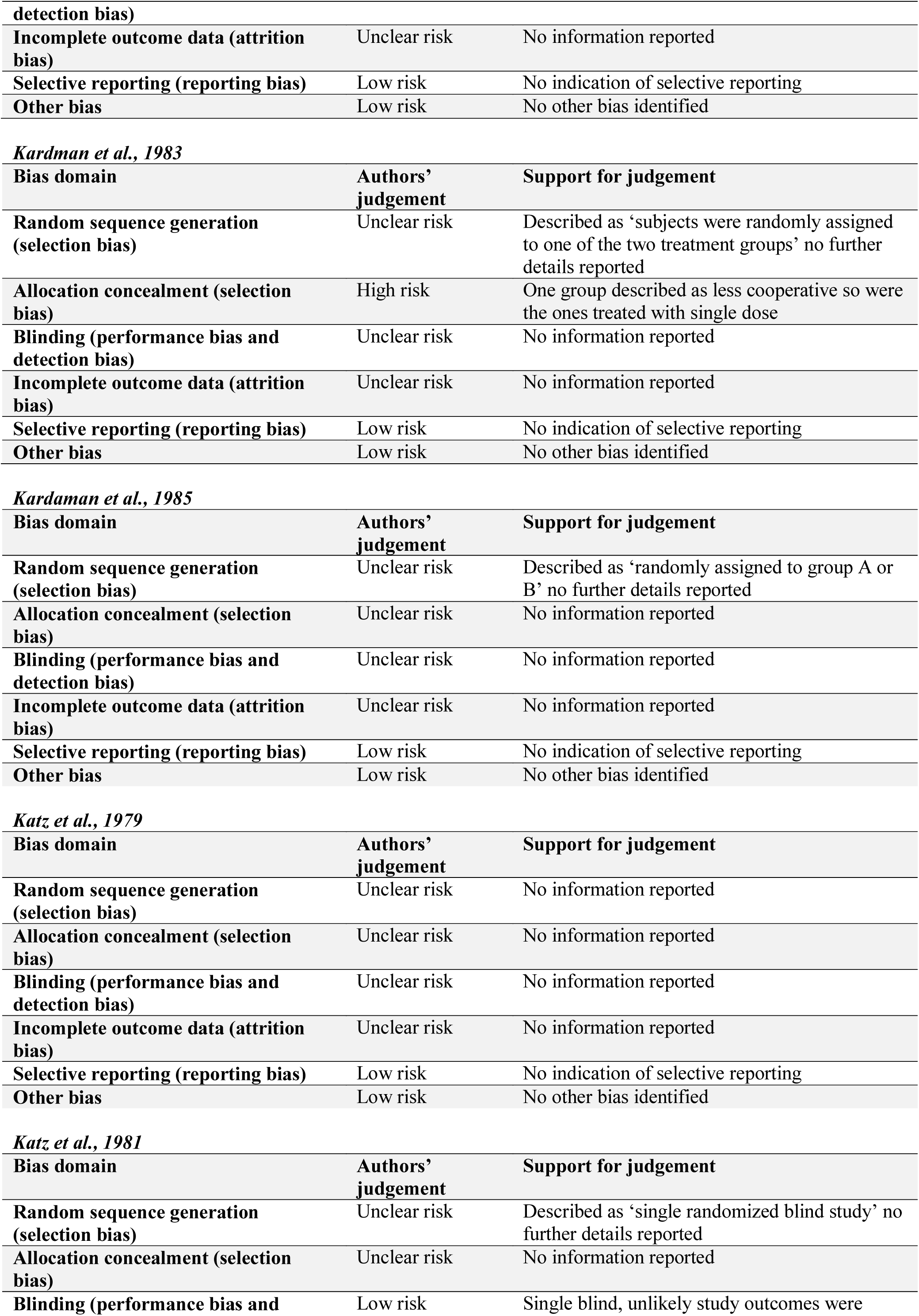
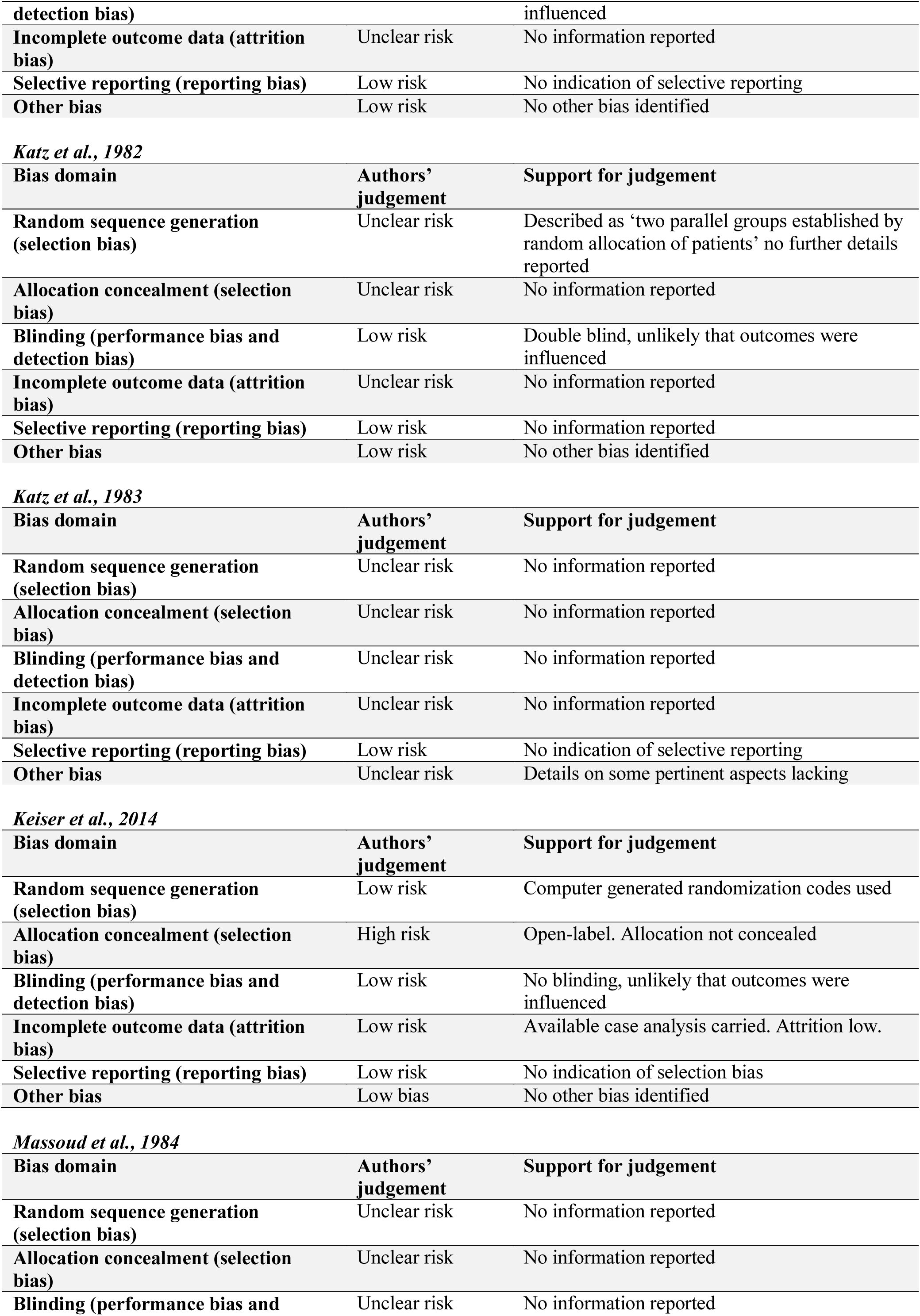
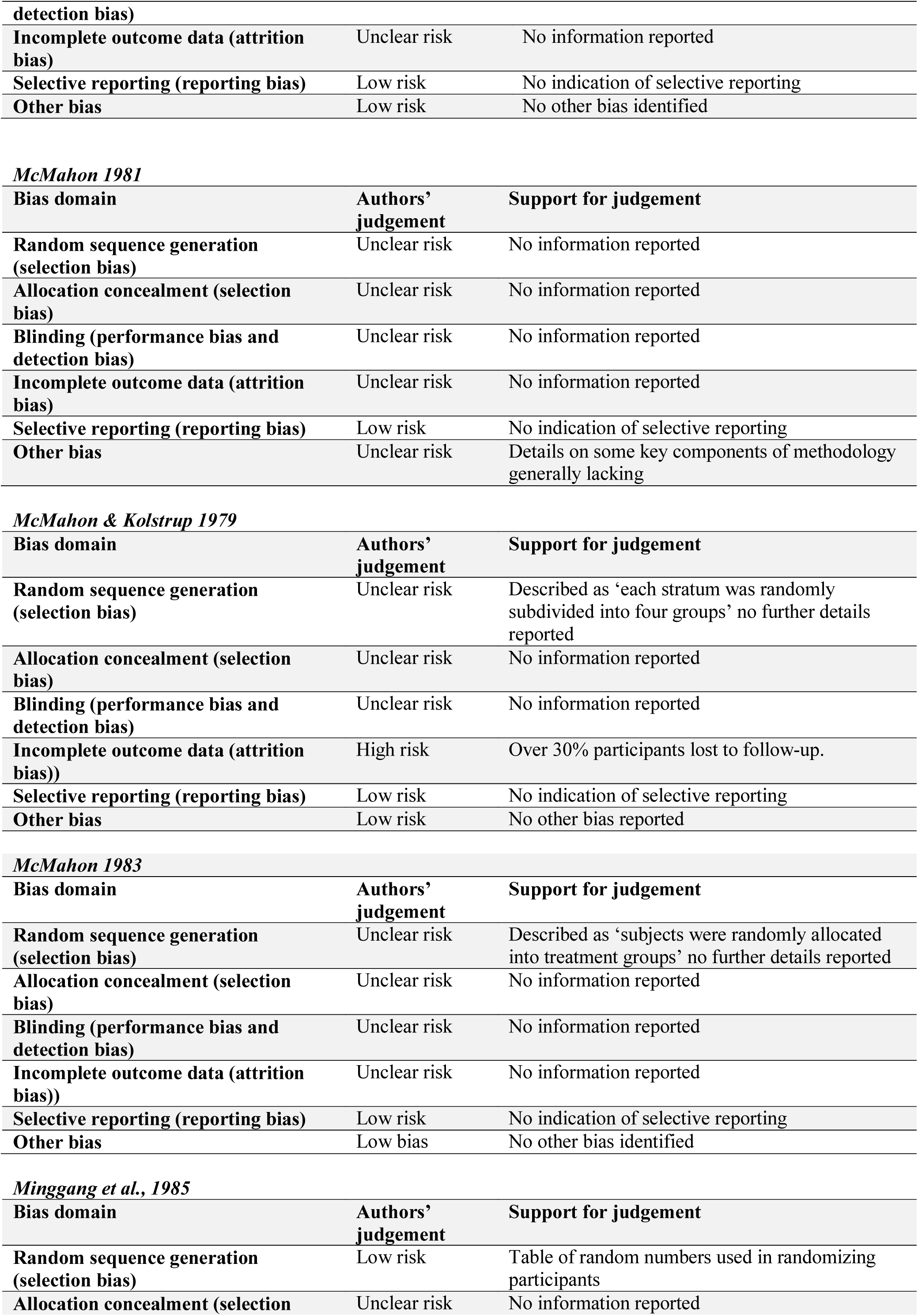
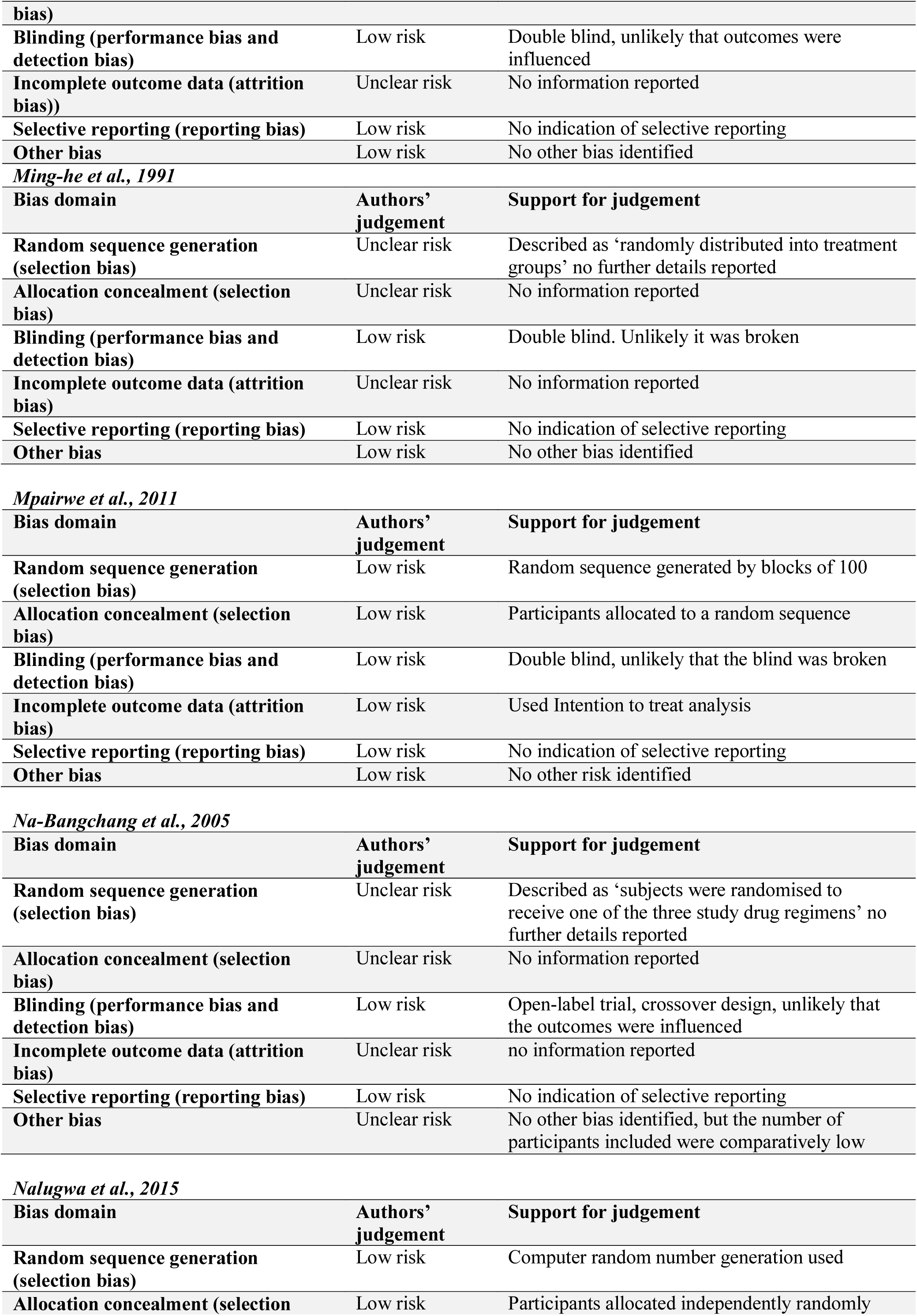
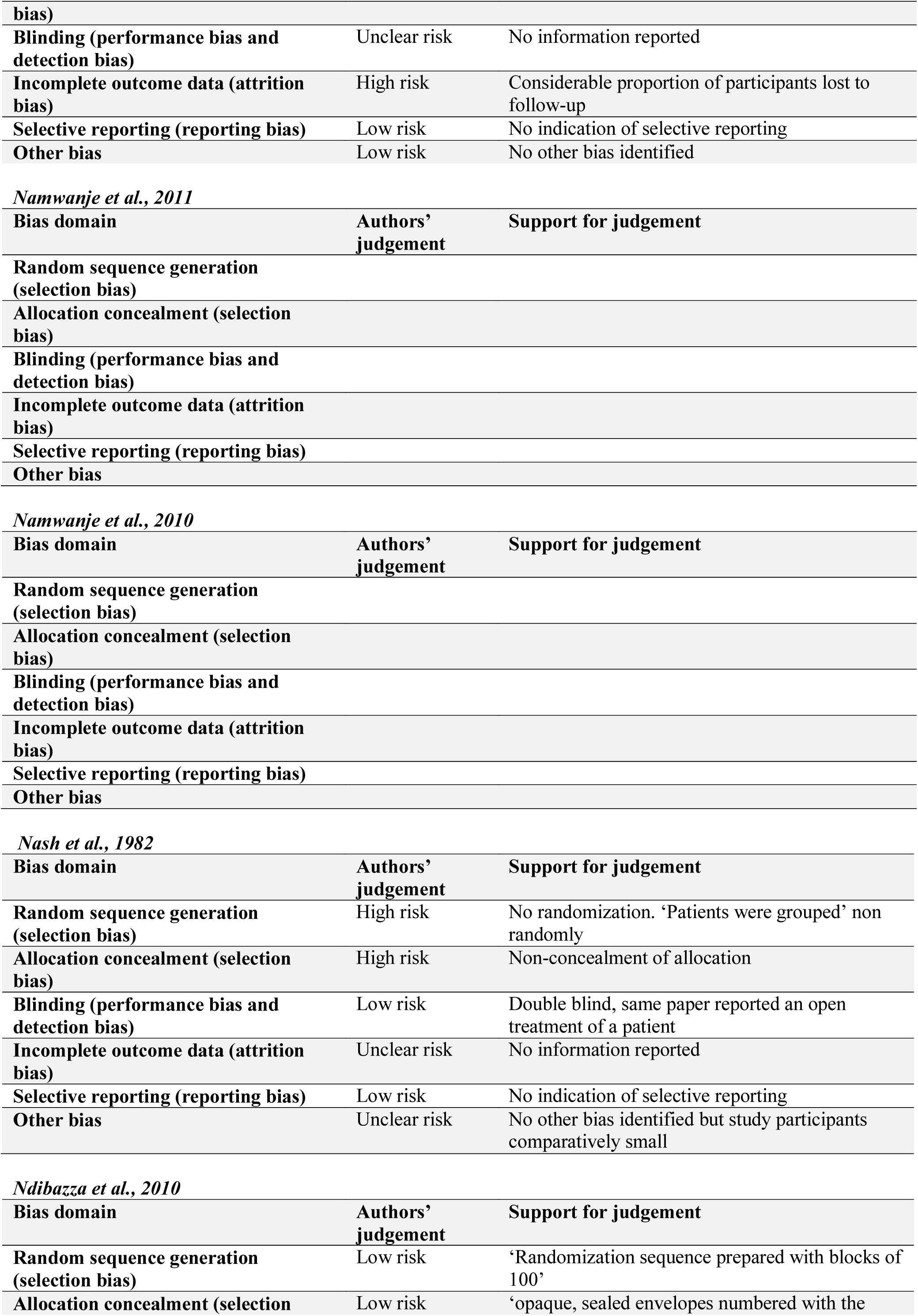
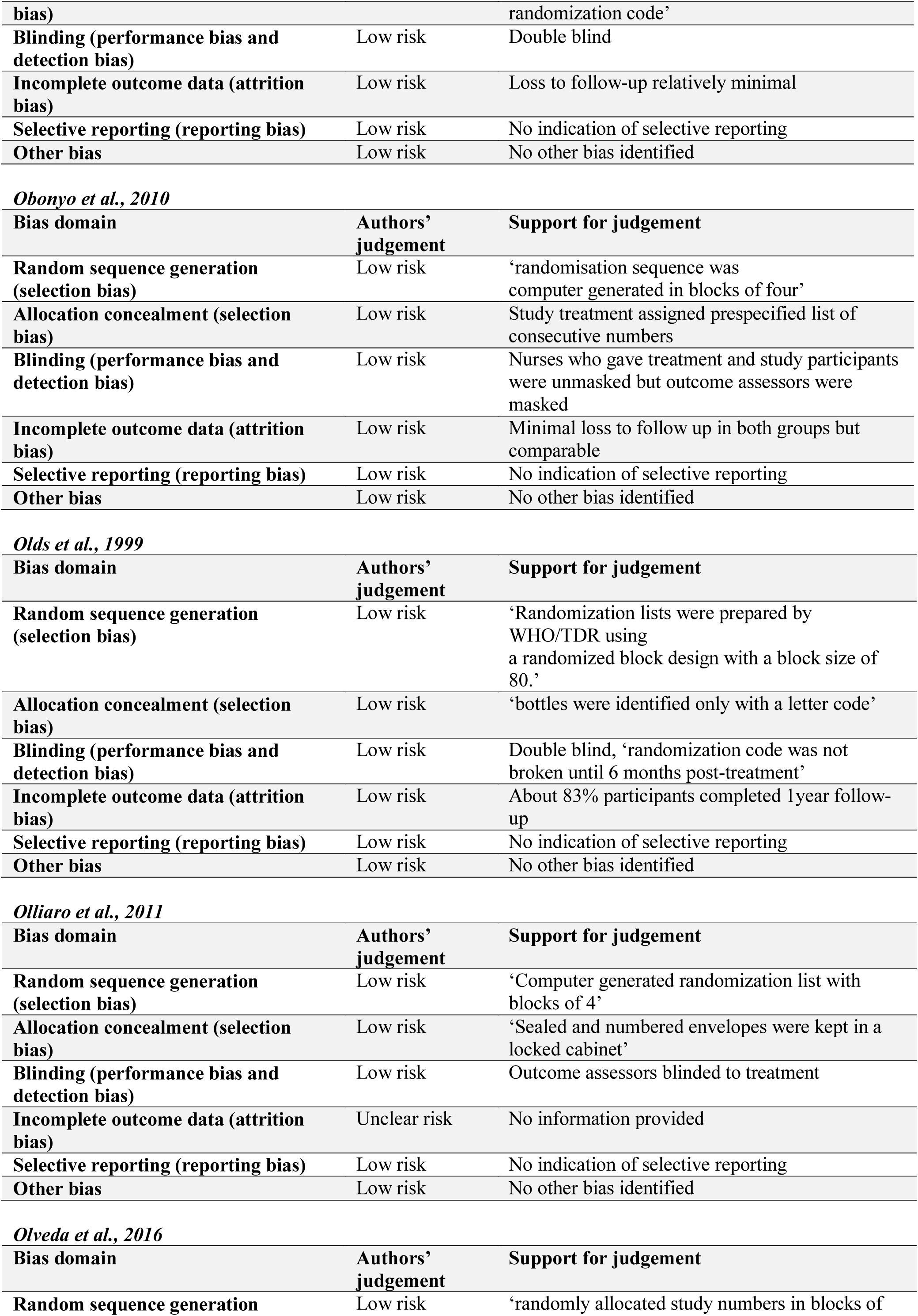
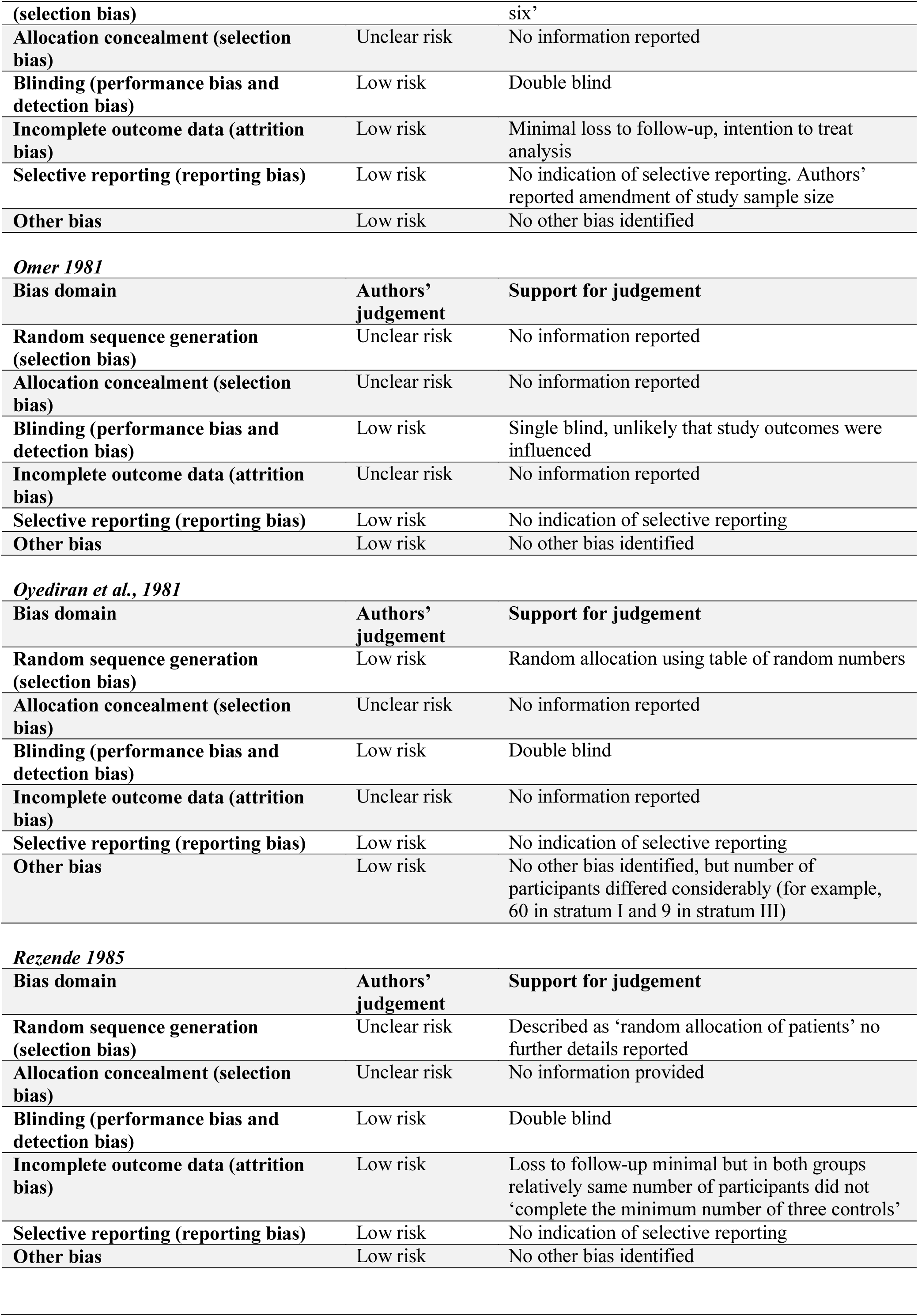
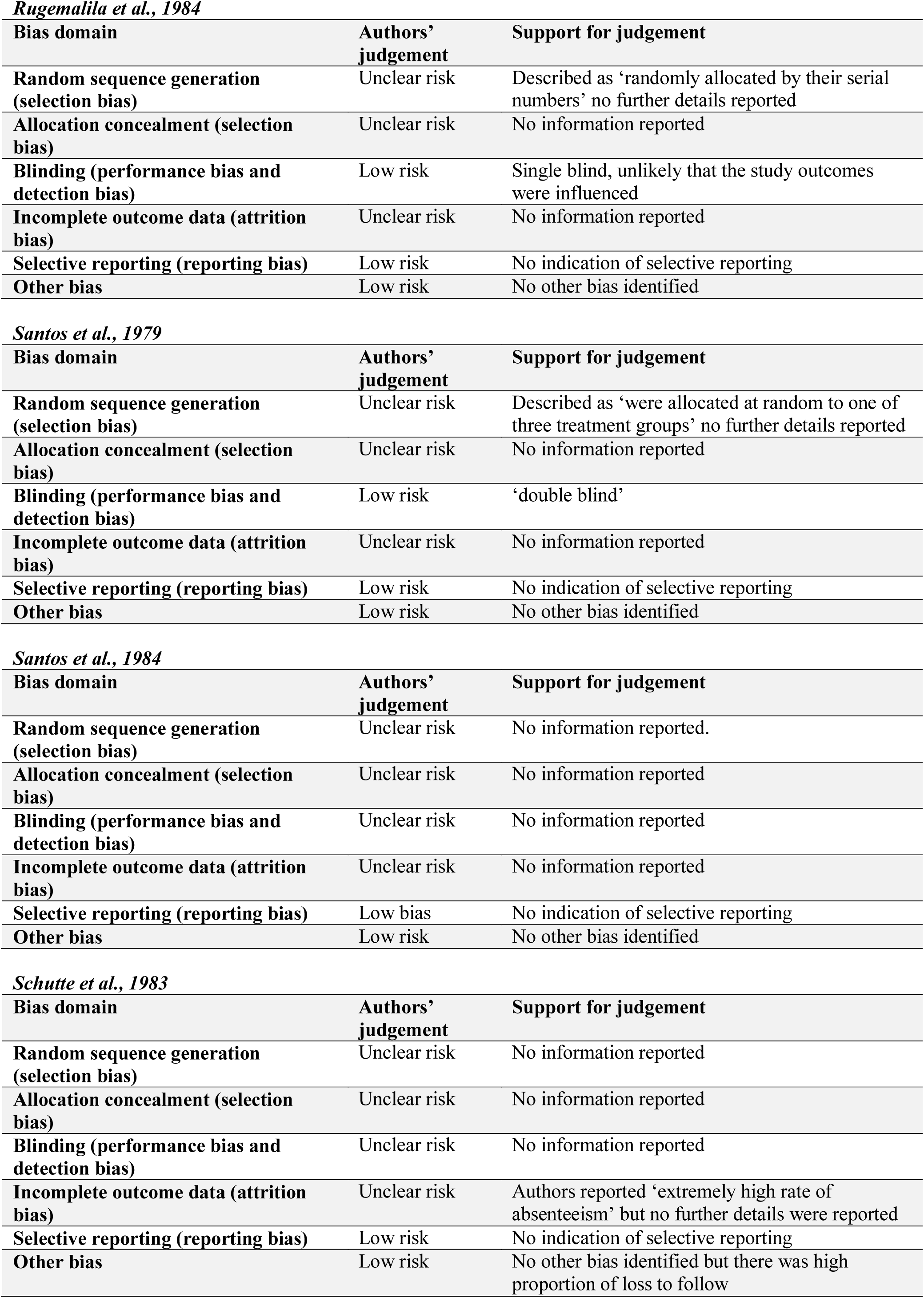
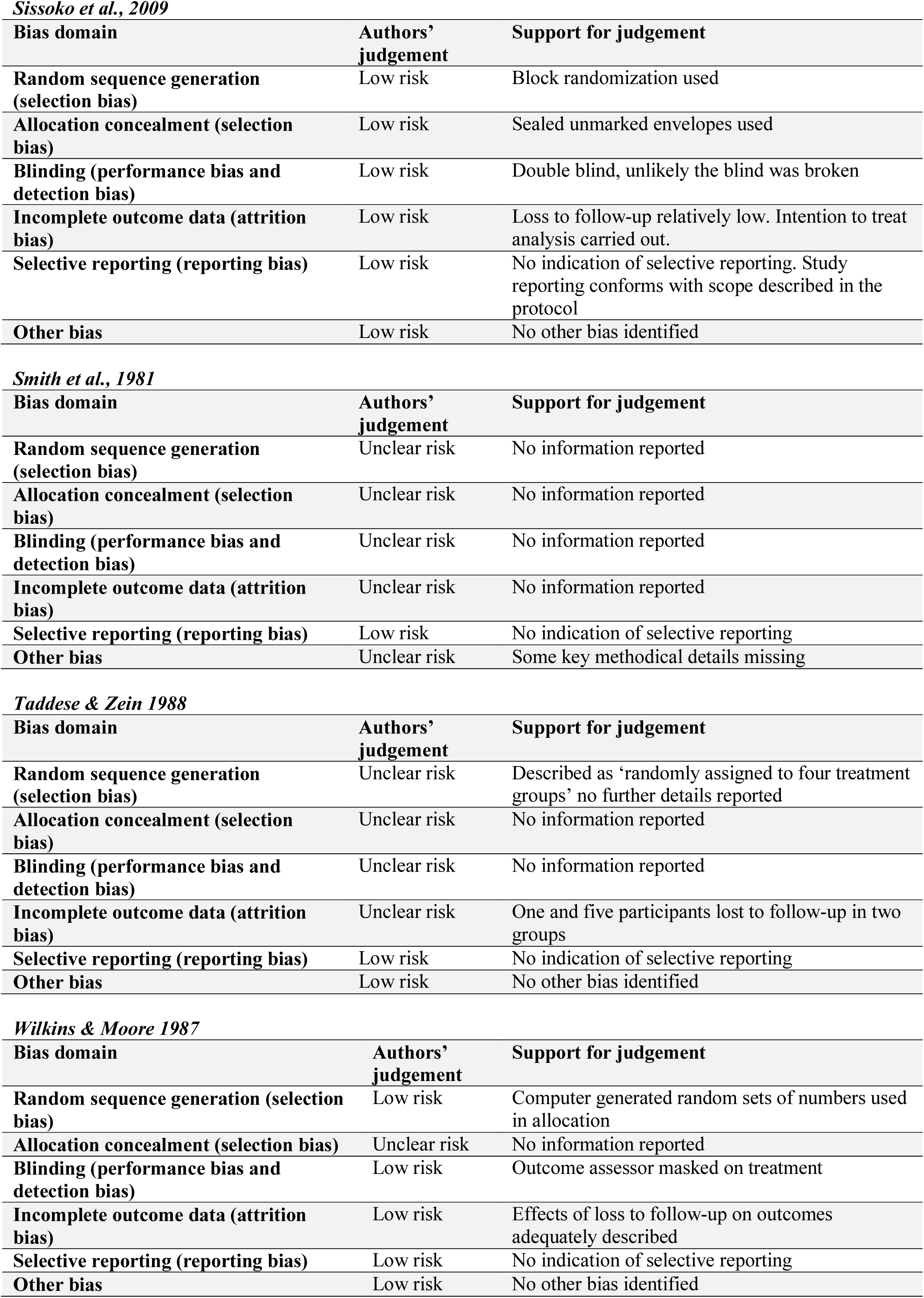
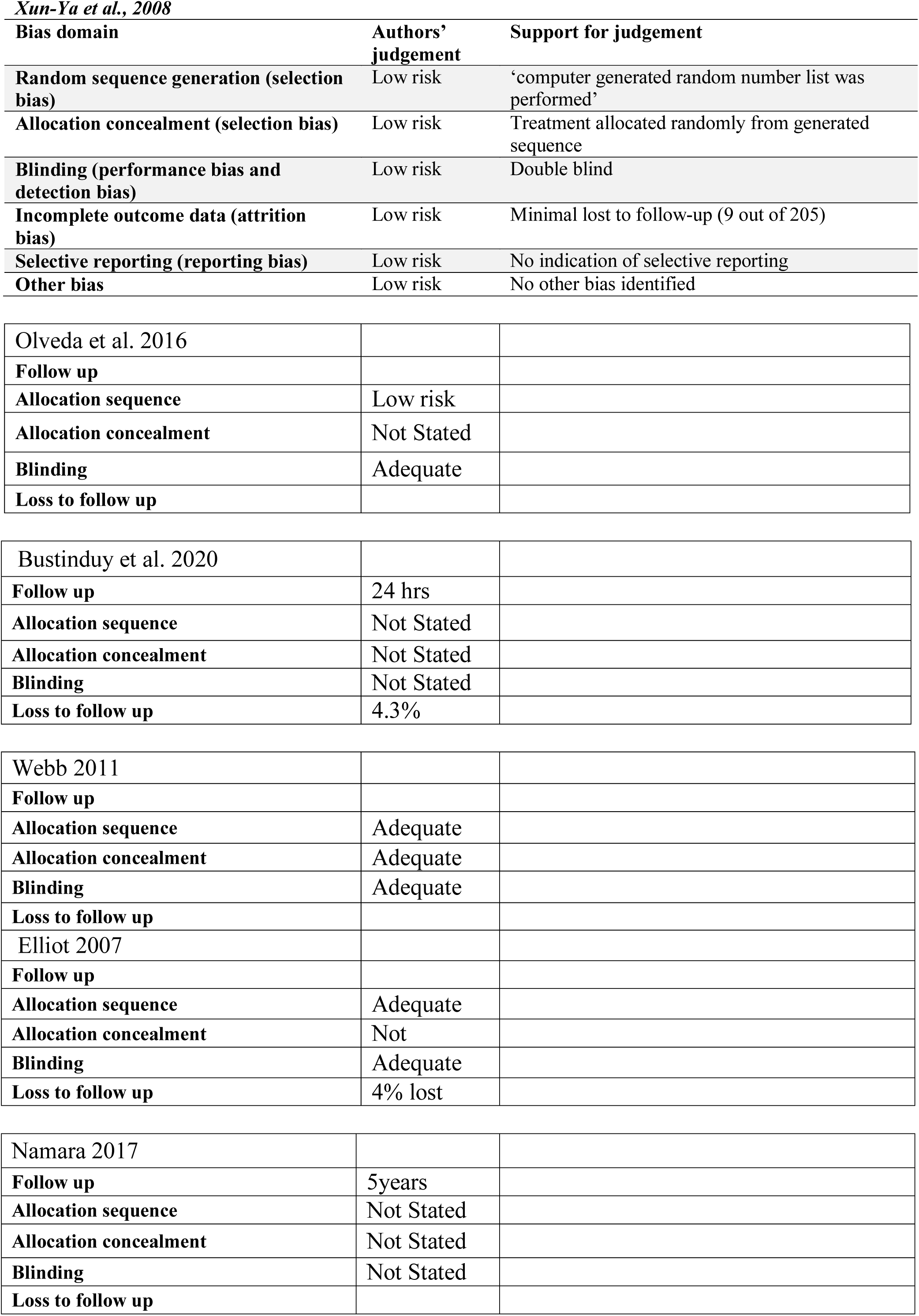
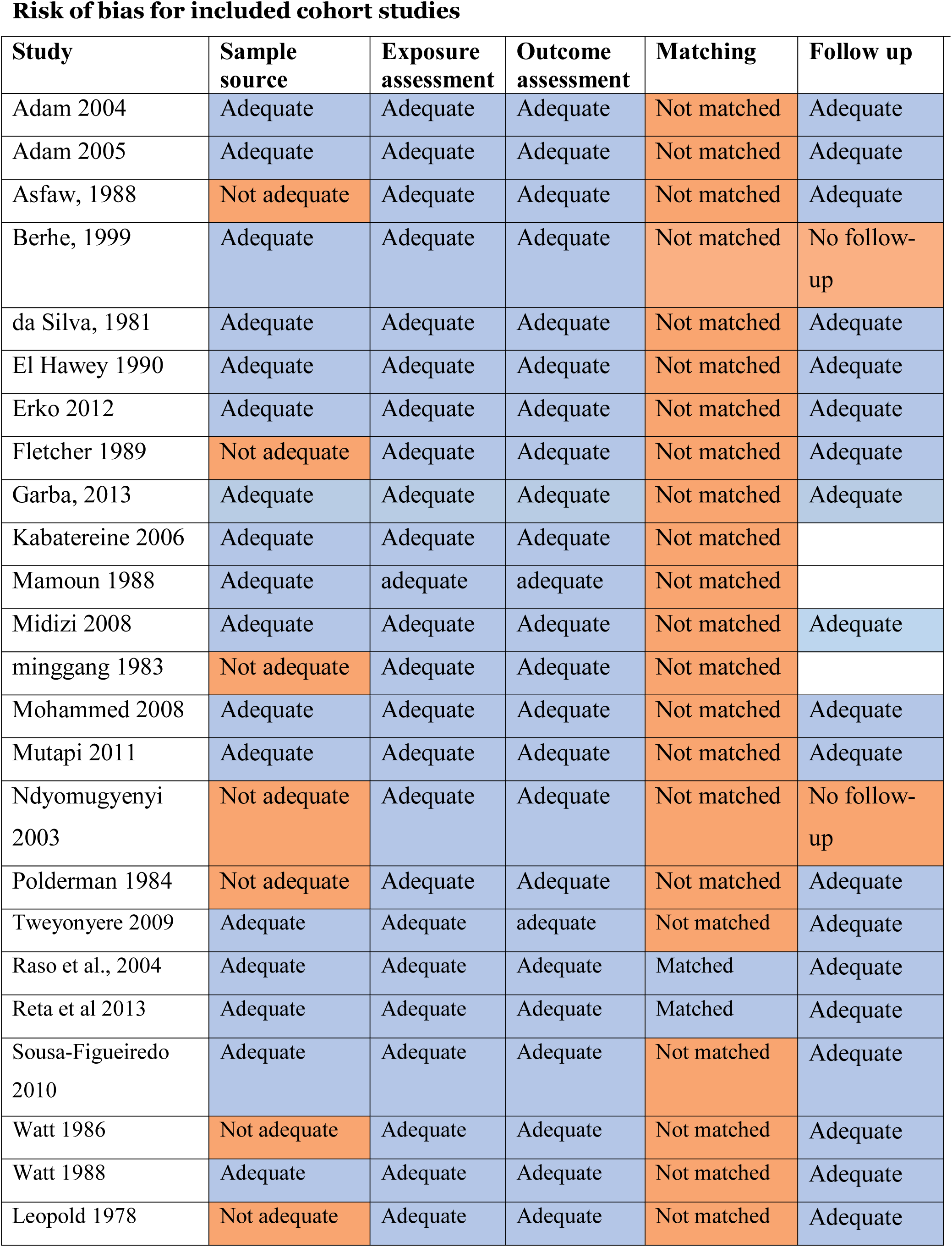

## References

1 Adam, I., Elwasila, E. T. and Homeida, M. (2004) Is praziquantel therapy safe during pregnancy? Transactions of the Royal Society of Tropical Medicine and Hygiene 98: 540–543.

2 Adam, I., Elwasila, E. and Homeida, M. (2005) Praziquantel for the treatment of schistosomiasis mansoni during pregnancy. Annals of Tropical Medicine and Parasitology 99(1): 37–40.

3 Adoubryn K.D., Kouadio-Yapo C.G., Ouhon J., Aka N.A.D., Bintto F., Assoumou A. Intestinal parasites in children in Biankouma, Ivory Coast (mountaineous western region): efficacy and safety of praziquantel and albendazole. Me decine et Sante Tropicales 2012; 22: 170–176.

4 Ahmed THE, Ali AA, Bassiouni GA, Omer SH and Shalaby MA (1988) Praziquantel in treatment of S.mansoni and S.haematobium infections. Journal of the Egyptian Society of Parasitology. 18(1)

5 Al-Aska AK, Al-Mofleh IA, Al-Rashed R, Hafez MA, Al-Nozha M, Abu-Aisha H, Al-Balla SR and Taha A. Praziquantel, Oxamniquine and Metrifonate in the treatment of Schistosomiasis in Riyadh. Annals of Saudi Medicine. 1990; 10(3):296–298

6 Alonso-Coello P, Schunemann HJ, Moberg J, et al. GRADE Evidence to Decision (EtD) frameworks: a systematic and transparent approach to making well informed healthcare choices. 1: Introduction. BMJ 2016; 353: i2016.

7 Amin MA, Swar M, Kardaman M , Elhussein D, Nouman G, Mahmoud A, Danso-Appiah A, Babiker A, Homeida M. Treatment of pre-school children under 6 years of age for schistosomiasis: safety, efficacy and acceptability of praziquantel. Sudan Journal of Medical Sciences. 2012; 7(2):67–79.

8 Anto F , Asoala V, Anyorigiya T, Oduro A, Adjuik M, Akweongo P, Aborigo R, Bimi L, Amankwa J, Hodgson A. Simultaneous administration of praziquantel, ivermectin and albendazole, in a community in rural northern Ghana endemic for schistosomiasis, onchocerciasis and lymphatic filariasis. Tropical Medicine & International Health, 2011; 16(9), 1112–1119.

9 Asfaw, Z., Wolde-Michael, T., & Wondimagegnehu, T. Assessment of side-effects of praziquantel in a trial treatment of *Schistosoma haematobium* infections in the Afar ethnic group of Ethiopia. Ethiopian medical journal, 1988; 26(2), 85–89.

10 Ayoub AM, Haider MM, Mamoun M, Gasim IG, Walid NE, Abd AA and Ishag A (2009) Artesunate plus sulfadoxine/pyrimethamine versus praziquantel in the treatment of Schistosoma mansoni in eastern Sudan. Trans. Of the Royal Society of Tropical Medicine and Hygiene. 32(7)

11 Azher, M, Faisal A, El-Kassitni, Wright SG, Mofti A. Exudative Polyserositis and Acute Respiratory Failure Following Praziquantel Therapy. CHEST, 1990 (98), 241. Copyright © 1990 by American College of Chest Physician. Downloaded from chestjournal.org Downloaded from on September 4, 2008.

12 Bada JL, Trevino B, Cabezos J. Convulsive seizures after treatment with praziquantel. British Medical Journal. 1988; (296):646.

13 Barda, B., Coulibaly, J. T., Puchkov, M., Huwyler, J., Hattendorf, J., & Keiser, J. Efficacy and safety of moxidectin, synriam, synriam-praziquantel versus praziquantel against *Schistosoma haematobium* and *S. mansoni* infections: a randomized, exploratory phase 2 trial. PLoS neglected tropical diseases, 2016; 10(9), e0005008.

14 Berhe, N., Gundersen, S. G., Abebe, F., Birrie, H., Medhin, G., & Gemetchu, T. Praziquantel side effects and efficacy related to *Schistosoma mansoni* egg loads and morbidity in primary school children in north-east Ethiopia. Acta tropica, 1999; 72(1), 53–63.

15 Bagheri H, Simiand E, Montastruc J-L, Magnaval J-F. Adverse reactions to anthelminthics. Annals of Pharmacotherapy. 2004; 38:383–8.

16 Belizario, V.Y., Amarillo, M.L.E., Martinez, R.M., Mallari, A.O and Tai, C.M.C (2007) Efficacy and safety of 40mg/kg and 60mg/kg single doses of praziquantel in the treatment of schistosomiasis. Journal of Pediatric Infectious Diseases 3(2008): 27–34

17 Bonate PL, Wang TL, Passier P, Bagchus W, Burt H, Lupfert C, et al. Extrapolation of praziquantel pharmacokinetics to a pediatric population: a cautionary tale. J Pharmacokinet Pharmacodyn. 2018;45(5):747–62. 10.1007/s10928-018-9601-1

18 Bossuyt P. DC, Deeks J., Hyde C., Leeflang M., Scholten R. . Chapter 11: Interpreting results and drawing conclusions. In: Deeks JJ, Bossuyt PM, Gatsonis C (editors), Cochrane Handbook for Systematic Reviews of Diagnostic Test Accuracy Version 0.9. 2013. http://methods.cochrane.org/sdt/sites/methods.cochrane.org.sdt/files/uploads/DTA%20Handbook%20Chapter%2011%20201312.pdf

19 Branchini ML, Pedro R de J, Dias LC, Deberaldini ER. Double-blind clinical trial comparing praziquantel with oxamniquine in the treatment of patients with schistosomiasis mansoni. Rev Inst Med Trop Sao Paulo, 1982;24(5):315–21.

20 Borrmann S, Szlezak N, Faucher J.F, Matsiegui PB, Neubauer R, Binder RK, Lell, B, Kremsner PG. Artesunate and praziquantel for the treatment of Schistosoma haematobium infections: a double-blind, randomized, placebo-controlled study. Journal of Infectious Diseases. 2001; 184,1363–1366.

21 Braae, U. C., Magnussen, P., Harrison, W., Ndawi, B., Lekule, F., & Johansen, M. V. (2016). Effect of National Schistosomiasis Control Programme on Taenia solium taeniosis and porcine cysticercosis in rural communities of Tanzania. Parasite epidemiology and control, 1(3), 245–251.

22 Branchini, M. L. M., Pedro, R. D. J., Dias, L. C., & Deberaldini, E. R. (1982). Double-blind clinical trial comparing praziquantel with oxamniquine in the treatment of patients with schistosomiasis mansoni. Rev. Inst. Med. Trop. Säo Paulo, 24(5), 315–21.

23 Bustinduy, A. L., Kolamunnage-Dona, R., Mirochnick, M. H., Capparelli, E. V., Tallo, V., Acosta, L. P., … & Hope, W. W. (2020). Population pharmacokinetics of praziquantel in pregnant and lactating Filipino women infected with Schistosoma japonicum. Antimicrobial agents and chemotherapy, 64(9), e00566–20.

24 Chen, M. G., Fu, S., Hua, X. J., & Wu, H. M. (1983). A retrospective survey on side effects of praziquantel among 25,693 cases of schistosomiasis japonica. The Southeast Asian journal of tropical medicine and public health, 14(4), 495–500.

25 Ben□Chetrit, E., Lachish, T., Mørch, K., Atias, D., Maguire, C., & Schwartz, E. (2015). Schistosomiasis in pregnant travelers: a case series. Journal of Travel Medicine, 22(2), 94–98

26 Chen, M. G., Fu, S., Hua, X. J., & Wu, H. M. (1983). A retrospective survey on side effects of praziquantel among 25,693 cases of schistosomiasis japonica. The Southeast Asian journal of tropical medicine and public health, 14(4), 495–500.

27 Colley DG, Secor WE. Immunology of human schistosomiasis. Parasite Immunol. 2014;36(8):347–57.

28 Coulibaly, J. T., Panic, G., Silué, K. D., Kova, J., Hattendorf, J., & Keiser, J. Efficacy and safety of praziquantel in preschool-aged and school-aged children infected with *Schistosoma mansoni:* a randomised controlled, parallel-group, dose-ranging, phase 2 trial. The Lancet Global Health. 2017; 5(7), e688–e698.

29 Coulibaly, J. T., N’Gbesso, Y. K., Knopp, S., Keiser, J., N’Goran, E. K., & Utzinger, J. (2012). Efficacy and Safety of Praziquantel in Preschool-Aged Children in an Area Co-Endemic for Schistosoma mansoni and S. haematobium. PLoS Neglected Tropical Diseases, 6(12). http://doi.org/10.1371/journal.pntd.0001917.

30 Coutinho, A., Domingues, A. L. C., Neves, J., Almeida, S. T. Treatment of Hepatosplenic Schistosomiasis mansoni with Praziquantel. Arzneim-Forsch./Drug Research. 1983; 33(*1*), Nr. 5(1983), pp787–791.

31 Creasey AM, Taylor P, and Thomas JEP. Dosage trial of a combination of oxamniquine and praziquantel in the treatment of schistosomiasis in Zimbabwean schoolchildren. The Central African Journal of Medicine. 1986; 32(7):165–7.

32 da Cunha, A. S., Cancado, J. R., Rezende, G. L. Therapeutical evaluation of different dose regimens of praziquantel in schistosomiasis mansoni, based on the quantitative oogram technique. Rev. Inst. Med. Trop. Sao Paulo. 1987; 295:295-304.

33 da Cunha, A. S., Pedrosa, R. C. Double-blind evaluation based on the quantitative technique, comparing praziquantel and oxamniquine in human schistosomiasis mansoni. Rev. Inst. Med. Trop. Sao Paulo. 1986; 28(5):337-351.

34 da Silva, L. C. D., Zeitune, J. M. R., Rosa-Eid, L. M. F., Lima, D. M. C., Antonelli, R. H., Christo, C. H., … & Carboni, A. D. C. Treatment of patients with schistosomiasis mansoni: a double-blind clinical trial comparing praziquantel with oxamniquine. Revista do Instituto de Medicina Tropical de São Paulo. 1986; 28(3), 174–180.

35 da Silva, L.C., Sette, H., Christo, C. H., Saez-Alquezar, A., Carneiro, C.R.W., Lacet., C.M., Ohtsuki, N. and Raia, S. Praziquantel in the treatment of the hepatosplenic form of *Shcistosomiasis mansoni*. Arzneimittelforschung. 1981;31(3a):601–3.

36 Davis, A., Biles, J. E., Ulrich, A. M. and Dixon, H. (1981). Tolerance and efficacy of praziquantel in phase IIA and II B therapeutic trials in Zambian patients. Arzneimittelforschung 31, 568–574.

37 De Clercq, D., Vercruysse, J., Kongs, A., Verle, P., Dompnier, J. P. and Faye, P. C.(2002). Efficacy of artesunate and praziquantel in *Schistosoma haematobium* infected schoolchildren. Acta Tropica 82, 61–66

38 de Queiroz, L. C., Drummond, S. C., de Matos, M. L. M., Paiva, M., Batista, T. S., Kansaon, A. Z., & Lambertucci, J. R. (2010). Comparative randomised trial of high and conventional doses of praziquantel in the treatment of schistosomiasis mansoni. Memórias do Instituto Oswaldo Cruz, 105(4), 445–448.

39 DerSimonian R, Laird N. Meta-analysis in clinical trials revisited. Contemp Clin Trials [Internet]. 2015;45:139–45. Available from: http://dx.doi.org/10.1016/j.cct.2015.09.002.

40 Ejezie, C., & Okeke, G. C. Chemotherapy in the control of urinary schistosomiasis in Nigeria. The Journal of Tropical Medicine and Hygiene, 1987; 90(3), 149–151.

41 El-Alamy, M. A., Habib, M. A., McNeeley, D. F., & Cline, B. L. (1981). Preliminary results of chemotherapy using praziquantel on a large scale in Qalyub Bilharziasis Project where simultaneous infection with S. mansoni and S. haematobium exists. Arzneimittelforschung, 31, 612–615.

42 El-Hawey, A.M., Massoud, A.M., El-Rakieby, A., Rozeik, M.S. and Nassar, M.O. Side effects of praziquantel in bilharzial children on a field level. Journal of the Egyptian Society of Parasitology. 1990; 20(2):599–605.

43 Elliott, A. M., Kizza, M., Quigley, M. A., Ndibazza, J., Nampijja, M., Muhangi, L., … & Whitwortha, J. A. (2007). The impact of helminths on the response to immunization and on the incidence of infection and disease in childhood in Uganda: design of a randomized, double-blind, placebo-controlled, factorial trial of deworming interventions delivered in pregnancy and early childhood [ISRCTN32849447]. Clinical trials, 4(1), 42–57.

44 El Masry, N.A., Bassily, S. and Farid, Z (1988). A comparison of the efficacy of various regimens of praziquantel for the treatment of schistosomiasis. Transactions of the Royal Society of Tropical Medicine and Hygiene, 82: 719–720

45 Erko, B., Degarege, A., Tadesse, K., Mathiwos, A., & Legesse, M. Efficacy and side effects of praziquantel in the treatment of Schistosomiasis mansoni in schoolchildren in Shesha Kekele Elementary School, Wondo Genet, Southern Ethiopia. Asian Pacific Journal of Tropical Biomedicine, 2(3), 235–239. http://doi.org/10.1016/S2221-1691(12)60049-5.

46 Ezeamama AE, Friedman JF, Acosta LP, Bellinger DC, Langdon GC, Manalo DL, Olveda RM, Kurtis JD, Stephen T McGarvey ST. Helminth infection and cognitive impairment among Filipino children. American Journal of Tropical Medicine and Hygiene. 2005;72,540–548.

47 Farid Z, Woody J, Kamal M. Praziquantel and acute urban schistosomiasis. Tropical and Geographical Medicine. 1988;41(2):172.

48 Fernandes P and Oliveira CC. Estudo comparative da eficacia do praziquantel, em dois esquemas posologicos, e da oxaminiquina no tratamento da esquitossomose mansonica. Farmacologica Clinica. 1985; 93(5-6), 389-393.

49 Ferrari, M. L. A., Coelho, P. M. Z., Antunes, C. M. F., Tavares, C. A. P., & Da Cunha, A. S. (2003). Efficacy of oxamniquine and praziquantel in the treatment of Schistosoma mansoni infection: a controlled trial. Bulletin of the World Health Organization, 81, 190–196.

50 Fletcher M & Teklehaimanot A. Transactions of the Royal Society of Tropical Medicine and Hygiene. 1989; 83, 793–797.

51 Fonseca CT, Oliveira SC, Alves CC. Eliminating Schistosomes through vaccination: what are the best immune weapons? Frontiers in Immunology. 2015;6:95.

52 Frohberg H, Schulze Schencking H. Toxicological profile of praziquantel, a new drug against cestode and schistosome infections, as compared to some other schistosomicides. Arzneimittelforschung. 1981;31(3a):555–65.

53 Fu-Yuan, et al., Further experience with Praziquantel in Schistosoma japonicum infection. Report of 716 cases. Chinese Medical Journal. 1984; 97(1); 47–52.

54 Garba, A., Lamine, M. S., Barkiré, N., Djibo, A., Sofo, B., Gouvras, A. N., … & Utzinger, J. (2013). Efficacy and safety of two closely spaced doses of praziquantel against Schistosoma haematobium and S. mansoni and re-infection patterns in school-aged children in Niger. Acta tropica, 128(2), 334–344.

55 Groning, E., Bakathir, H., Salem, A., Albert, L., & Fernández, R. (1985). Effectiveness and tolerance of praziquantel in schistosomiasis. Revista cubana de medicina tropical, 37(2), 215–219.

56 Gryseels B, Nkulikyinka l, Coosemans MH. Field trials of praziquantel and oxamniquine for the treatment of schistosomiasis mansoni in Burundi. Transactions of the Royal Society of Tropical Medicine and Hygiene (1987) 81, 641–644.

57 el Guiniady MA, el Touny MA, Abdel-Bary MA, Abdel-Fatah SA, Metwally A. Clinical and Pharmacokinetic study of praziquantel in Egyptian Schistosomiasis patients with and without liver cell failure. American Journal of Tropical Medicine and Hygiene. 1994; 51 (6) 809 – 818.

58 Guisse F, Polman K, Stelma FF, Mbaye A, Talla I, Niang M, Deelder MA, Ndir O, Gryseels B. Therapeutic evaluation of Two different dose regimens of praziquantel in a recent Schistosoma mansoni focus in Northern Senegal. American Journal of Tropical Medicine and Hygiene 1997; 56(5):511–514.

59 Higgins JPT, Savović J, Page MJ, Elbers RG, Sterne JAC. Chapter 8: Assessing risk of bias in a randomized trial. In: Higgins JPT, Thomas J, Chandler J, Cumpston M, Li T, Page MJ, Welch VA (editors). Cochrane Handbook for Systematic Reviews of Interventions version 6.2 (updated February 2021). Cochrane, 2021. Available from www.training.cochrane.org/handbook.

60 Homeida MMA, Sulaiman SM, Ali HM, Bennet JL. Tolerance of two brands of praziquantel. The Lancet. 1989; 391.

61 Homeida, M.M.A., El Tom, I.A., Sulaiman, S.M., Daffalla, A.A. and Bennett, J.L. Efficacy and tolerance of praziquantel in patients with Schistosoma mansoni infection and Symmers’ fibrosis: a field study in the Sudan. American Journal of Tropical Medicine and Hygiene. 1988; 38(3):496–498.

62 Yan, D. E. N. G., Ya-lan, Z. H. A. N. G., Su-hua, L. I., Wei-qi, C. H. E. N., Xi-meng, L. I. N., Xi, C. H. E. N., … & Hong-wei, Z. H. A. N. G. (2018). A KAP survey on taeniasis and cysticercosis in Fangcheng County of Henan Province in 2016. CHINESE JOURNAL OF PARASITOLOGY AND PARASITIC DISEASES, 36(3), 19.

63 Hoy D, Brooks P, Woolf A, et al. Assessing risk of bias in prevalence studies: modification of an existing tool and evidence of interrater agreement. Journal of Clinical Epidemiology 2012; 65:934–9.

64 Ishizaki, T., Kamo, E., & Boehme, K. (1979). Double-blind studies of tolerance to praziquantel in Japanese patients with Schistosoma japonicum infections. Bulletin of the World Health Organization, 57(5), 787.

65 Jaoko WG, Muchemi G, Oguya FO. Praziquantel side effects during treatment of Schistosoma mansoni infected pupils in Kibwezi, Kenya. East African Medical Journal. 1996;73(8):499–501.

66 Leopold, G., Ungethum, W., Groll, E., Diekmann, H. W., Nowak, H. and Wegner, D. H.(1978). Clinical pharmacology in normal volunteers of praziquantel, a new drug against schistosomes and cestodes. An example of a complex study covering both tolerance and pharmacokinetics. European Journal of Clinical Pharmacology 14, 281–291

67 Kabatereine, N. B., Tukahebwa, E., Kazibwe, F., Namwangye, H., Zaramba, S., Brooker, S. & Fenwick, A. Progress towards countrywide control of schistosomiasis and soil-transmitted helminthiasis in Uganda. Transactions of the Royal Society of Tropical Medicine and Hygiene, 2006; 100(3), 208–215.

68 Kabatereine NB, Kemijumbi J, Ouma JH, Sturrock RF, Butterworth AE, Maden H, Ornbjerg N, Dune DW and Vennervald BJ Efficacy and side effects of praziquantel treatment in a highly endemic Schistosoma mansoni focus at Lake Albert, Uganda. Transactions of the Royal Society of Tropical Medicine and Hygiene. 2003; 97, 599-603.

69 Kardaman, M. W., Amin, M. A., Fenwick, A., Cheesmond, A. K., & Dixon, H. G. A field trial using praziquantel (BiltricideR) to treat *Schistosoma mansoni* and *Schistosoma haematobium* infection in Gezira, Sudan. Annals of Tropical Medicine & Parasitology, 1983; 77(3), 297–304.

70 Kardaman, M. W., Fenwick, A., El Igail, A. B., El Tayeb, M., Daffalla, A. A., & Dixon, H. G. Treatment with praziquantel of schoolchildren with concurrent *Schistosoma mansoni* and *S. haematobium* infections in Gezira, Sudan. The Journal of tropical medicine and hygiene, 1985; 88(2), 105–109.

71 Katz, N., Rocha, R. S., & Chaves, A (a). Preliminary trials with praziquantel in human infections due to *Schistosoma mansoni*. Bulletin of the World Health Organization, 1979;57(5),781–5.

72 72 Katz N (b). Preliminary trials with praziquantel in human infections due to *Schistosoma mansoni*. Bulletin of the World Health Organization 1979;57(5):781 5.

73 Katz, N., Rocha, R. S., & Chaves, A. Clinical trials with praziquantel in schistosomiasis mansoni. Revista do Instituto de Medicina Tropical de São Paulo. 1981; 23(2), 72–78.

74 Katz N, Rocha RS. Double-blind clinical trial comparing praziquantel with oxamniquine in *Schistosomiasis mansoni*. Revista do Instituto de Medicina Tropical de São Paulo. 1982; 24(5):310–314.

75 Katz N, Rocha RS, Lambertucci JR, Greco DB, Pedroso ER, Rocha MO, Flan S. Clinical trial with oxamniquine and praziquantel in the acute and chronic phases of Schistosomiasis mansoni. Revista do Instituto de Medicina Tropical de São Paulo. 1983;25(4):173–7.

76 Katz N, Rocha RS, Chaves A. Clinical trials with praziquantel in schistosomiasis mansoni. Revista do Instituto de Medicina Tropical de São Paulo. 1981;23(2):72–8.

77 Keiser, J., Silue JD, K. D., Adiossan, L. K., N’Guessan, N. A., Monsan, N., Utzinger, J., N’Goran, E. K. Praziquantel, Mefloquine-Praziquantel, and Mefloquine-Artesunate-Praziquantel against Schistosoma haematobium: A Randomized, Exploratory, Open-Label Trial. PLoS Neglected Tropical Diseases, 2014; 8(7).

78 Keiser J, N’Guessan NA, Adoubryn KD, Silue KD, Vounatsou P, Hatz C, Utzinger J and N’Goran EK. Efficacy and safety of mefloquine, artesunate, mefloquine-artesunate and praziquantel against Schistosoma haematobium: randomized, exploratory open-label trial Clinical Infectious Diseases. 2010; 50(9)1205–1213.

79 King CH, Sutherland LJ, Bertsch D. Systematic review and meta-analysis of the impact of chemical-based mollusciciding for control of Schistosoma mansoni and S. haematobium transmission. PLoS Neglected Tropical Diseases. 2015; 9(12):e0004290.

80 Lo NC, Gurarie D, Yoon N, Coulibaly JT, Bendavid E, Andrews JR, et al. Impact and cost-effectiveness of snail control to achieve disease control targets for schistosomiasis. Proceedings of the National Academy of Sciences of the United States of America. 2018; 115(4):E584–E591.

81 Leopold, G., Ungethum, W., Groll, E., Diekmann, H.W., Nowak, H. and Wegner, D.H.G Clinical pharmacology in normal volunteers of praziquantel, a new drug against schistosomes and cestodes. European Journal of Clinical Pharmacology. 1978; 14, 281–291.

82 Massoud, A.A.E., El Kholy, A.M and Anwar, W.A. Assessment of efficacy of praziquantel against schistosoma mansoni infection. Journal of Tropical Medicine and Hygiene. 1984; 87, 119–121.

83 McMahon JE. A comparative trial of praziquantel, metrifonate and niridazole against *Schistosoma haematobium*. Annals of Tropical Medicine and Parasitology. 1983;77(2):139–42.

84 McMahon JE. Observations on Praziquantel against Schistosoma haematobium. Arzneimittelforschung. 1981;31(3a):579–80.

85 Niger MDA. Mass drug administration for the control of S. haematobium infection in Niger (unpublished data).

86 McMahon JE, Kolstrup N. Praziquantel: a new schistosomicide against *Schistosoma haematobium*. British Medical Journal 1979;2(6202):1396–8.

87 Midzi, N., Sangweme, D., Zinyowera, S., Mapingure, M. P., Brouwer, K. C., Kumar, N. & Mduluza, T. (2008). Efficacy and side effects of praziquantel treatment against *Schistosoma haematobium* infection among primary school children in Zimbabwe. Transactions of the Royal Society of Tropical Medicine and Hygiene, 102(8), 759–766.

88 Minggang, C., Xiangjin, H., Mingjie, W., Rongji, X., Changbao, Y and Shoubai, J. Dose finding double-blind clinical trial with praziquantel in schistosomiasis japonica patients. Southeast Asian Journal of Tropical Medicine and Public Health. 1985;16(2):228–33.

89 Mohammed, K. A., Haji, H. J., Gabrielli, A. F., Mubila, L., Biswas, G., Chitsulo, L. & Molyneux, D. H. (2008). Triple co-administration of ivermectin, albendazole and praziquantel in Zanzibar: a safety study. PLoS Neglected Tropical Diseases. 2(1), e171.

90 Mpairwe, H., Webb, E. L., Muhangi, L., Ndibazza, J., Akishule, D., Nampijja, M. & Elliott, A. M. Anthelminthic treatment during pregnancy is associated with increased risk of infantile eczema: randomised□controlled trial results. Pediatric Allergy and Immunology, 2011; 22(3), 305–312.

91 Muhumuza, S., Olsen, A., Katahoire, A., & Nuwaha, F. (2013). Uptake of Preventive Treatment for Intestinal Schistosomiasis among School Children in Jinja District, Uganda: A Cross Sectional Study. PLoS ONE, 8(5). http://doi.org/10.1371/journal.pone.0063438.

92 Mutapi, F., Rujeni, N., Bourke, C., Mitchell, K., Appleby, L., Nausch, N. & Mduluza, T. (2011). Schistosoma haematobium treatment in 1–5 year old children: safety and efficacy of the antihelminthic drug praziquantel. PLoS Neglected Tropical Diseases. 5(5), e1143.

93 Na-Bangchang, K., Kietinun, S., Pawa, K. K., Hanpitakpong, W., Na-Bangchang, C., & Lazdins, J. Assessments of pharmacokinetic drug interactions and tolerability of albendazole, praziquantel and ivermectin combinations. Transactions of the Royal Society of Tropical Medicine and Hygiene, 2006; 100(4), 335–345.

94 Nalugwa, A., Nuwaha, F., Tukahebwa, E. M., & Olsen, A. Single versus double dose praziquantel comparison on efficacy and *Schistosoma mansoni* re-infection in preschool-age children in Uganda: a randomized controlled trial. PLoS Neglected Tropical Diseases.2015; 9(5), e0003796.

95 Namwanje, H., Kabatereine NB and Olsen A (2011) The acceptability and safety of praziquantel alone and in combination with mebendazole in the treatment of Schistosoma mansoni and soil-transmitted helminthiasis in children aged 1 – 4 years in Uganda. Parasitology.138, 1586--1592.

96 Namwanje, H., Kabatereine NB and Olsen A (2011) A randomized controlled clinical trial on the safety of co-administration of albendazole, ivermectin and praziquantel in infected schoolchildren in Uganda. Transactions of the Royal Society of Tropical Medicine and Hygiene.105, 181–188.

97 Nash, T. E, Hofstetter. M, Cheever, A. W, Ottensen, E. A. (1982). Treatment of Shistosoma mekongi with praziquantel: A double-blind study. American Journal of Tropical Medicine and Hygiene. 31(3), 1982, pp972-982

98 Ndyomugyenyi, R., & Kabatereine, N. Integrated community directed treatment for the control of onchocerciasis, schistosomiasis and intestinal helminths infections in Uganda: advantages and disadvantages. Tropical Medicine & International Health. 2003; 8(11), 997–1004.

99 N’Goran EK, Gnaka HN, Tanner M and Utzinger J (2003) Efficacy and side-effects of two praziquantel treatments against Schistosoma haematobium infection, among schoolchildren from Cote D’Ivoire. Annals of Tropical Medicine and Parasitology. 97(1) 37-51

100 Njenga, S. M., Ng’Ang’a, P. M., Mwanje, M. T., Bendera, F. S., & Bockarie, M. J. (2014). A school-based cross-sectional survey of adverse events following co-administration of albendazole and praziquantel for preventive chemotherapy against urogenital schistosomiasis and soil-transmitted helminthiasis in Kwale County, Kenya. PLoS ONE, 9(2), 6–10.

101 Obonyo, C. O., Muok, E. M., & Mwinzi, P. N. Efficacy of artesunate with sulfalene plus pyrimethamine versus praziquantel for treatment of Schistosoma mansoni in Kenyan children: an open-label randomised controlled trial. The Lancet infectious diseases, 2010; 10(9), 603–611.

102 Olds, G. R., King, C., Hewlett, J., Olveda, R., Wu, G., Ouma, J., Peters, P., McGarvey, S., Odhiambo, O., Koech, D., Liu, C. Y., Aligui, G., Gachihi, G., Kombe, Y., Parraga, I., Ramirez, B., Whalen, C., Horton, R. J. and Reeve, P.(1999). Double-blind placebo-controlled study of concurrent administration of albendazole and praziquantel in school children with schistosomiasis and geohelminths. Journal of Infectious Diseases 179, 996– 1003

103 Olliaro, P. L., Vaillant, M. T., Belizario, V. J., Lwambo, N. J., Ould Abdallahi, M., Pieri, O. S., … & Chitsulo, L. (2011). A multicentre randomized controlled trial of the efficacy and safety of single-dose praziquantel at 40 mg/kg vs. 60 mg/kg for treating intestinal schistosomiasis in the Philippines, Mauritania, Tanzania and Brazil. PLoS Negl Trop Dis, 5(6), e1165.

104 Olveda RM, Acosta LP, Tallo V, Baltazar PI, Lesiguez JLS, Estanislao GG, Ayaso EB, Monterde DBS, Ida A, Watson N, McDonald EA, Wu HW, Kurtis JD, Friedman JF. ‘Efficacy and safety of praziquantel for the treatment of human schistosomiasis during pregnancy: a phase 2, randomised, double-blind, placebo-controlled trial’, Lancet Infectious Disease. 2016; 16(2), pp. 199–208.

105 Omer, AH. Praziquantel in the treatment of mixed *S. haematobium* and *S. mansoni* infections. Arzneimittelforschung. 1981; 31, 605–608.

106 Oyediran, A. B., Kofie, B. A., Bammeke, A. O. and Bamgboye, E. A. (1981). Clinical experience with praziquantel in the treatment of Nigerian patients infected with *S. haematobium*. Arzneimittelforschung 31, 581–584.

107 Page MJ, Moher D, Bossuyt PM, Boutron I, Hoffmann TC, Mulrow CD, et al. PRISMA 2020 explanation and elaboration: updated guidance and exemplars for reporting systematic reviews. Bmj. 2021;372:n160.

108 Polderman, A. M., Gryseels, B., Gerold, J. L., Mpamila, K., & Manshande, J. P. (1984). Side effects of praziquantel in the treatment of *Schistosoma mansoni* in Maniema, Zaire. Transactions of the Royal Society of Tropical Medicine and Hygiene, 78(6), 752–754.

109 Ouzzani M, Hammady H, Fedorowicz Z, Elmagarmid A. Rayyan—a web and mobile app for systematic reviews. Systematic reviews. 2016;5(1):1–10.

110 Qian, C., & Gong, F. (2016). Praziquantel for schistosomiasis in pregnancy. The Lancet Infectious Diseases, 16(5), 525–526.

111 Ramiandrasoa, N. S., Ravoniarimbinina, P., Solofoniaina, A. R., Andrianjafy Rakotomanga, I. P., Andrianarisoa, S. H., Molia, S., … & Rajaonatahina, D. (2020). Impact of a 3-year mass drug administration pilot project for taeniasis control in Madagascar. PLoS neglected tropical diseases, 14(9), e0008653.

112 Raso, G., N’Goran, E. K., Toty, A., Luginbühl, A., Adjoua, C. A., Tian-Bi, N. T., … & Utzinger, J. (2004). Efficacy and side effects of praziquantel against *Schistosoma mansoni* in a community of western Côte d’Ivoire. Transactions of the Royal Society of Tropical Medicine and Hygiene, 98(1), 18–27.

113 Reta, B., & Erko, B. (2013). Efficacy and side effects of praziquantel in the treatment for Schistosoma mansoni infection in school children in Senbete Town, northeastern Ethiopia. Tropical Medicine and International Health, 18(11), 1338–1343.

114 Rezende, G. L. D. Survey on the clinical trial results achieved in Brazil comparing praziquantel and oxamniquine in the treatment of mansoni schistosomiasis. Revista do Instituto de Medicina Tropical de São Paulo. 1985; 27(6), 328–336.

115 Rugemalila JB, Eyakuze VM. 1981. Use of metrifonate for selective population chemotherapy against urinary schistosomiasis in an endemic area at Mwanza, Tanzania. East African Medical Journal1981;58(1):37–43.

116 Santesso N, Glenton C, Dahm P, Garner P, Akl EA, Alper B, Brignardello-Petersen R, Carrasco-Labra A, De Beer H, Hultcrantz M, Kuijpers T, Meerpohl J, Morgan R, Mustafa R, Skoetz N, Sultan S, Wiysonge C, Guyatt G, Schünemann HJ; GRADE Working Group. GRADE guidelines 26: informative statements to communicate the findings of systematic reviews of interventions. J Clin Epidemiol. 2020;119:126–135.

117 Santos, A. T., Blas, B. L., Nosenas, J. S., Portillo, G. P., Ortega, O. M., Hayashi, M., & Boehme, K. (1979). Preliminary clinical trials with praziquantel in Schistosoma japonicum infections in the Philippines. Bulletin of the World Health Organization, 57(5), 793.

118 Santos, A. T., Blas, B. L., Portillo, G., Nosenas, J. S., Poliquit, O., & Papasin, M. Phase III clinical trials with praziquantel in *S. japonicum* infections in the Philippines. Arzneimittel-forschung, 1984; 34(9B), 1221–1223.

119 Schutte CH, Osman Y, Van Deventer JM, Mosese G. 1983. Effectiveness of praziquantel against the South African strains of *Schistosoma haematobium* and *S. mansoni*. South African Medical Journal (Suid-Afrikaanse Tydskrif Vir Geneeskunde) 1983; 64(1):7-101983;64(1):7–10.

120 Sissoko MS, Dabo A, Traore H, Diallo M, Traore B, Konate D, Niare B, Diakite M, Kamate B, Traore A, Bathily A, Tapily A, Traore OB, Cauwenbergh S, Jansen HF and Doumbo OK (2009) Efficacy of artesunate + sulfamethoxypyrazine/pyrimethamine versus praziquantel in the treatment of Schistosoma haematobium in children. PLoS One. 4(10):e6732 doi:10.1371/journal.pone.0006732.

121 Smith M, Clegg JA, Webbe G. 1981. Culture of *Schistosoma haematobium* in vivo and in vitro. Annals of Tropical Medicine and Parasitology;70(1):101–7

122 Sousa-Figueiredo, J. C., Pleasant, J., Day, M., Betson, M., Rollinson, D., Montresor, A., … Stothard, J. R. (2010). Treatment of intestinal schistosomiasis in Ugandan preschool children: best diagnosis, treatment efficacy and side-effects, and an extended praziquantel dosing pole. International Health, 2(2), 103–113.

123 Stelma, F. F., Talla, I., Sow, S., Kongs, A., Niang, M., Polman, K., … & Gryseels, B. (1995). Efficacy and side effects of praziquantel in an epidemic focus of *Schistosoma mansoni*. The American journal of tropical medicine and hygiene, 53(2), 167–170.Sterne JA, Hernan MA, Reeves BC, et al. ROBINS-I: a tool for assessing risk of bias in non-randomised studies of interventions. BMJ 2016; 355: i4919.

124 Sterne JAC, Hernán MA, McAleenan A, Reeves BC, Higgins JPT. Chapter 25: Assessing risk of bias in a non-randomized study. In: Higgins JPT, Thomas J, Chandler J, Cumpston M, Li T, Page MJ, Welch VA (editors). Cochrane Handbook for Systematic Reviews of Interventions version 6.2 (updated February 2021). Cochrane, 2021. Available from www.training.cochrane.org/handbook.

125 Stroup DF, Berlin JA, Morton SC, Olkin I, Williamson G, Moher D, et al. Meta-analysis of Observational Studies in Epidemiology: A Proposal for Reporting - Meta-analysis Of Observational Studies in Epidemiology (MOOSE) Group B. JAMA Neurol. 2000;283:2008–12.

126 Sukwa, T. Y. (1993). A community-based randomized trial of praziquantel to control schistosomiasis morbidity in schoolchildren in Zambia. Annals of Tropical Medicine & Parasitology, 87(2), 185–194.

127 Taddese, K., & Zein, Z. A. (1988). Comparison between the efficacy of oxamniquine and praziquantel in the treatment of *Schistosoma mansoni* infections on a sugar estate in Ethiopia. Annals of Tropical Medicine & Parasitology, 82(2), 175–180.

128 Tweyongyere R, Mawa PA, Ngom-Wegi S, Ndibazza J, Duong T, Vennervald BJ, Dunne DW, Katunguka-Rwakishaya E, Elliott AM. Effect of praziquantel treatment during pregnancy on cytokine responses to schistosome antigens: results of a randomized, placebo-controlled trial. Journal of Infectious Diseases. 2008 Dec 15;198(12):1870–9. doi: 10.1086/593215.

129 Tweyongyere R, Mawa PA, Kihembo M, Jones FM, Webb EL, Cose S, Dunne DW, Vennervald BJ, Elliott AM. Effect of praziquantel treatment of Schistosoma mansoni during pregnancy on immune responses to schistosome antigens among the offspring: results of a randomised, placebo-controlled trial. BMC Infect Dis. 2011 Sep 2;11:234. doi: 10.1186/1471-2334-11-234.

130 Tweyongyere R, Naniima P, Mawa PA, Jones FM, Webb EL, Cose S, Dunne DW, Elliott AM. Effect of maternal Schistosoma mansoni infection and praziquantel treatment during pregnancy on Schistosoma mansoni infection and immune responsiveness among offspring at age five years. PLoS Neglected Tropical Diseases. 2013 Oct 17;7(10):e2501.

131 Watt, G., White, N. J., Padre, L., Ritter, W., Fernando, M. T., Ranoa, C. P., & Laughlin, L. W. (1988). Praziquantel pharmacokinetics and side effects in *Schistosoma japonicum*-infected patients with liver disease. Journal of Infectious Diseases, 157(3), 530–535.

132 Watt, G. Bloody diarrhea after praziquantel therapy. Transactions of the Royal Society of Tropical Medicine and Hygiene. 1986; 80 pp345. Watt G, Adapon B, Long GW, Fernando MT, Ranoa CP, Cross JH. Praziquantel in treatment of cerebral schistosomiasis. Lancet. 1986; 6;2(8506):529-32. doi: 10.1016/s0140-6736(86)90110-8.

133 Steinmann P, Keiser J, Bos R, Tanner M, Utzinger J. Schistosomiasis and water resources development: systematic review, meta-analysis, and estimates of people at risk. Lancet Infect Dis. 2006 Jul;6(7):411–25. doi: 10.1016/S1473-3099(06)70521-7.

134 Sterne JA, Hernán MA, Reeves BC, Savović J, Berkman ND, Viswanathan M, Henry D, Altman DG et al. ROBINS-I: a tool for assessing risk of bias in non-randomised studies of interventions. British Medical Journal. 2016;355:i4919. doi: 10.1136/bmj.i4919.

135 Sturrock RF. Schistosomiasis epidemiology and control: how did we get here and where should we go? Mem Inst Oswaldo Cruz. 2001;96 Suppl:17–27.

136 WHO 2012a. Schistosomiasis: Progress Report 2001-2011 and Strategic Plan 2012-2020.

137 WHO 2012b. Accelerating work to overcome the global impact of neglected tropical disease: a road map for implementation. WHO/HTM/NTD/2012, 37 pp.

138 WHO. Elimination of schistosomiasis. Sixty-fifth World Health Assembly Resolution WHA 65.21. http://www.who.int/neglected_diseases/mediacentre/WHA_65.21_Eng.pdf

139 WHO. Schistosomiasis and soil-transmitted helminthiases: number of people treated in 2020. Weekly Epidemiological Record, 2016,9;91(49-50):585-95.

140 WHO. Schistosomiasis and soil-transmitted helminthiases: progress report, 2020. Weekly Epidemiological Record, No 48, 2021, 96, 585–595

141 WHO. WHO guideline on control and elimination of human schistosomiasis. February 2022.

142 Hou XY, McManus DP, Gray DJ, Balen J, Luo X-S, He Y-K, Ellis M, Williams GM, Li YS (2008). A randomized, double-blind, placebo-controlled trial of safety and efficacy of combined praziquantel and artemether treatment for acute schistosomiasis japonica in China. Bulletin of the World Health Organization, 86(10), 788–95.

143 WHO. Ending the neglect to attain the Sustainable Development Goals – A road map for neglected tropical diseases 2021–2030. Geneva: World Health Organization; 2020. Licence: CC BY-NC-SA 3.0 IG

144 WHO. Preventive chemotherapy data portal. Geneva: World Health Organization; 2021 (https://www.who.int/data/preventive-chemotherapy, accessed November 2021).

145 WHO. The control of schistosomiasis: Second report of the WHO Expert Committee (WHO technical report series; 912). Geneva: World Health Organization, 2002.

146 WHO. Prevention and control of schistosomiasis and soil-transmitted helminthiasis: A Report of WHO Expert Committee. 2006 (WHO Technical Report Series 912) Geneva: World Health Organization, 2006

147 147 WHO. Accelerating work to overcome the global impact of neglected tropical diseases : a roadmap for implementation World Health Organization, 2012.

148 148 WHO. Schistosomiasis: progress report 2001 - 2011, strategic plan 2012 - 2020: World Health Organization, 2013.

149 149 WHO. Safety in administering medicines for neglected tropical diseases. Geneva: World Health Organization; 2021 (https://www.who.int/publications/i/item/9789240024144, accessed November 2021).

150 150 WHO. Global health estimates 2016: Disease burden by cause, age, sex, by country and by region, 2000–2016. Geneva: World Health Organization; 2018 (http://www.who.int/healthinfo/global_burden_disease/estimates/en/index1.html, accessed November 2021).

151 151 WHO. Global health estimates 2019: Disease burden by cause, age, sex, by country and by region, 2000–2019. Geneva: World Health Organization; 2020 (https://www.who.int/data/gho/data/themes/mortality-and-global-health-estimates, accessed November 2021).

152 152 WHO. Preventive chemotherapy data portal. Geneva: World Health Organization; 2021 (https://www.who.int/data/preventive-chemotherapy, accessed November 2021).

153 Wilkins, H. A. and Moore, P. J. (1987). Comparative trials of regimes for the treatment of urinary schistosomiasis in The Gambia. Journal of Tropical Medicine and Hygiene 90, 83–92.

154 154 WHO. WHO guideline on control and elimination of human schistosomiasis. 2022, ISBN 978-92-4-004160-8

155 Woldemichael T, Wondimagegnehu T. Schistosoma haematobium treatment with praziquantel: preliminary clinical observations. Ethiop Med J. 1986 Jul;24(3):155–6.

156 Wu, Ming-He, Chen-Ci Wei, Zhao-Yue Xu, Hong-Chang Yuan, Wei-Neng Lian, Qiu-Ji Yang, Min Chen et al. Comparison of the therapeutic efficacy and side effects of a single dose of levo-praziquantel with mixed isomer praziquantel in 278 cases of schistosomiasis japonica. The American journal of tropical medicine and hygiene, 45, no. 3 (1991): 345–349.

157 Wu, M.H., Wei, C.C., Xu, Z.Y., Yuan, H.C., Lian, W.N., Yang, Q. J., Chen, M., Jiang, Q.W., Wang, C.Z., Zhang, S.J., Liu, Z-D., Wei, R.M., Yuan, S.J., Hu, L.S. and Wu, Z.S. Comparison of the therapeutic efficacy and side effects of a single dose of levo-praziquantel with mixed isomer praziquantel in 278 cases of schistosomiasis japonica. American journal of tropical medicine and hygiene. 1991;45(3):345–9.

158 von Elm E, Altman DG, Egger M, et al. The Strengthening the Reporting of Observational Studies in Epidemiology (STROBE) statement: guidelines for reporting observational studies. Lancet 2007; 370(9596): 1453–7.

